# Evaluating the Impact and Cost-Effectiveness of Typhoid Conjugate Vaccine Schedules Across Diverse Settings: A Multi-Model Comparison

**DOI:** 10.64898/2026.03.09.26346651

**Authors:** Catherine G.C. Wenger, Kyra H. Grantz, Tigist F. Menkir, Alina M. Muellenmeister, Zeaan Pithawala, Raymond Hutubessy, Vittal Mogasale, Alicia N.M. Kraay, Nick Scott, Romesh G. Abeysuriya, Jason R. Andrews, Jillian Gauld, Nathan C. Lo, Virginia E. Pitzer

## Abstract

**Background:** Given emerging evidence on the waning of immunity from typhoid conjugate vaccines (TCV), the World Health Organization (WHO) commissioned a multi-model comparison to determine the optimal schedule in terms of health and economic impact to inform updated recommendations for TCV use across different settings.

**Methods and findings:** To identify optimal vaccination strategies across different incidence settings and vaccine waning assumptions, we compared two agent-based and two compartmental dynamic models of typhoid transmission. All models were fitted to harmonized age-specific incidence data from medium, high, and very high incidence settings. We assessed different TCV schedules under slow- and fast-waning scenarios to evaluate the best age for routine vaccination and the potential need for booster doses and catch-up campaigns. We evaluated the public health and economic impact predicted for each model and scenario using the net-monetary-benefit framework to determine cost-effectiveness under two representative scenarios for the health outcomes and costs of vaccination and treatment. Over a 10-year time horizon, routine vaccination at 9 months with a catch-up campaign to 15 years and a booster dose at 5 years was predicted to have the greatest impact, reducing cases by a median of 48-64% across the incidence settings. Across all four models, TCV introduction with a catch-up campaign was cost-effective at willingness-to-pay (WTP) thresholds >$1,250 per disability-adjusted life-year (DALY) averted in medium incidence settings when costs and case-fatality risk (CFR) are high and in high incidence settings when costs and CFR are low. The optimal strategy was to delay vaccination to 2 or 5 years of age if waning is fast, depending on the age of peak incidence. In very high incidence settings, TCV introduction at 9 months or 2 years of age was cost-saving, and adding a booster dose at 5 years was cost-effective at most WTP values across all scenarios.

**Conclusions:** Model predictions for the impact and cost-effectiveness of different TCV schedules were fairly robust to uncertainty in parameter values and model structure, but the optimal strategy depends on the typhoid incidence rate, CFR, and waning rate of vaccine protection.

## Introduction

Typhoid fever, an acute febrile illness caused by the bacterial pathogen *Salmonella enterica* serovar Typhi (*S*. Typhi), causes an estimated 7.2-12.0 million cases and 56,000-181,000 deaths worldwide [1]. *S*. Typhi is transmitted through the fecal-oral route, and despite being eliminated in high-income countries with adequate water and sanitation infrastructure, typhoid fever remains a major public health concern in low- and middle-income countries (LMICs). The majority of the disease burden occurs in children under 15 years old, and incidence peaks in 2-4-year-olds in high-incidence settings [2]. While incidence and mortality due to typhoid fever have declined with improved clinical management and antibiotic therapy, the emergence of antimicrobial resistance (AMR) threatens to reverse these gains [3]. Thus, given the growing threat from AMR, interventions to maximize prevention of typhoid fever are critical [3–4].

In 2018, the World Health Organization (WHO) recommended the introduction of the typhoid conjugate vaccine (TCV) into the childhood routine immunization programs for infants and children aged 6 months to 15 years old in endemic countries, but vaccine adoption has been slow [5]. This was the first time typhoid vaccines were recommended for routine use in highly endemic settings. To support this decision, prior studies found that TCVs may be cost-effective in certain settings [6–9].

Emerging data have demonstrated potential waning of vaccine-derived protection, which necessitates consideration of a revised vaccine schedule, including whether to add booster doses of TCV. Vaccine efficacy data from TCV trials in children less than 15 years in Malawi and Bangladesh show differences in the duration of vaccine protection, potentially influenced by the force of infection in each setting [10–11]. While a high vaccine efficacy (>75%) was maintained over four and a half years of follow-up in Malawi, protection declined to ∼50% three to five years after vaccination in Bangladesh and was even lower (24%) among those <2 years old at inoculation [10–13].

Considering heterogeneous typhoid epidemiology, TCV waning, and increasingly constrained resources, which potentially limit childhood immunization efforts, re-assessing vaccine impact under different conditions can guide decision-makers to identify the optimal vaccine strategy for their local context.

Mathematical models are important for informing vaccine introduction decisions because they can simulate disease dynamics and predict outcomes under a range of intervention settings where there may be limited empirical data. However, no single model can capture all uncertainties in the dynamics influencing typhoid epidemiology. Comparing the results of multiple models can inform the robustness of model predictions and indicate which assumptions and parameters have the greatest influence on the simulated impacts [14]. To inform the deliberations of the WHO Strategic Advisory Group of Experts (SAGE) on updated global recommendations for the TCV vaccine schedule, including whether to include a booster dose in endemic settings, the WHO commissioned a multi-model comparison to identify the optimal TCV schedule in terms of disease and economic impact under the WHO Multi-Model Comparisons for Typhoid Conjugate Vaccine Adequate Schedules (MMC-TAS) project. During the project, the WHO Immunization and Vaccines Related Implementation Advisory Committee (IVIR-AC) reviewed the robustness and quality of the modeling methods and final results [15–16].

We compared four mathematical models estimating the impact and cost-effectiveness of potential TCV vaccination schedules in multiple epidemiologic, vaccine waning, and cost settings. The models used harmonized input data and delivery scenarios to predict vaccine impact while maintaining unique structural features and assumptions to allow for within- and between-model uncertainty. Following WHO guidance regarding the use of cost-effectiveness analysis for decision-making on vaccine introduction and multi-model comparisons, we systematically compared the health and economic impact of the four typhoid models to identify the optimal strategy under different baseline incidence, vaccine waning, and economic scenarios to inform updated WHO SAGE recommendations [14,17].

## Methods

### Delivery strategies and scenarios

We evaluated five potential vaccine delivery strategies over a 10-year time horizon: (1) no vaccination, (2) routine vaccination at 9 months, (3) routine vaccination at 2 years, (4) routine vaccination at 5 years, and (5) routine vaccination at 9 months with a booster dose at 5 years. Each strategy included a four to twelve-week catch-up campaign upon introduction for individuals at the age of routine vaccination, up to 15 years.

To understand the range of potential benefits across settings, we conducted analyses for multiple scenarios based on disease incidence, waning of vaccine effectiveness, and treatment and vaccination costs. We modeled three age-specific incidence archetypes: (1) medium incidence (10-99 cases per 100,000 person-years), (2) high incidence (100-499 cases per 100,000 person-years), and (3) very high incidence (500+ cases per 100,000 person-years) [18]. For each incidence setting, we predicted TCV impact under slow and fast waning scenarios based on the Malawi and Bangladesh clinical trials (details below). We additionally evaluated two scenarios, representing lower and higher disease severity and vaccination/treatment costs. While we generalize the scenario names to “Africa” (higher severity/higher costs) and “Asia” (lower severity/lower costs) regional settings, the data informing these scenarios are from a subset of countries within these regions. In total, this yielded 12 scenario combinations (Figure S6.1.1). For this analysis, we do not account for funding support from Gavi when estimating vaccine procurement or delivery costs.

### Transmission model descriptions and harmonized assumptions

Four modelling groups contributed to this study: the Burnet Institute (Burnet), the Institute for Disease Modeling (IDM), Stanford University (Stanford), and Yale University (Yale). All models are age-structured and have an SIR-like structure and include a chronic carrier state (in which individuals are susceptible, can become infectious, and either recover with natural immunity or develop chronic infection) to simulate typhoid transmission dynamics with and without vaccination. Two models are agent-based (Burnet and IDM), modeling disease transmission and progression on an individual level, and two models are compartmental (Stanford and Yale), modeling the flow of a population through different infection states. The models included unique features, such as maternal immunity (Burnet, IDM), environmental transmission (IDM), 10-year duration (Stanford) versus lifelong (Burnet, IDM, Yale) chronic carriage, and differentiation between primary and secondary infections (Yale). We harmonized assumptions and input data for the population demographics, baseline typhoid incidence, vaccine efficacy, and vaccine coverage (Table 1). Full model descriptions and diagrams can be found in the supplement.

**Table 1.** Parameters and assumptions harmonized across models.

| Harmonized parameters and assumptions |  |
| --- | --- |
| Demographics | Assumed a roughly constant population size with the age composition of a low-income country [19]<br><u>Crude birth rate</u>: 22.8 births per 1000 persons per year [19]<br><u>Average life expectancy</u>: 64 years (Africa) and 75 years (Asia) [19] |
| Incidence* | <u>Medium incidence</u>: annual incidence of 52 cases per 100,000 person-years, peak incidence in the 10-15-year age category<br><u>High incidence</u>: annual incidence of 214 cases per 100,000 person-years, peak incidence in the 2-4-year age category<br><u>Very high incidence</u>: annual incidence of 1,255 cases per 100,000 person-years, peak incidence in the 2-4-year age category |
| Vaccine delivery strategies | No vaccination<br>Routine vaccination at 9 months with a catch-up campaign 9 months to 15 years<br>Routine vaccination at 2 years with a catch-up campaign 2 to 15 years<br>Routine vaccination at 5 years with a catch-up campaign 5 to 15 years<br>Routine vaccination at 9 months + a booster at 5 years with a catch-up campaign up to 15 years; booster doses only administered to previously vaccinated individuals |
| Vaccine coverage | <u>9 months</u>: Based on MCV1<sup>†</sup> coverage estimates for low- and lower-middle-income countries<br><u>2 years</u>: Uniform(0.5, 1) x MCV2<sup>†</sup> coverage estimates<br><u>5 years</u>: Uniform(0.5, 1) x school attendance estimates<br><u>Catch-up campaign</u>: Based on Supplementary Immunization Activity estimates |
| Vaccine effectiveness<sup>§</sup> | <u>Slow-waning</u>: Based on trial data from Malawi [10,12]<br>*Age at vaccination <5 years*:<br>74.4% (1.75 years follow-up), 70.6% (4.61 years follow-up)<br>*Age at vaccination 5-15 years*:<br>83.7% (1.75 years follow-up), 79.3% (4.61 years follow-up)<br><u>Fast-waning</u>: Based on trial data from Bangladesh [11,13]<br>*Age at vaccination <2 years*:<br>81% (1-2 years follow-up), 24% (3-5 years follow-up), -5% (5-7 years follow-up)<br>*Age at vaccination 2-4 years*:<br>80% (1-2 years follow-up), 59% (3-5 years follow-up), 14% (5-7 years follow-up)<br>*Age at vaccination 5-15 years*:<br>88% (1-2 years follow-up), 74% (3-5 years follow-up), 32% (5-7 years follow-up) |
| Case-fatality risk | <u>Africa</u>: 0.81% (95% confidence interval: 0.10%, 6.22%) [3]<sup>#</sup><br><u>Asia</u>: 0.16% (95% confidence interval: 0.07%, 0.40%) [3]<sup>#</sup> |
| Treatment probabilities | Region-specific; see supplement Table 6.3.1 |
| Cost of treatment | Age-, treatment-, and region-specific; see supplement Table 6.3.1<br><u>Pediatric</u>: ≤15 years old<br><u>Adult</u>: >15 years old |
| Cost of TCV vaccination | <u>Procurement cost</u>: \$1.50, without Gavi support [20]<br><u>Syringes & safety equipment</u>: average of \$0.29 [21]<br><u>Delivery cost</u>: strategy-, and region-specific; see supplement Table 6.3.1 |

\* See Supplement 2 for age-specific incidence.
<sup>†</sup> MCV1, measles containing vaccine first dose; MCV2, measles containing vaccine second dose.
<sup>§</sup> Vaccine effectiveness estimates used for model fitting. See Table S3.1 for model-specific parameter estimates.
<sup>#</sup> Based on a sub-analysis of studies published since 2010.

To achieve a stable population and age structure, the models were fit to the demographic characteristics of low- and lower-middle-income countries as reported by the United Nations World Population Prospects [19]. Baseline age-specific incidence of symptomatic typhoid fever was based on a review of published incidence rates with case counts and person-time adjusted for healthcare seeking, enrollment, and blood-culture sensitivity (see Tables S2.1-2, and Figure S2.1), subset to studies that reported data for all age categories. Transmission and natural history parameters for each model were estimated to reproduce both the magnitude and age distribution of incidence for each of the three incidence archetypes (see Supplement Sections 2 and 5).

For the slow-waning scenario, we estimated age-specific vaccine efficacy (VE) and duration of protection for ages 9 months to <5 years and 5 to 15 years based on the TyVAC-Malawi randomized controlled trial (RCT) following 2 years and 4.6 years of follow-up (Table 1) [10,12]. Estimates for the fast-waning scenario for ages 9 months to <2 years, 2 to <5 years, and 5 to 15 years at vaccination were derived from the TyVAC-Bangladesh cluster RCT following 2 years of follow-up [13] and the TyVOID-Bangladesh study for 3-5 years [11] and 5-7 years (unpublished) of follow-up (Table 1). The IDM and Yale groups used a statistical model implemented using Markov Chain Monte Carlo methods to estimate the initial vaccine efficacy (VE_0_) and average duration of protection (1/w_v_) by fitting to the mean and 95% confidence interval (CI) of the VE estimates, while the Burnet and Stanford groups estimated VE_0_ and w_v_ by fitting the transmission models to the observed mean VE estimates over time. Each of the models was then either validated against or calibrated to reproduce the total (85%, 97.5% CI: 76% to 91%), overall (57%, 97.5% CI: 43% to 68%), and indirect (19%, 97.5% CI: -12% to 41%) effects observed in the Bangladesh RCT [13]. Strategy-specific vaccine coverage was based on historical estimates for measles-containing vaccine (MCV) coverage, supplementary immunization campaign MCV coverage, and school attendance (Supplement 4).

### Economic models

Each epidemiological transmission model is accompanied by an economic decision-tree model that calculates vaccine implementation costs, treatment costs, and health outcomes under each vaccine strategy. The economic models had similar structures (Figure S6.2.1) and sampled from harmonized uncertainty distributions for each stochastic parameter. Yearly symptomatic incidence data from the epidemiological simulations of each model were used as inputs for the cost-effectiveness models in addition to other disease severity and cost parameters. These parameters are sourced from specific country or multi-country studies in Africa and Asia, which we then generalize to each region for this analysis. We conducted the analysis from the healthcare payer perspective and did not account for indirect costs such as transportation and lost wages.

Of those with symptoms, we assumed a fraction sought care and either received outpatient or inpatient treatment, and a fraction of hospitalized cases was assumed to experience complications. We used region-specific estimates for the care-seeking probabilities, case fatality risks (CFR), and vaccine delivery and treatment costs, drawing from available country-level surveillance and costing cohort data available in each region. For the Africa region, represented countries included Malawi and those included in the Severe Typhoid in Africa programme (SETA) and the Strategic Typhoid Alliance across Africa and Asia (STRATAA). For the Asia region, represented countries included India (multiple sites from the Surveillance for Enteric Fever in India project), those included in the Surveillance Fever in Asia Project (SEAP), and those included in STRATAA. Given a paucity of data on how the CFR varies by treatment status and AMR, we applied overall estimates of the CFR to all symptomatic cases. The overall CFR for each region was estimated using data from Murthy et al. [3], restricting to studies published since 2010 to conservatively account for declines in the CFR over time.

Direct medical costs for outpatient and inpatient treatment are age-stratified by pediatric (≤15 years old) and adult (>15 years old) populations. We assumed the same treatment cost distributions for inpatients with and without complications due to a lack of stratified data, but separate disability weights were applied. There was also insufficient data to estimate costs and probabilities associated with AMR; the estimates we used encompass both antibiotic-resistant and sensitive cases, as the source studies were primarily conducted in locations with a high prevalence of multi-drug resistant or fluoroquinolone non-susceptible cases.

Vaccination costs included region-specific estimates of delivery costs for routine and supplementary immunization activity doses, school-based delivery costs for booster doses, and the costs of syringes and disposal (Table S6.3.1). Delivery costs were based on modeled country-specific estimates from Bilcke et al., combining estimates across countries captured in each of the two cost regions [6]. We assumed a procurement cost of $1.50 per dose, which is the market price of TCVs manufactured by Bharat Biotech and Biological E [20].

Harmonized outputs included: cases, hospitalizations, deaths, disability-adjusted life-years (DALYs), treatment costs, and vaccine costs. Outcomes from each model were summarized using medians and 95% prediction intervals in the supplement, as well as combined into an overall median and range of the model medians in Table 2. All costs were reported in 2025 USD, and both costs and DALYs were discounted at an annual rate of 3%, following WHO recommendations [17].

### Cost-effectiveness analysis

For the cost-effectiveness analysis, we used the net benefits framework to assess the optimal vaccine strategy from the health system perspective across a range of willingness-to-pay (WTP) thresholds from $0 to $2,500 (approximately equal to the average GDP per capita of a lower-middle-income country) [22]. Net monetary benefit was calculated for each strategy in all simulations using incremental costs and DALYs relative to no vaccination.

To capture uncertainty, we estimated the probability that each strategy had the highest net benefit at each WTP value across stochastic simulations. These results were visualized as cost-effectiveness acceptability frontiers (CEAFs) to illustrate the probability that the optimal strategy at each WTP value has the highest net benefit. To show how preferred strategies change across incidence settings, WTP values, and models, the CEAFs are presented as heatmaps where the dominant strategy and the uncertainty are indicated by color and shade, respectively. Cost-effectiveness acceptability curves (CEACs), which show the probabilities of the highest net benefit for all strategies at each WTP for each model, can be found in Supplement 7.3.

### Sensitivity analyses

We also performed a threshold analysis to determine how the optimal strategy varied depending on the baseline incidence level (and age distribution) and WTP threshold. By modeling the cost-effectiveness frontier across a range of values for the baseline incidence, but preserving the age distribution of cases for each incidence setting, we demonstrated how the age of peak incidence influences the preferred strategy.

To identify the most influential parameters in each model, we conducted probabilistic one-way sensitivity analyses (OWSA) using the *dampack* R package (version 1.0.2.1000) [23]. The OWSA is conducted using the simulated incremental costs and net benefits of each strategy, along with the corresponding parameter samples, in a regression-based framework to estimate how the outcomes change across each parameter’s range, holding all other variables constant. Additional scenario analyses, including varying the age at first vaccination, the time-horizon of the analysis, and evaluating strategies without catch-up campaigns, are included in the supplement.

## Results

All four transmission models were able to reproduce the age distribution observed in the harmonized age-specific incidence archetypes (Figures S5.2.2, S5.3.2, S5.4.2, and S5.5.2). In medium incidence settings, incidence peaks in the 5- to 14-year age groups, while in high and very high incidence settings, peak incidence occurs in the 2-4-year age group, consistent with a higher force of infection. The models also captured the differences in VE by duration of follow-up as well as the indirect and overall effectiveness observed in the TyVAC-Bangladesh cluster RCT, although the four groups took different approaches to calibrating the models to the VE data (see Supplement section 5).

### Vaccine impact

Over a 10-year time horizon, all vaccination strategies were predicted to provide a positive public health impact, reducing cases, hospitalizations, and deaths compared to no vaccination. Figure 1 shows the model-predicted annual incidence of typhoid fever over 20 years in the high incidence setting for no vaccination, routine vaccination at 9 months, and routine vaccination at 9 months with a booster at 5 years; results for the medium and very high incidence settings are given in the Supplement (Figures S7.1.1, S7.1.3). The 20-year horizon is highlighted here to show additional waning that was predicted by some models later in the simulations. The IDM model predicted increasing incidence for the baseline no-vaccination strategy (likely due to the long time needed to reach quasi-equilibrium), while all other models simulate constant incidence over time with no vaccination (Figure 1a,b). For both waning scenarios, there was an initial drop in cases driven by the catch-up campaign, followed by a sustained reduction in cases below baseline in the slow-waning scenario and a rebound of cases in the fast-waning scenario. Adding a booster dose at 5 years in the fast-waning scenario led to more sustained vaccine benefits than routine vaccination alone. Routine vaccination at 9 months had the greatest overall impact compared to routine vaccination at 2 or 5 years in the slow-waning scenario due to the broader age range included in the catch-up campaign (Supplement 7.1). In the fast-waning scenario, the 9-month strategy had the greatest initial impact, but routine vaccination at 5 years averted more cases over time due to the slower waning rate of VE estimated for older children. The number of vaccine doses administered per 100,000 individuals varied across the four models and for each strategy, likely due to differences in the population size of the vaccine-eligible age groups (Figure 1c). All models predict a slower rebound in cases in the medium incidence setting and a faster rebound in the very high incidence setting for both waning scenarios (Supplement 7.1).

**Figure 1.**
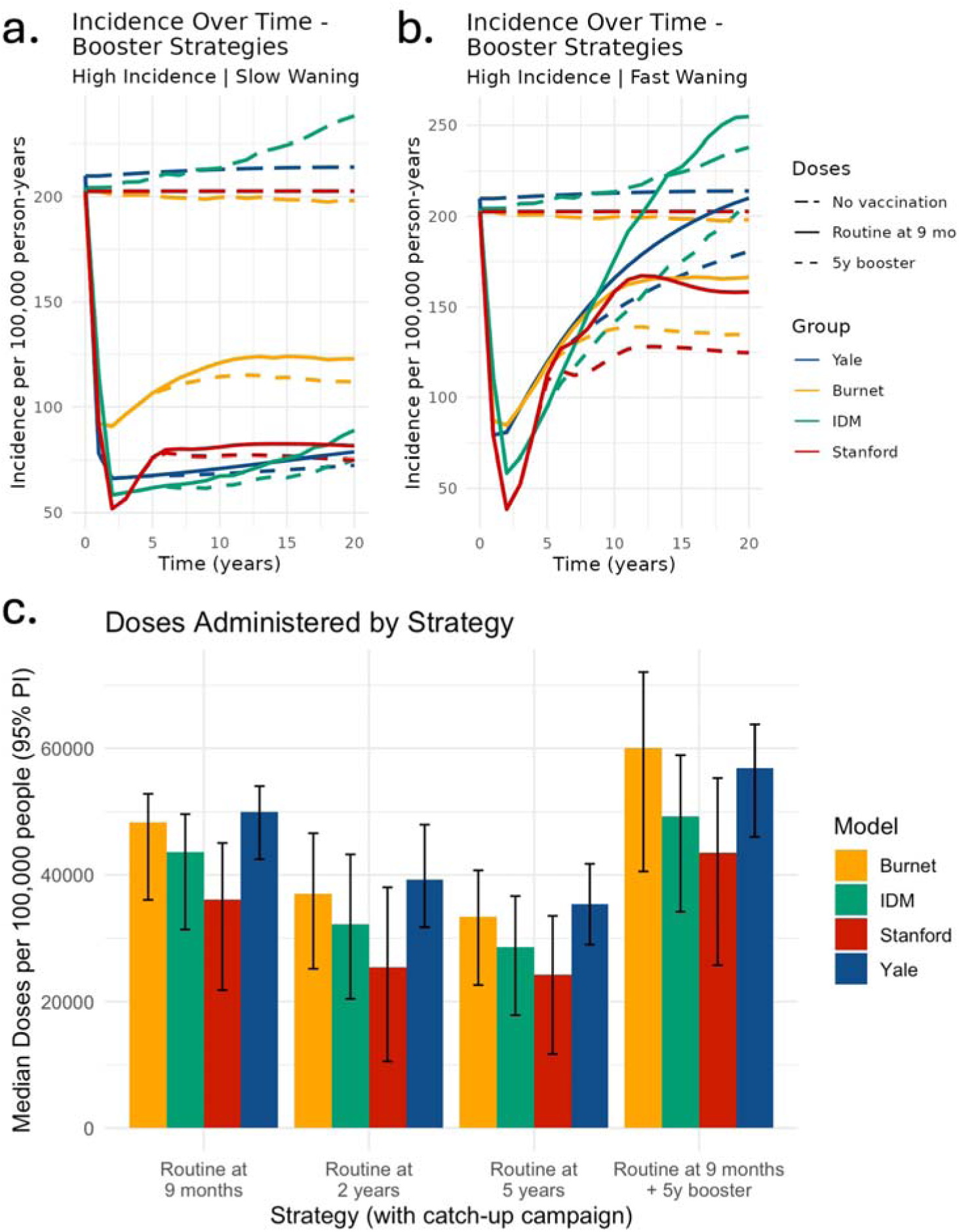
Model-predicted vaccine impact over time and doses administered per strategy for each model. The incidence curves (a,b) show the differences in vaccine impact between models under routine vaccination at 9 months with and without a booster at 5 years for the (a) slow-waning and (b) fast-waning scenario over a 20-year time horizon to illustrate the effects of waning in our models. (c) The median and 95% prediction intervals for the total doses administered per 100,000 people are plotted for each model and delivery strategy over the 10-year time horizon of our primary analysis.

Among the four models, the Stanford model predicted the largest vaccine impact in terms of typhoid cases averted in the medium and high incidence settings and the smallest impact in the very high incidence setting relative to the other models (Figure 2a); it also exhibited the most uncertainty in vaccine impact (Figure 2b). The Burnet model consistently predicted lower vaccine impact compared to the IDM and Yale models due to faster waning of vaccine protection estimated for both waning scenarios compared to the other models (Table S3.1). The IDM and Yale models predicted similar vaccine impact across all scenarios.

**Figure 2.**
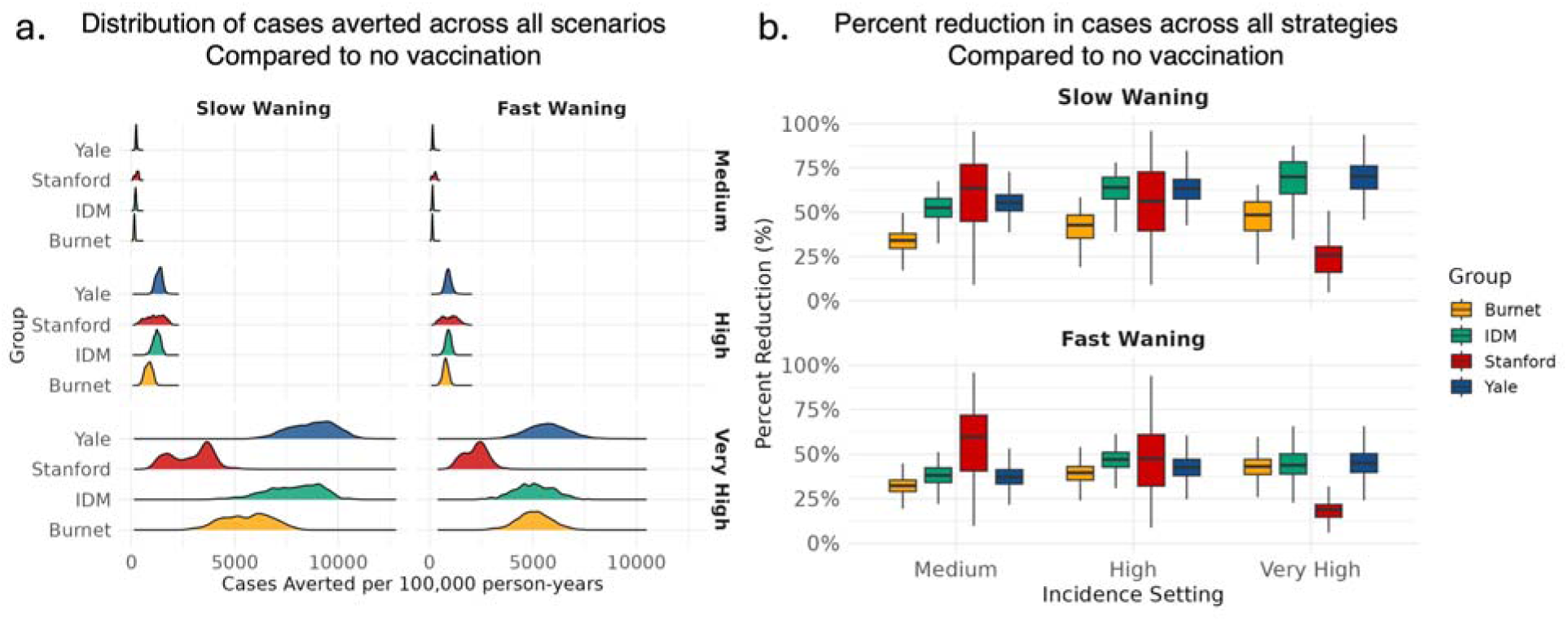
Distribution of the modeled impact of all vaccine strategies combined for each model. (a) Distribution of cases averted per 100,000 person-years for each model (Burnet, yellow; IDM, green; Stanford, red; Yale, blue) across all vaccine simulations compared to no vaccination by incidence and waning scenario. (b) Median and inter-quartile range of the percent reduction in cases across all vaccine simulations for the slow (top) and fast (bottom) waning scenarios and medium (left), high (middle), and very high (right) incidence settings.

The median number of cases, hospitalizations, deaths, and DALYs averted per 100,000 people over a 10-year time horizon is presented in Table 2, along with the range of median values predicted by each model. Among the strategies, the TCV booster strategy had the highest median impact, i.e., adding a booster dose would maximize the public health impact of TCV introduction. Under this strategy, the models estimated a 44-56% reduction in cases and 10 to 80 DALYs averted per 100,000 people in medium incidence settings, a 48-64% reduction in cases (52 to 457 DALYs averted) in high incidence settings, and a 42-61% reduction in cases (272 to 2,625 DALYs averted) in very high incidence settings depending on the region and waning scenario. Under slow waning, the difference between routine vaccination at 9 months and adding a booster dose at 5 years is minimal (an additional 1% median reduction). Among the alternative vaccine schedules without a booster dose, routine vaccination at 5 years had the lowest median impact, reducing cases by 34-42% and averting 222 to 1,809 DALYs in the very high incidence settings.

Considering the total costs of each strategy, treatment costs were greatest under the baseline no-vaccination strategy and lowest under the booster strategy across all scenarios (Table 2). In very high incidence settings, the relative treatment costs of the routine-only strategies varied by waning scenario; routine vaccination at 9 months with a catch-up campaign resulted in the lowest treatment costs when waning was slow, while routine vaccination at 2 years with a catch-up campaign resulted in the lowest costs when waning was fast. Conversely, vaccine procurement and delivery costs were greatest for the booster strategy at about $145,000 per 100,000 people in Africa and $131,000 per 100,000 people in Asia (Table 2). Routine vaccination at 5 years with a catch-up, which covered the smallest proportion of the total population, cost the least at $76,000-$79,000 per 100,000 people.

**Table 2.**
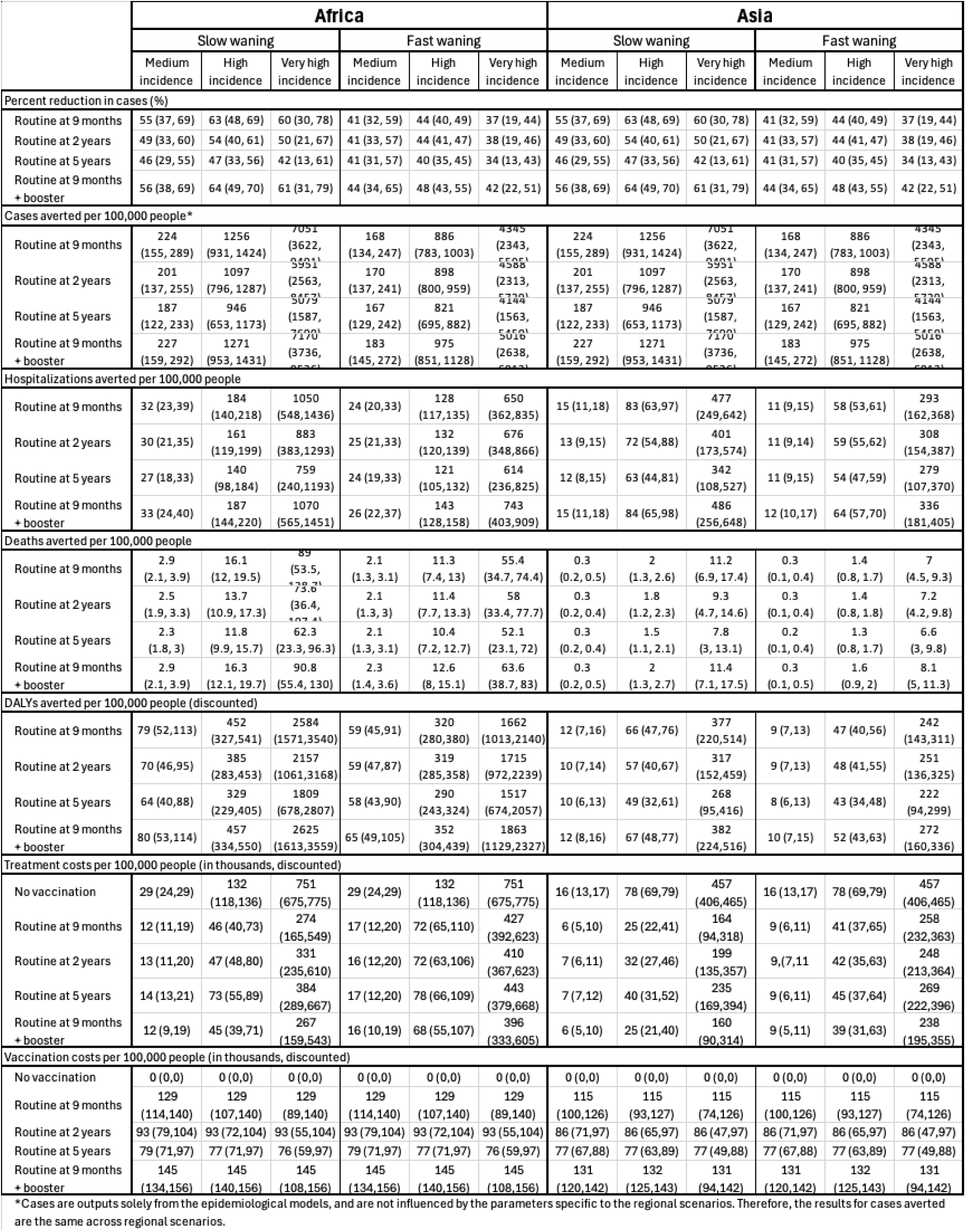
Health and economic outcomes by vaccination strategy across incidence and vaccine waning scenarios, by cost/CFR region (Africa vs Asia), over a 10-year time horizon. Values are presented as the median and range of all the model medians. Incremental metrics are the difference between the impact of the given strategy and no vaccination. Deaths are rounded to the nearest tenth, and all other values are rounded to the nearest whole number.

### Cost-effectiveness

Figures 3 and 4 show the preferred vaccination strategies across WTP thresholds from $0 to $2,500 per DALY averted for the four models and the different incidence settings and waning scenarios [24]. For the higher-CFR/higher-costs Africa scenario (Figure 3), vaccination is cost-effective in the medium incidence setting above a WTP threshold of ∼$1,250 per DALY averted; for the fast-waning scenario, all models find that routine vaccination at 5 years is the preferred strategy above this WTP threshold, while for the slow-waning scenario, routine vaccination at either 2 or 5 years of age is preferred. For high incidence settings in the Africa region, TCV introduction is cost-effective at WTP thresholds above $100 per DALY averted, but the preferred strategy varies depending on the waning scenario. Routine vaccination at 9 months is preferred for most WTP values in the slow-waning scenario, whereas routine vaccination at 2 years of age is mostly the preferred strategy in the fast-waning scenario. The addition of a booster dose may be cost-effective at higher WTP values, particularly for the Stanford model under the fast-waning scenario. When incidence is very high, vaccination is cost-saving, and routine vaccination at 9 months with a booster at 5 years is cost-effective at WTP thresholds above $500 for all models, regardless of the vaccine waning scenario.

**Figure 3.**
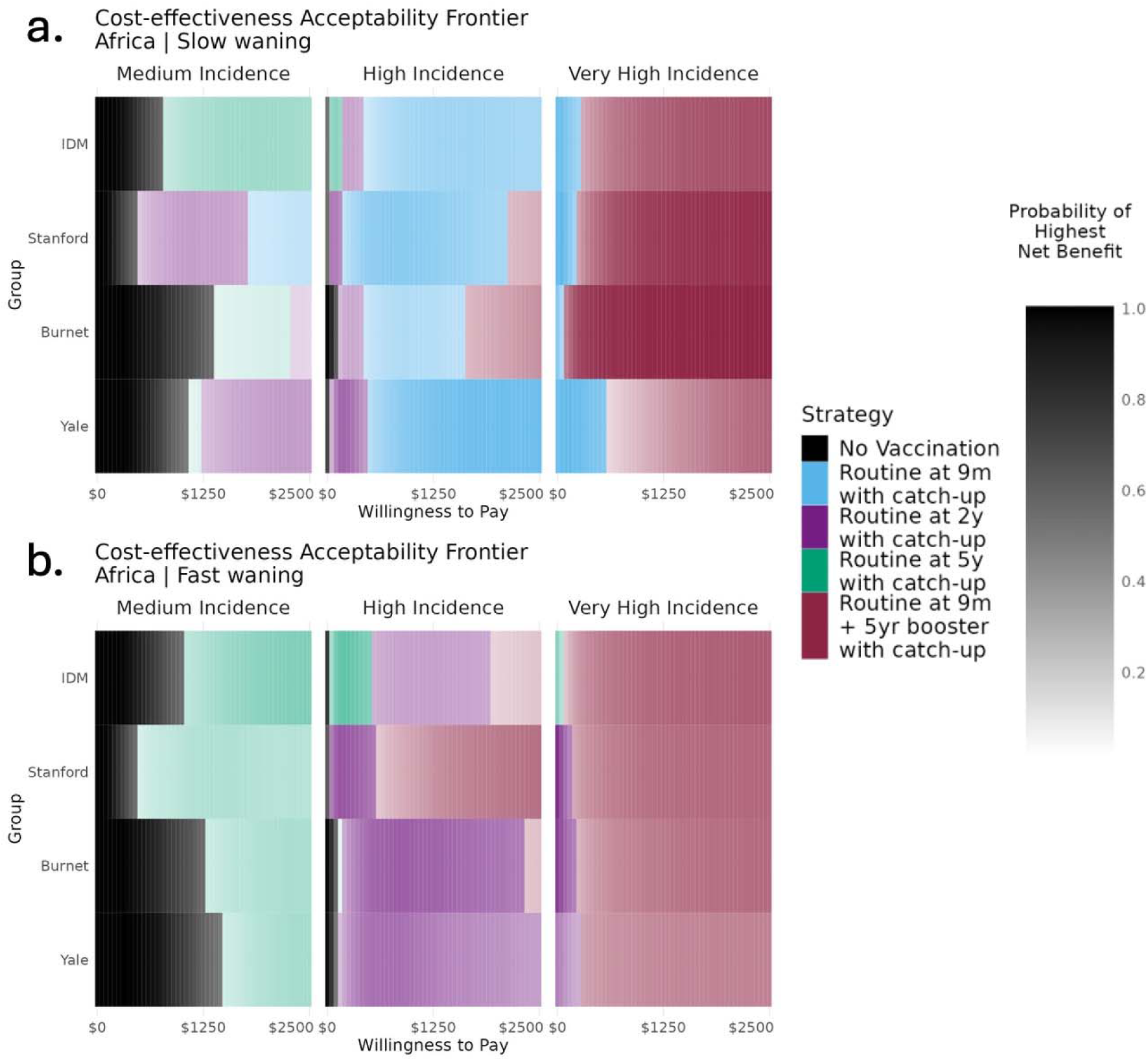
Heatmap of the cost-effectiveness acceptability frontier (CEAF) for the four models under the higher-CFR/higher-cost Africa scenario. The strategy with the greatest net benefit predicted by each model over a 10-year time horizon is indicated by the color plotted at a given willingness-to-pay threshold for (a) the slow-waning scenario and (b) the fast-waning scenario across the three incidence settings (left: medium, middle: high, right: very high). The shading indicates the probability that the strategy will provide the highest net benefit, with darker shades representing a higher probability. The willingness-to-pay threshold represents the cost per disability-adjusted life-year averted by the intervention and is given in USD up to $2,500, which is approximately equal to the average GDP of a lower-middle-income country in 2025.

In the lower-CFR/lower-costs Asia setting, vaccination is not cost-effective in medium incidence settings for WTP values below $2,500 per DALY averted (Figure 4). In high incidence settings, vaccination is cost-effective above a WTP value of ∼$700 per DALY averted, and routine vaccination at 2 years is generally preferred under both waning scenarios. Vaccination is again cost-saving in very high incidence settings, although the optimal age of vaccination differs by waning scenario. At low WTP values, the 9-month strategy is optimal when waning is slow, while routine vaccination at 2 years is preferred when waning is fast. The booster strategy was estimated to be cost-effective at WTP values above $750 (Burnet) to $4,500 (Yale) per DALY averted for the very high incidence setting when waning is slow, and at WTP values above $625 (IDM) to $1,800 (Yale) when waning is fast.

**Figure 4.**
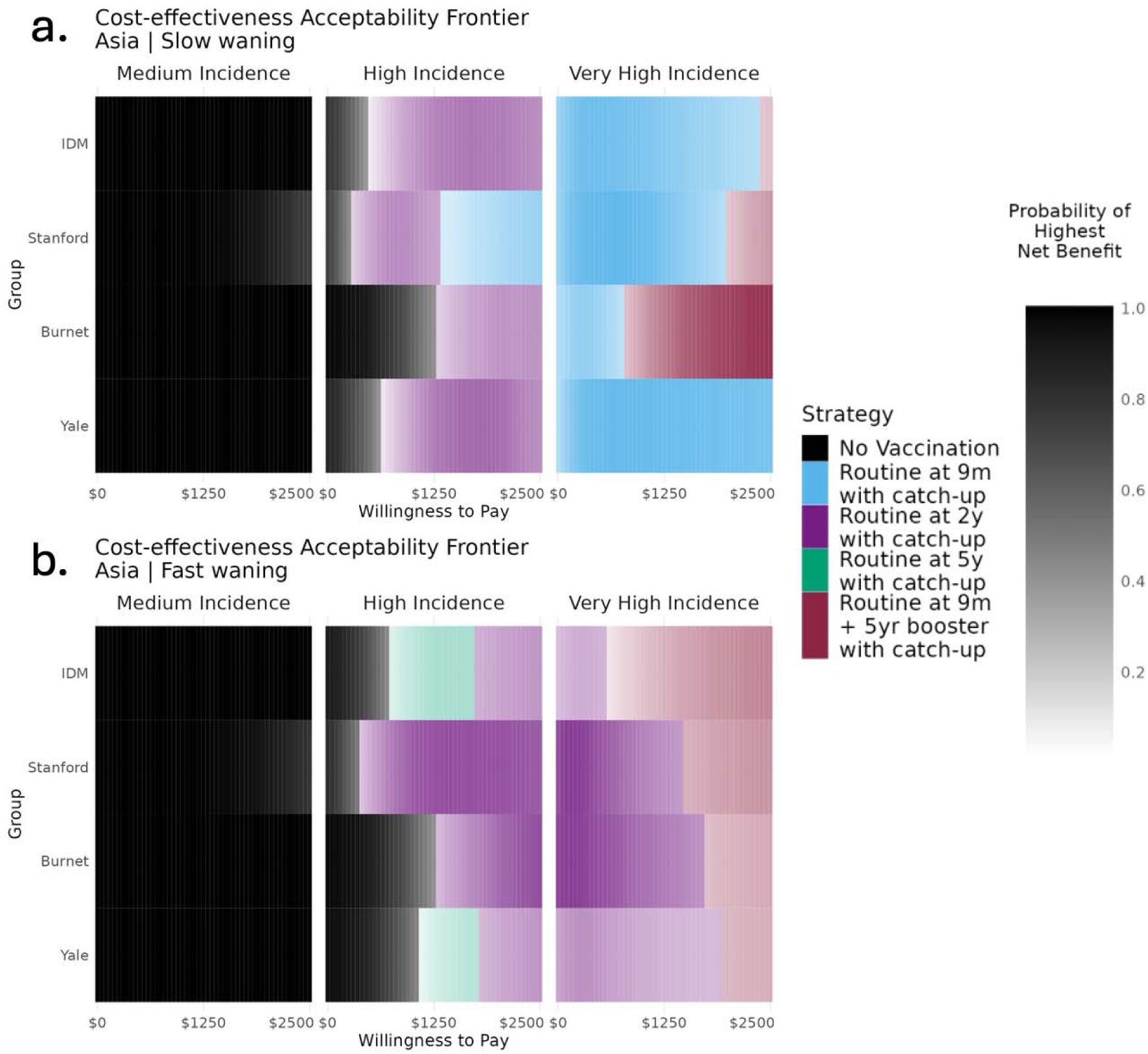
Heatmap of the cost-effectiveness acceptability frontier (CEAF) for the four models under the lower-CFR/lower-cost Asia scenario. The strategy with the greatest net benefit predicted by each model over a 10-year time horizon is indicated by the color plotted at a given willingness-to-pay threshold for (a) the slow-waning scenario and (b) the fast-waning scenario across the three incidence settings (left: medium, middle: high, right: very high). The shading indicates the probability that the strategy will provide the highest net benefit, with darker shades representing a higher probability. The willingness-to-pay threshold represents the cost per disability-adjusted life-year averted by the intervention and is given in USD up to $2,500, which is approximately equal to the average GDP of a lower-middle-income country in 2025.

When considering the cumulative health costs averted by implementing routine vaccination at 9 months with a catch-up campaign, we found that in the high and very high incidence settings, the health savings from routine vaccination at 9 months exceed the additional cost of implementing the booster dose strategy (Supplement 7.8).

### Sensitivity analyses

At a WTP threshold of $1,250 (∼0.5 times average GDP per capita for a lower-middle-income country), vaccination is cost-effective when incidence is greater than 20-60 cases per 100,000 person-years in the higher-CFR/higher-cost African setting and greater than 150-250 cases per 100,000 person-years in the lower-CFR/lower-cost Asian setting (Figure 5). As incidence and WTP increase, the optimal age of routine vaccination tends to decrease, from 2 years of age to 9 months under the slow-waning scenario and from 5 years to 2 years under the fast-waning scenario in Asia. When peak incidence is in the 10-15-year-old age group (i.e., mirroring the age distribution of medium incidence settings), vaccinating at 5 years is preferred for lower incidence levels and a larger range of WTP values (Figure S7.7.1). The addition of a booster is cost-effective at incidence levels above 100-500 cases per 100,000 person-years in Africa and above 550-1500 cases per 100,000 person-years under the fast-waning scenario in Asia across the four models (Figure 5, Figures S7.7.2-5). Extrapolating from the threshold analysis and applying the average CFR in each setting, TCV introduction is cost-effective when the typhoid mortality rate exceeds 1-4 deaths per 1 million person-years, and the addition of a booster dose is cost-effective in settings where the mortality rate exceeds 8-20 deaths per 1 million person-years across all settings.

**Figure 5.**
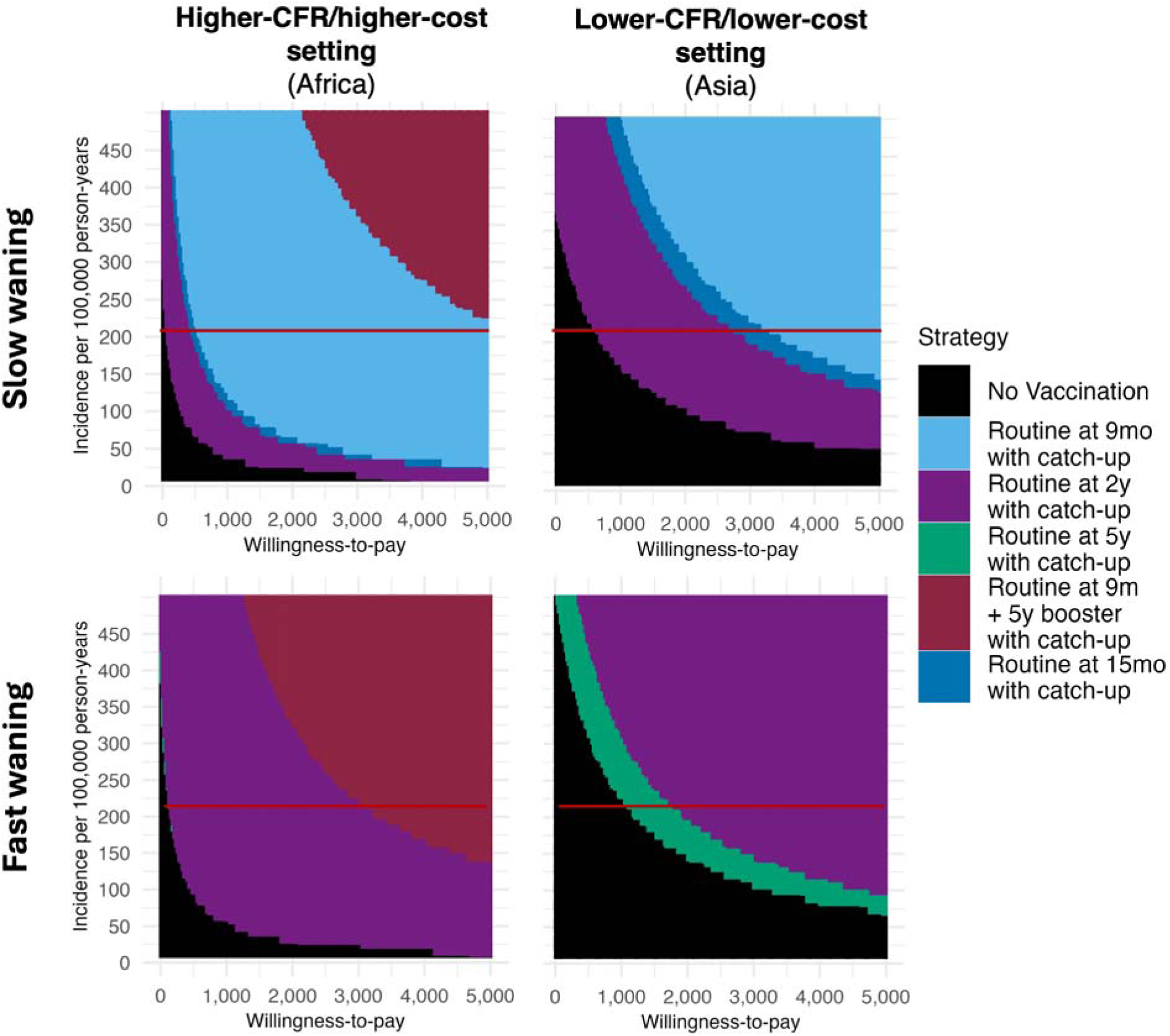
Threshold analysis of the preferred vaccination strategy across different baseline incidence values with peak incidence in the 2- to 4-year age group. The optimal strategy predicted by each model (from the cost-effectiveness acceptability frontier) is indicated by the color plotted for different baseline typhoid incidence rates (from 10 to 500 cases per 100,000 person-years) at a given willingness-to-pay per disability-adjusted life-year averted (from $0 to $5,000, x-axis). Results are plotted for a 10-year time horizon under the slow-waning scenario (top) and fast-waning scenario (bottom) from the higher-CFR/higher-cost African setting (left) and lower-CFR/lower-cost Asian setting (right). The red lines indicate the calibrated incidence for high incidence settings used in the main analysis. These plots present the results for the Yale model, which generally predicted vaccination to be cost-effective at higher WTP values than the other models; results for the other models are presented in the Supplement section 7.7.

For a 20-year time horizon, when we considered the option of adding a second booster dose at 10 years of age, we found that the two-booster strategy largely replaced the one-booster strategy as the optimal strategy in the very high incidence settings and the high incidence, higher-CFR/higher-cost African setting (Supplement 7.4). Including two boosters required about 10,000 more doses and had lower total costs compared to the one-booster strategy in the fast-waning scenario.

The most influential parameters identified by one-way sensitivity analyses differed across models, scenarios, and WTP thresholds. However, we found that the CFR, vaccine coverage, and VE waning rates were key drivers of our results across the models (Supplement 7.9).

## Discussion

Based on the predicted impact of TCV introduction from four different models, we found that TCV introduction is likely to be cost-effective in most countries with a high incidence of typhoid fever or medium incidence and higher CFR, although the optimal vaccine strategy varies based on the level of endemicity (magnitude and age distribution of incidence) of a country. Our study will provide insights into the best strategies for context-specific adoption of TCVs to inform updated WHO SAGE recommendations in light of emerging evidence on waning of vaccine protection. In medium and high incidence settings, delaying vaccination to 2 or 5 years of age may be preferred over other strategies due to longer duration of protection when TCV is administered to older children and older age of peak incidence in medium incidence settings. In very high incidence settings, where infection peaks among the youngest age groups, the addition of a booster dose at 5 years of age is likely to be cost-effective. Considering constrained health budgets and changes to Gavi funding availability, this analysis can aid decision-makers in choosing optimal strategies for TCV use.

While the four models predicted slight differences in the impact of TCV strategies on typhoid incidence, there was general agreement regarding the most cost-effective strategies according to baseline incidence and VE waning scenarios. Differences between models can be attributed to unique features influencing the results of each model. For example, the Stanford model exhibited the greatest uncertainty in vaccine impact due to the sensitivity of model projections to small changes in calibrated vaccine parameters for the distinct hybrid mode of protection assumed (see Supplement for more details). The predicted vaccine impact for the Stanford model was also substantially lower in very high incidence settings; this can be explained by a high estimated force of infection, such that individuals were first infected before being vaccine-eligible, thus reducing the benefit of vaccination. Additionally, the calibrated duration of vaccine protection for the Burnet model was considerably lower in the slow-waning scenario compared to other models. This led to lower vaccine impact and a preference for the booster strategy at lower WTP. There was also a lack of consensus on whether routine vaccination at 2 or 5 years of age is preferred in the medium-incidence, higher-CFR African setting under slow waning and in the high-incidence, lower-CFR Asian setting under both waning assumptions.

A previous multi-model comparison by Burrows et al. evaluated no vaccination, routine TCV vaccination at 9 months, and routine vaccination at 9 months with a catch-up campaign in Kolkata, India, a high incidence setting [8]. They found routine vaccination with TCV to be cost-effective and recommended implementation with a catch-up campaign. Our primary results did not consider strategies without a catch-up campaign because initial analyses revealed those strategies to have lower health benefits. Supplementary analyses considering only strategies without a catch-up campaign found vaccination to be cost-effective in some scenarios, but with much higher uncertainty (Supplement 7.6). Nevertheless, our results are consistent with the conclusions of the previous model comparison, which used similar incidence and waning conditions. Extended follow-up data from the Bangladesh trial have suggested that the duration of protection may be shorter even than the 6 years previously assumed in the pessimistic scenario, especially for individuals vaccinated before 2 years of age. Under even faster-waning assumptions, we found delaying vaccination in lower-incidence settings and adding booster doses in higher-incidence settings would be preferred at lower WTP. Our findings also parallel other results in the literature. Previous analyses also found that TCV introduction is likely to be cost-effective in settings where the typhoid incidence rate exceeds 50 to 300 cases per 100,000 person-years (depending on assumed CFR) [6–7,9,25–27], which helped to inform initial WHO SAGE recommendations for the introduction of TCVs [28]. The modeled evidence base supporting TCV introduction is growing, and our analysis provides insight into the ideal age of vaccination and the number of doses for a range of endemic settings based on recent evidence of waning vaccine-induced immunity.

Across the scenarios, the preferred strategy was influenced by the age distribution of baseline typhoid incidence, as well as the disease severity and waning parameters. Our threshold analysis showed that under fast-waning assumptions, delaying routine vaccination to 5 years of age is preferred when incidence is highest in the 5- to 15-year age group in the medium-incidence range/higher-CFR and high-incidence range/lower-CFR settings, but when incidence peaks in the 2- to 4-year age group (consistent with higher incidence settings), it is preferable to vaccinate at a younger age, potentially with a booster dose at 5 years. Under slow-waning assumptions, vaccination is preferred at younger ages because doses administered at 9 months avert cases early in life while still providing long-term protection.

Data from Pakistan suggests that waning of VE and associated rebounds in typhoid incidence may have occurred faster in Sindh versus Punjab province following the introduction of routine vaccination at 9 months with a catch-up campaign because of a higher baseline typhoid incidence in Sindh [29]. If faster waning of TCV-induced immunity is associated with a higher force of infection, then this reinforces our findings that the introduction of a booster dose is likely to be cost-effective in very high incidence settings. Given the influence of age-specific peak incidence and waning on the preferred strategy, countries with knowledge of the waning dynamics may consider adding a booster dose to combat a rebound in cases. If investment in additional doses is infeasible, tailoring the age of vaccination to the age of peak incidence presents an alternative option.

Differences in disease severity and treatment costs parameters sourced from studies conducted in African versus Asian settings also influenced the preferred vaccination strategy, driven primarily by differences in the CFR according to the one-way sensitivity analyses. This is likely due to delays in care-seeking and diagnosis of typhoid in the African studies, rather than representative of regional differences per se. When the CFR is less than 0.4%, as we assumed for the Asian scenario, vaccination was preferred only in higher incidence settings and at higher WTP values. Conversely, because the African scenario had high treatment costs and a higher CFR, there were greater health savings and DALYs averted on a per-case basis, making vaccination more cost-effective.

Despite agreement amongst the models, there was no one TCV strategy that was optimal across scenarios and settings, requiring nuanced schedule recommendations. Determining the optimal strategy for a specific country is beyond the scope of this study. Typhoid surveillance is challenging due to the non-specific symptoms and reliance on blood culture for definitive diagnosis, which prevents many countries from having a reliable understanding of typhoid incidence, age distribution, and mortality rates, making it difficult to identify the best TCV schedule. Furthermore, it is unclear what dictates the durability of vaccine protection in different settings, although it is hypothesized that faster waning may be associated with a higher force of infection. By monitoring cases after vaccine introduction, decision-makers can evaluate whether a rebound in typhoid is occurring and introduce booster doses as needed.

From an implementation perspective, there are concerns about the cost of introducing booster doses and the feasibility of implementing vaccination outside of the routine Essential Programme on Immunization (EPI) schedule. However, we found that in settings where a booster dose is recommended, health savings from implementing TCVs exceeded the costs of booster introduction after a few years, which in turn would lead to longer-term health savings. Furthermore, we did not consider Gavi support, so for countries receiving cost-sharing assistance, highly beneficial but more costly strategies would become cost-effective at lower WTP values. We also identified strategies that align with other high-priority vaccine schedules, including the fourth dose of malaria vaccine at 2 years of age and campaign or school-based vaccination (e.g., against human papillomavirus) for older ages [30–32]. Finally, there are equity concerns regarding children who would be missed by school-based vaccination, so programmatic efforts to reach zero-dose children should include vaccination with TCV as well [33].

This study has multiple limitations. All models made a number of simplifying assumptions related to natural history, transmission, health outcomes, and costing. While chronic carriage plays a role in the modeled impact of TCVs, we made assumptions about the duration and relative infectiousness of carriage, despite these parameters not being well studied. Furthermore, the models were calibrated to reproduce the weak indirect protection from TCVs observed in a cluster RCT conducted in Bangladesh, but indirect protection may vary for other settings [11,13]. The incidence data used to fit age-specific incidence rates only included studies that reported cases from all age groups, thus potentially excluding studies with inadequate surveillance but substantial unreported burden. We only included vaccine coverage data for countries that had estimates for all four types of coverage, which may overrepresent countries with strong EPI programs.

While we aimed to explicitly account for the higher treatment costs and poorer outcomes associated with AMR in our cost-effectiveness analysis, we were unable to find the data needed to reliably identify the probability of hospitalization and costs of treatment for resistant and sensitive cases. As a result, we sought parameters for populations consisting of both types of cases from settings with known resistance.

Furthermore, the emergence of resistance to last-line drugs may be associated with higher treatment costs and mortality (although cost-of-illness estimates are unavailable), making our findings potentially conservative in terms of cost-effectiveness for settings where such resistance exists [33]. We were also unable to identify treatment-specific CFRs due to minimal data available on the CFR for outpatient and untreated cases of typhoid fever. We assumed the overall CFR accounted for all treatment types and chose conservative estimates based on more recent studies to not overinflate the benefit of vaccination. Similarly, our cost-effectiveness parameters were taken from country-level studies and generalized to the region, which may not be a representative average, nor does it account for regional heterogeneity. Finally, we did not include start-up costs for the implementation of new TCV strategies, which may increase the WTP threshold at which vaccination is cost-effective.

Local surveillance systems and monitoring are imperative for parameterizing models, evaluating real-world vaccine impact, characterizing local burden, and making decisions on the optimal TCV strategy in the local and global contexts [33]. In addition to incidence, more data is needed to quantify the relative costs and impact of AMR cases and their risk of hospitalization and complications [33,34]. Vaccination alone will not eliminate typhoid, so TCV introduction should be considered in conjunction with sanitation efforts and plans for antimicrobial stewardship. Future analyses could evaluate the cost-effectiveness of implementing supplementary water, sanitation, and hygiene interventions and would benefit from more specific data on typhoid outcomes by treatment type and setting [34].

This multi-model comparison of TCV impact and cost-effectiveness provides a comprehensive analysis of the optimal vaccine schedules across diverse settings in support of updated global recommendations from WHO. We also provide evidence to help inform country-level decision-making based on disease burden, vaccine waning, vaccine delivery and treatment costs, and willingness to pay for health interventions. Importantly, we demonstrate that TCV introduction remains a critical component of efforts to combat the considerable burden of typhoid fever in LMICs, despite recent evidence on the waning of vaccine protection. Delaying the age of vaccination or adding booster doses could enhance population-level vaccine impact in typhoid-endemic regions.

## Supporting information

Supplemental Table - Model Characteristics

CHEERS Checklist

## Data Availability

All data produced in the present work are contained in the manuscript or available online.

https://zenodo.org/records/18687859

https://github.com/starsimhub/typhoidsim

https://github.com/goshgondar2018/TCV_booster_modeling

https://doi.org/10.5281/zenodo.18837535

