## Supplemental Text for "Evaluating the Impact and Cost-Effectiveness of Typhoid Conjugate Vaccine Schedules Across Diverse Settings: A Multi-Model Comparison"

^†^co-last authors

Disclaimer: RH and VM are staff members of the World Health Organization. The views expressed are their personal ones and do not necessarily represent the views of their organization. KHG, AK, and JG are employees of the Gates Foundation; however, this study does not necessarily represent the views of the Gates Foundation.

### 1 Demographic data

We harmonized the four epidemiological models by assuming the same population dynamics based on the demographic parameters of low- and lower-middle-income countries (LMICs) from the United Nations World Population Prospects [1]. We used crude birth rates and the proportion of the population by age group to fit age-specific all-cause death rates that result in a stable population size and age distribution over time. The crude birthrate was 22.8 births per 1000 persons per year, and the life expectancy was 68.88 years. The age composition of the population is detailed in Table S1.1.

**Table S1.1: Proportion of the population in each age group.**

| **Age group** | **Proportion of the population** |
| --- | --- |
| 0-<2y | 0.044 |
| 2-<5y | 0.064 |
| 5-<10y | 0.105 |
| 10-<15y | 0.102 |
| 15-<20y | 0.096 |
| 20-<25y | 0.089 |
| 25+y | 0.502 |

### 2 Incidence Data

To define the average incidence and age distribution of cases for each incidence archetype, we conducted a literature search to identify studies that conducted population-based or hybrid surveillance of typhoid fever. We did not include studies from low incidence settings (<10 cases per 100,000 person-years) as these are typically from high- or upper-middle-income countries in which typhoid is not endemic. Only studies that reported incidence rates for all age categories (0-<5, 5-<15, and 15+ years old) were included. We extracted data on blood-culture-confirmed case counts and person-time of follow-up, which were adjusted for the probability of seeking care, the proportion of cases meeting the case definition with blood drawn for culture, and blood culture sensitivity, as reported by the studies. The data extracted for each country was categorized into medium incidence (10-99 cases per 100,000 person-years), high incidence (100-499 cases per 100,000 person-years), and very high incidence (500+cases per 100,000 person-years) based on the adjusted overall incidence. We ultimately included 36 studies from 18 countries (Table S2.2).

To achieve a finer breakdown of age groups, we used a subset of studies that reported cases and person-time separately for the 0-<2-year, 2-<5-year, 5-<10-year, and 10-<15-year age categories (but may not have reported all age groups) to calculate the ratio of incidence in the 0-<2-year age group compared to the 2-<5-year age group and the 5-<10-year age group compared to the 10-<15-year age group for each incidence setting. These ratios were then used to redistribute the cases in the 0-<5-year and 5-<15-year age groups of the complete data set into the finer age categories. From there, the person-time was divided based on the proportion of the population in each of the smaller age groups. We added up the cases and person-time in each age group across all of the studies in each of the three incidence settings, and age-specific incidence was calculated for the new age groups.

**Table S2.1: Age distribution of incidence in each incidence archetype.**

| **Age groups** | **Cases** | **Person-years (PY)** | **Incidence per 100K PY** |
| --- | --- | --- | --- |
| ***Medium incidence settings*** | | | |
| 0-<2y | 3 | 13801 | 22 |
| 2-<5y | 10 | 20074 | 50 |
| 5-<10y | 30 | 45891 | 66 |
| 10-<15y | 32 | 44580 | 71 |
| 15+y | 18 | 52249 | 34 |
| All | 93 | 176595 | 53 |
| ***High incidence settings*** | | | |
| 0-<2y | 39 | 24601 | 157 |
| 2-<5y | 185 | 35784 | 518 |
| 5-<10y | 275 | 71742 | 383 |
| 10-<15y | 235 | 69692 | 337 |
| 15+y | 450 | 350883 | 128 |
| All | 1184 | 552703 | 214 |
| ***Very high incidence settings*** | | | |
| 0-<2y | 53 | 3976 | 1343 |
| 2-<5y | 241 | 5784 | 4159 |
| 5-<10y | 359 | 13019 | 2757 |
| 10-<15y | 239 | 12647 | 1890 |
| 15+y | 367 | 64928 | 565 |
| All | 1259 | 100355 | 1255 |

**Figure S2.1: Age distribution of incidence in each incidence archetype.** Incidence rates are adjusted for the probability of seeking care for febrile illness, the fraction of cases meeting the case definition for which blood was drawn, and the sensitivity of blood culture for typhoid diagnosis.

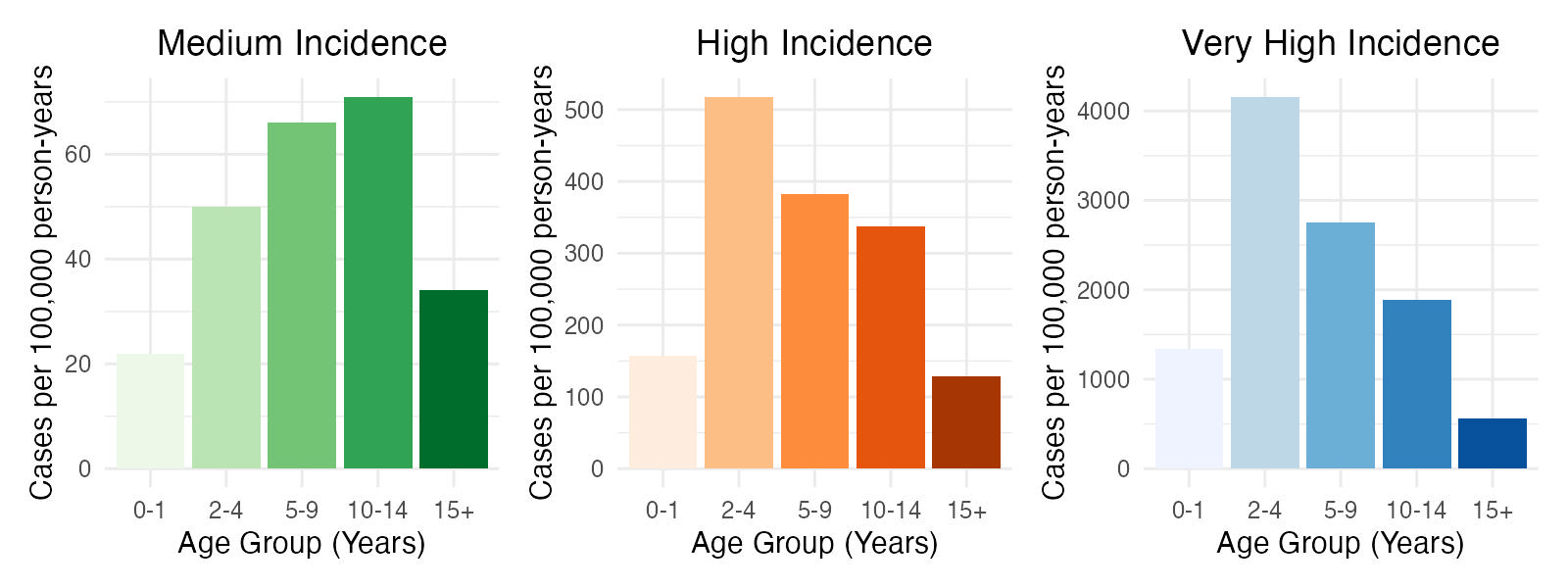

**Table S2.2: Studies used to inform the age-specific incidence in archetypal settings.** Location, median year, and source of data is noted. Studies are categorized by the overall adjusted incidence of typhoid fever (medium: 10-99 cases per 100,000 person-years; high: 100-499 cases per 100,000 person-years; very high: 500+ cases per 100,000 person-years).

| **Location** | **Year of data collection** | **Source** |
| --- | --- | --- |
| ***Medium incidence*** | | |
| Bandim, Guinea-Bissau | 2012 | [2] |
| Butajira, Ethiopia | 2013 | [2] |
| Ibadan, Nigeria | 2018 | [3] |
| Isotry, Madagascar | 2012 | [2] |
| Moshi, Tanzania (rural) | 2013 | [2] |
| Imerinsiatosika, Madagascar | 2012 | [2] |
| Pikine, Senegal | 2012 | [2] |
| ***High incidence*** | | |
| Blantyre, Malawi | 2016 | [4] |
| Agogo, Ghana | 2017 | [3] |
| Dhulikhel, Nepal | 2018 | [5] |
| East Champaran, India | 2019 | [6] |
| Nioko, Burkina Faso | 2012 | [1] |
| Karachi, Pakistan | 2018 | [5] |
| Karimganj, India | 2019 | [6] |
| Kathmandu, Nepal | 2018 | [5] |
| Kavuaya and Nkandu, Democratic Republic of the Congo | 2019 | [3] |
| Kolkata, India | 2004 | [7] |
| Kullu, India | 2019 | [6] |
| Lwak, Kenya | 2008 | [8] |
| Moshi, Tanzania (urban) | 2013 | [2] |
| Nandurbar, India | 2019 | [6] |
| Kibera, Kenya | 2013 | [2] |
| Polesgo, Burkina Faso | 2012 | [2] |
| North Jakarta, Indonesia | 2002 | [9] |
| Pemba, Tanzania | 2009 | [10] |
| Samarkand City, Uzbekestan | 2002 | [11] |
| ***Very high incidence*** | | |
| Anantpur, India | 2019 | [6] |
| Chandigarh, India | 2019 | [6] |
| Communes Dong Thap, Vietnam | 1996 | [12] |
| Delhi, India | 1996 | [13] |
| Dhaka, Bangladesh | 2001 | [14] |
| Dhaka, Bangladesh | 2016 | [4] |
| Kathmandu, Nepal | 2016 | [4] |
| Kibera, Kenya | 2008 | [8] |
| Kolkata, India | 2005 | [7] |
| Sumatra, Indonesia | 1988 | [15] |

### 3 Vaccine Effectiveness and Waning

We considered two different scenarios for the waning of vaccine effectiveness (VE) over time based on observations from randomized controlled trials (RCTs). The differences in the waning of VE between RCTs conducted in Malawi and Bangladesh may be due to differences in the local disease dynamics—potentially transmission intensity—although the true cause is unknown. Therefore, we model both fast- and slow-waning scenarios to understand how the preferred vaccination strategy may change based on waning assumptions.

The “slow-waning” scenario is based on data from the TyVAC-Malawi RCT conducted in Blantyre, Malawi [16,17]. Trial participants 9 months to 12 years of age were randomized to receive either the Tybar-TCV typhoid conjugate vaccine (Bharat Biotech) or a control vaccine (meningococcal A conjugate vaccine) and were followed for up to 4.61 years after vaccination [16]. The overall VE for all age groups in the intention-to-treat analysis declined from 80.7% (95% confidence interval (CI): 62.4%, 89.6%) after 2 years of follow-up [16] to 78.3% (95% CI: 66.3%, 86.1%) after 4-5 years of follow-up [17], suggesting minimal waning of immunity among study participants. Data was available to stratify participants into two age groups based on age at vaccination: 9 months to <5 years and 5 to <15 years of age [16,17].

The “fast-waning” scenario is based on data from the TyVAC-Bangladesh cluster RCT and TyVOID study conducted in Dhaka, Bangladesh [18,19]. Beginning in April 2018, the TyVAC-Bangladesh trial randomized participants 9 months to <16 years of age living in 150 geographic clusters to either receive the Tybar-TCV (Bharat Biotech) or a control vaccine (SA 14-14-2 against Japanese encephalitis) based on their cluster of residence. Participants were followed for 17.1 months on average until December 2019. Total vaccine effectiveness for all age in the TyVAC-Bangladesh trial was 85% (97.5% CI: 76%, 91%) [18]. Data was available to stratify participants into three age groups based on age at vaccination: 9 months to <2 years, 2 years to <5 years, and 5 to 15 years of age [18-19].

Following the trial (and a hiatus in surveillance due to the COVID-19 pandemic), participants who had received the control vaccine were offered the Tybar-TCV vaccine. The TyVOID study then followed the recent TCV recipients (vaccinated in 2021) and compared them to previous TCV recipients (vaccinated in 2018-19) to estimate relative vaccine efficacy as well as conducted a test-negative case-control study comparing the vaccination status of those who test blood-culture-positive for *Salmonella* Typhi to those who tested negative or positive for another organism [19]. The VE for all age groups in the previous-TCV group 3 to 5 years after vaccination was 55% (95% CI: 36%, 68%) based on the test-negative design, which was similar to the VE of 50% (95% CI: -13%, 78%) extrapolated from the relative efficacy among the previous versus recent TCV recipients [19]. Follow-up has continued, and VE has declined further based on unpublished data through May 2025. The VE estimates by age group and duration of follow-up are available in Table S3.1.

**Table S3.1: Vaccine efficacy and effectiveness estimates by age group and duration of follow-up for the TyVAC-Malawi, TyVAC-Bangladesh and TyVOID-Bangladhesh studies.**

| ***Malawi*** | | Follow-up period | | | |
| --- | --- | --- | --- | --- | --- |
| Midpoint of follow-up | | 1 year | | 2.305 years | |
| Age group | <5 years | 74.4% (31.7%, 90.4%) | | 70.6% (6.4%, 93%) | |
|  | 5-15 years | 83.7% (63.6%, 92.7%) | | 79.3% (63.5%, 89.0%) | |
| ***Bangladesh*** | | Follow-up period | | | |
| Midpoint of follow-up | | 1 year | 4 years | | 6 years |
| Age group | <2 years | 81% (39%, 94%) | 24% (-29%, 55%) | | -5% (-118%, 49%) |
|  | 2-<5 years | 80% (62%, 89%) | 59% (12%, 81%) | | 14% (-196%, 75%) |
|  | 5-15 years | 88% (78%, 93%) | 74% (41%, 89%) | | 32% (-63%, 71%) |

To estimate age-specific initial vaccine efficacy and mean duration of protection from the VE estimates in Table S3.1, we used Bayesian inference implemented via Markov Chain Monte Carlo using the JAGS (Just Another Gibbs Sampler) software. The parameters were jointly estimated using an observational model and a time-varying protection model using uninformative prior distributions.

For both waning scenarios, we evaluated different models assuming vaccine protection follows an exponential distribution, a gamma distribution with uninformative shape and scale parameters, and gamma distribution with fixed shape parameters at $s$ = 2, 3, 5, 7, and 10; note that the exponential distribution is equivalent to a gamma distribution with a shape parameter of $s=1$. We compared the models based on Akaike information criteria (AIC), Bayesian information criteria (BIC), and deviance information criteria (DIC) and chose the most parsimonious fit.

For the slow-waning scenario, the expected VE for those vaccinated in age group *a* at time *t* post-vaccination is described by the exponential waning function:

$${VE}_{a}\left( t \right)={VE}_{0,a}exp({-\omega}_{v,a}t)$$

where 𝑉𝐸_0,𝑎_ is the initial vaccine efficacy, which represents the reduction in risk of infection for a TCV recipient shortly after vaccination (before any waning has occurred), and the duration of TCV protection is exponentially distributed with mean 1/𝜔_𝑣,𝑎_, where 𝜔_𝑣,𝑎_ is the rate of waning of TCV-induced immunity in age group *a*.

For the fast-waning scenario, the expected VE for those vaccinated in age group *a* at time *t* post-vaccination is described by the function:

$${VE}_{a}\left( t \right)={VE}_{0,a}\times\left[ 1-F_{\Gamma}(t,s,\omega_{v,a}) \right]$$

where 𝑉𝐸_0,𝑎_ is again the initial vaccine efficacy, and the duration of TCV protection follows a gamma distribution with a fixed shape parameter $s$, where 𝜔_𝑣,𝑎_ is the rate of waning of TCV-induced immunity in age group *a*. The overall best performing model was identified to be the model with shape $s=2$.

We used an observational model to account for uncertainty in the VE estimates, given that the trials were conducted in a limited sample of the full population. We defined the likelihood of the observed values of 𝑉𝐸_𝑎_(𝑡) from Table S3.1, given the “true” level of protection and the precision of the trial measurements. Since relative risk (=1-VE) follows an approximately normal distribution once it is log-transformed, we modeled:

$$\log\left( 1-\hat{VE}\left( t \right) \right)\sim Normal(log(1-VE\left( t \right),\sigma^{2})$$

where the variance, 𝜎^2^, can be approximated from the confidence interval of the observed VE, $\hat{VE}\left( t \right)$.

The fitted models are shown in Figure S3.1. Two models (IDM and Yale) used these estimates directly in the simulations, while two models (Burnet and Stanford) directly fit their models to the clinical trial data. See supplement sections 5.2 and 5.4 for a description of the fitting approaches.

**Table S3.2: Estimated vaccine effectiveness and waning parameters for the transmission models.** Values are presented as the mean estimates and 95% prediction intervals.

| Average duration of protection (years) | | | |
| --- | --- | --- | --- |
| Group | **Burnet** | **Stanford** | **IDM/Yale** |
| *Fast waning* | | | |
| <2y | 1.82 | 1.1 (1.07-1.17) | 2.8 (0.01-6.6) |
| 2 to <5y | 4.2 | 7.0 (2.7-18.5) | 7.3 (2.6-34.0) |
| >5y | 5.1 | 7.1 (5.2-10.6) | 10.7(4.2-42.3) |
| *Slow waning* | | | |
| <5y | 7.2 | 73.9 (41.2-83.3) | 46.4 (3.7-97.3) |
| >5y | 7.2 | 36.9 (21.8-81.4) | 52.4 (10.4-97.6) |
| VE0 | | | |
| Group | **Burnet** | **Stanford** | **IDM/Yale** |
| *Fast waning* | | | |
| <2y | 85% | 99%  (99%,100%) | 85.8%  (85.3%, 86.3%) |
| 2 to <5y | 78% | 82.8%  (71.2%, 97.2%) | 82.7%  (82.3%, 83.1%) |
| >5y | 80% | 95.6%  (92.8%, 97.6%) | 89.3%  (89.0%, 89.6%) |
| *Slow waning* | | | |
| <5y | 75% | 89.0%  (86.8%, 90.7%) | 80.2%  (79.7%, 80.6%) |
| >5y | 75% | 93.6%  (89.4%, 95.8%) | 84.1%  (83.8%, 84.3%) |

**Figure S3.1: Modeled statistical estimates of waning of vaccine effectiveness over time by age group**. The lines represent the modeled vaccine protection, while the shaded regions correspond to the 95% confidence intervals for the slow-waning scenario (blue) and fast-waning scenario (red). The points with vertical lines are the observed vaccine effectiveness estimates and 95% confidence intervals from the RCTs.

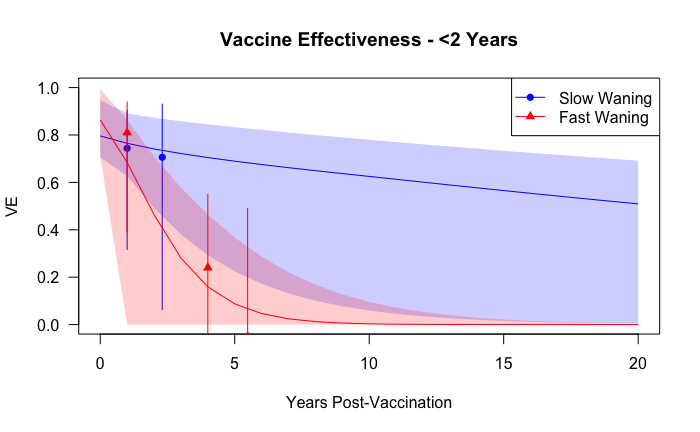

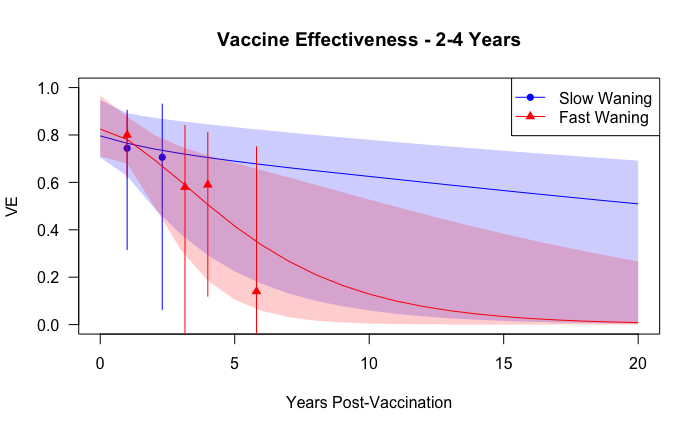

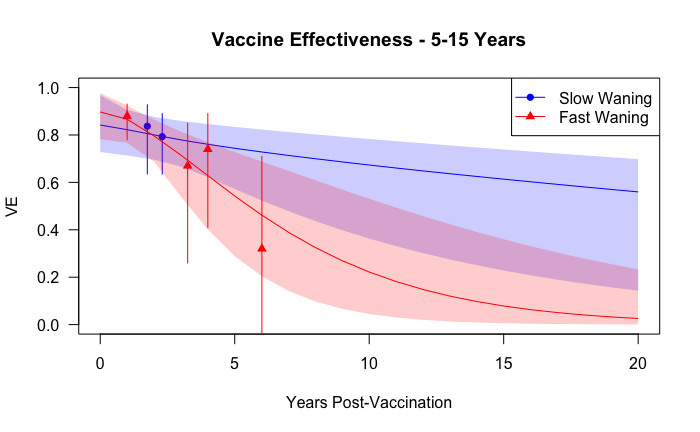

c.

b.

a.

### 4 Vaccine Coverage

The WHO has limited data on TCV coverage (<10) for countries within the AFR and WPR regions [20]. Instead, we used coverage estimates for more widely used vaccines that are administered at timepoints of interest for potential typhoid vaccination schedules. For routine vaccination at 9 months and 15 months, we used WHO/UNICEF Estimates of National Immunization Coverage (WUENIC) estimates for measles-containing vaccine first and second dose (MCV1 and MCV2), respectively, for low- and lower-middle-income countries [21]. The MCV1 estimates were similar to coverage estimates for TCVs in countries that introduced routine immunization [20,21]. MCV2 estimates were multiplied by a fraction (randomly sampled uniformly from 50-100%) to estimate coverage for routine vaccination at 2 years, which assumes it is harder to reach a child for vaccination at 2 years. For routine vaccination at 5 years and booster doses at 5 and 10 years, we assumed coverage to be some fraction (randomly sampled uniformly from 50-100%) of UNICEF school attendance estimates [22]. Finally, catch-up campaign coverage was based on unpublished estimates of MCV supplementary immunization activity (SIA) coverage (unpublished data).

Coverage estimates were matched by country, sampled with replacement, and used for each simulation to preserve correlation between coverage types while allowing for parameter uncertainty. We only included countries that fell within the WHO regions of interest (Africa, Asia, Western Pacific) and had coverage estimates for all vaccine schedules.

**Table S4.1: Mean and 95% confidence intervals of vaccine coverage estimates.**

| Vaccine Schedule | Mean Coverage (95% CI) |
| --- | --- |
| Supplementary Immunization Activities | 86.8% (82.8%, 90.9%) |
| MCV1 | 78.4% (74.0%, 82.8%) |
| MCV2 | 64.3% (57.9%, 70.6%) |
| School attendance | 80.5% (76.4%, 84.6%) |

**Figure S4.1: Distribution of vaccine coverage estimates**

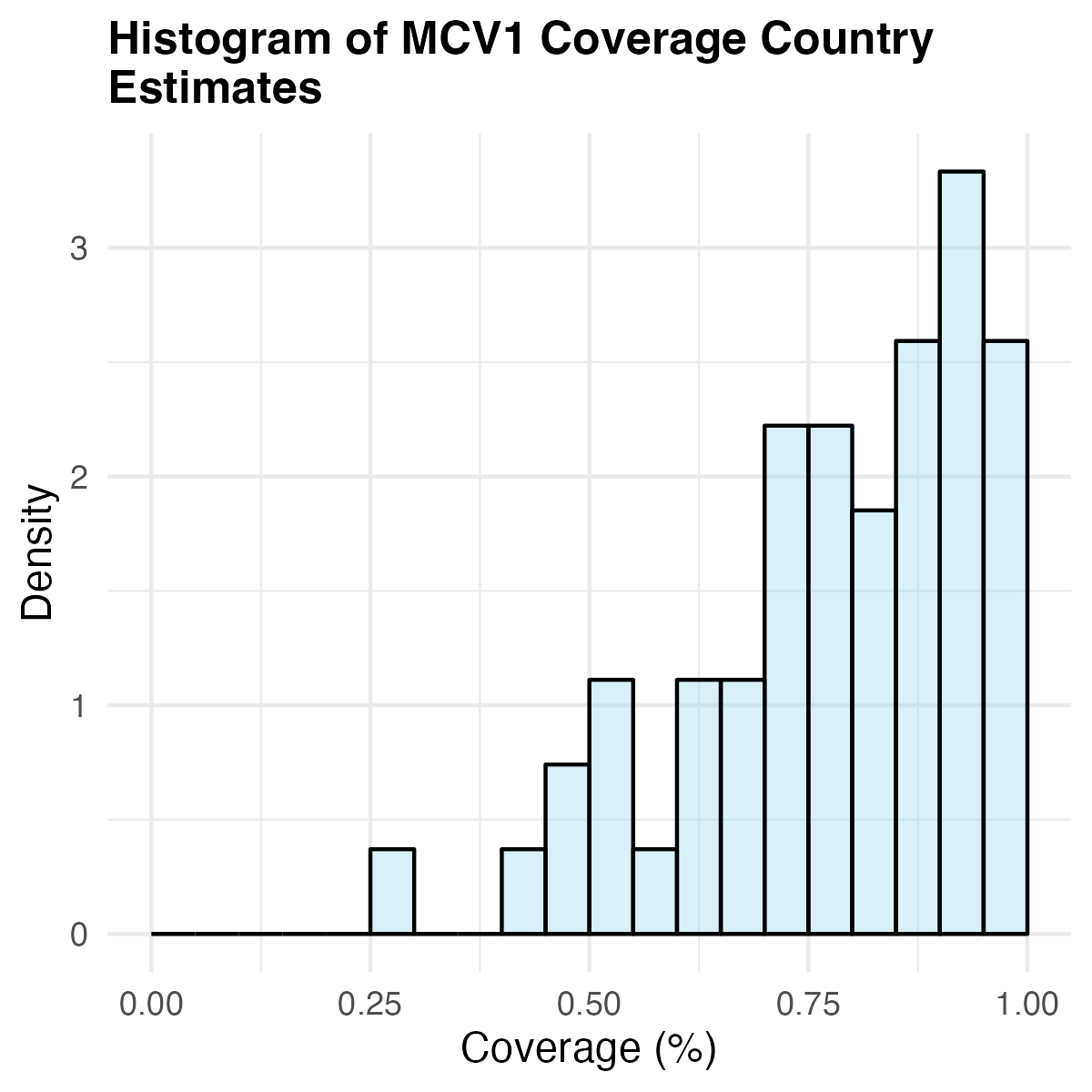

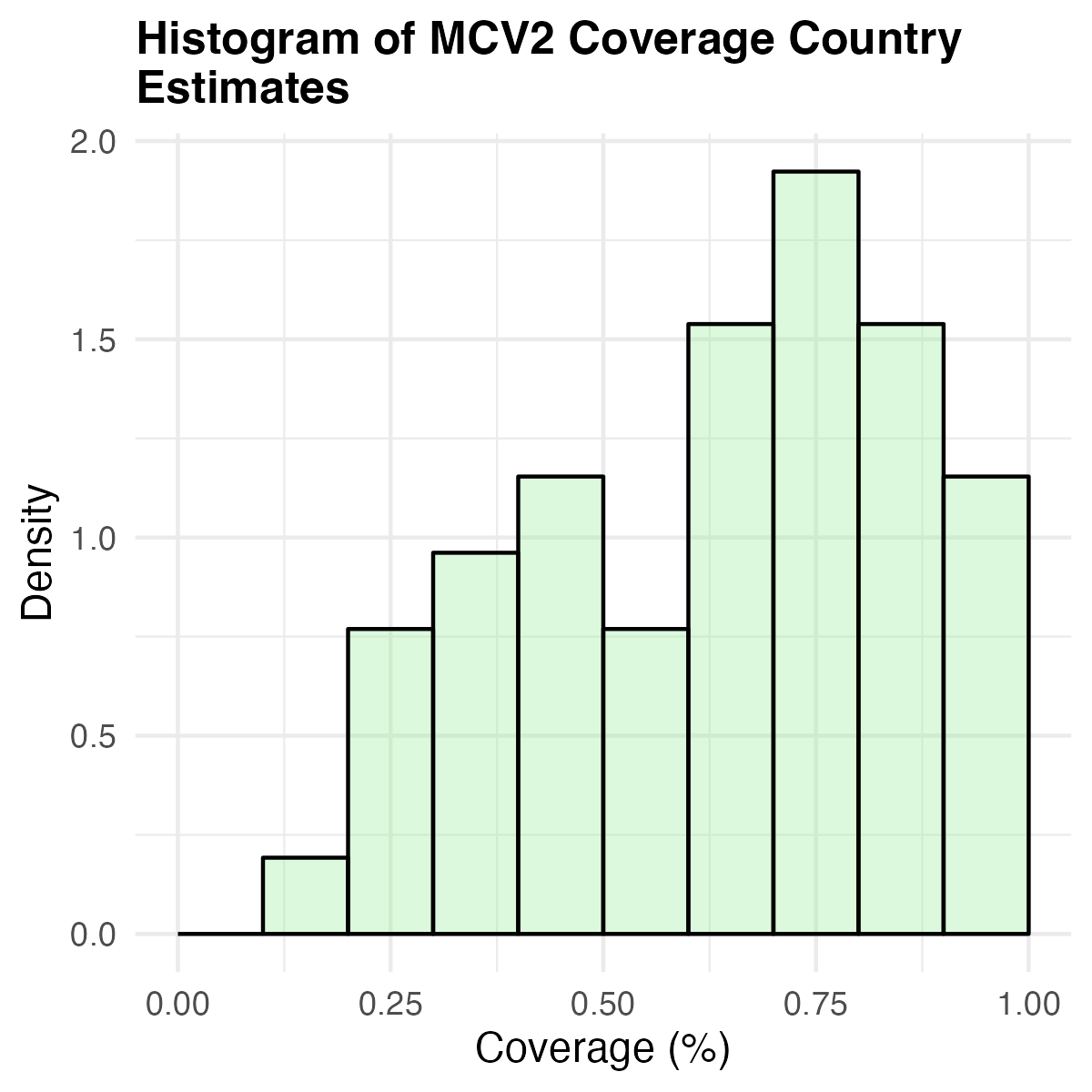

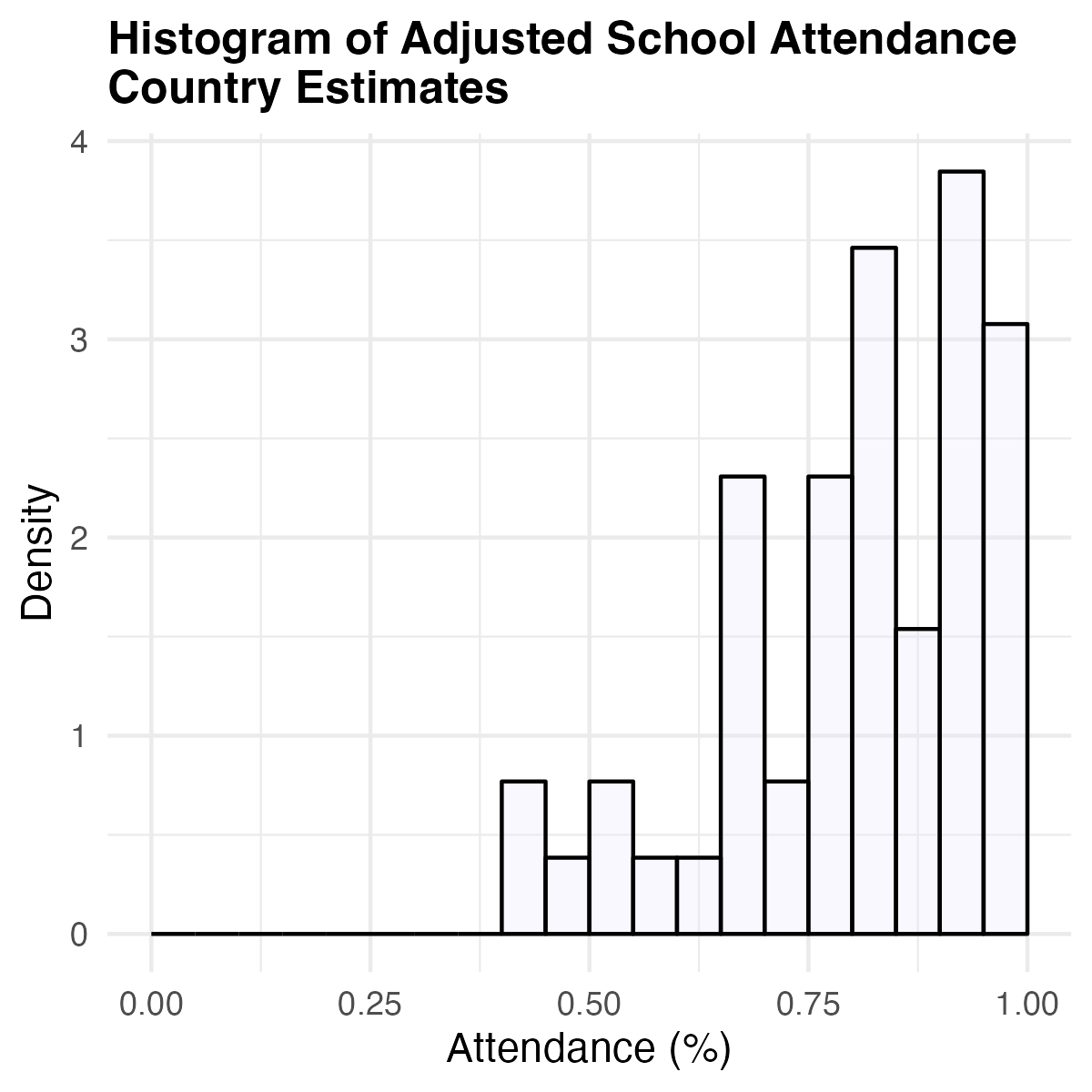

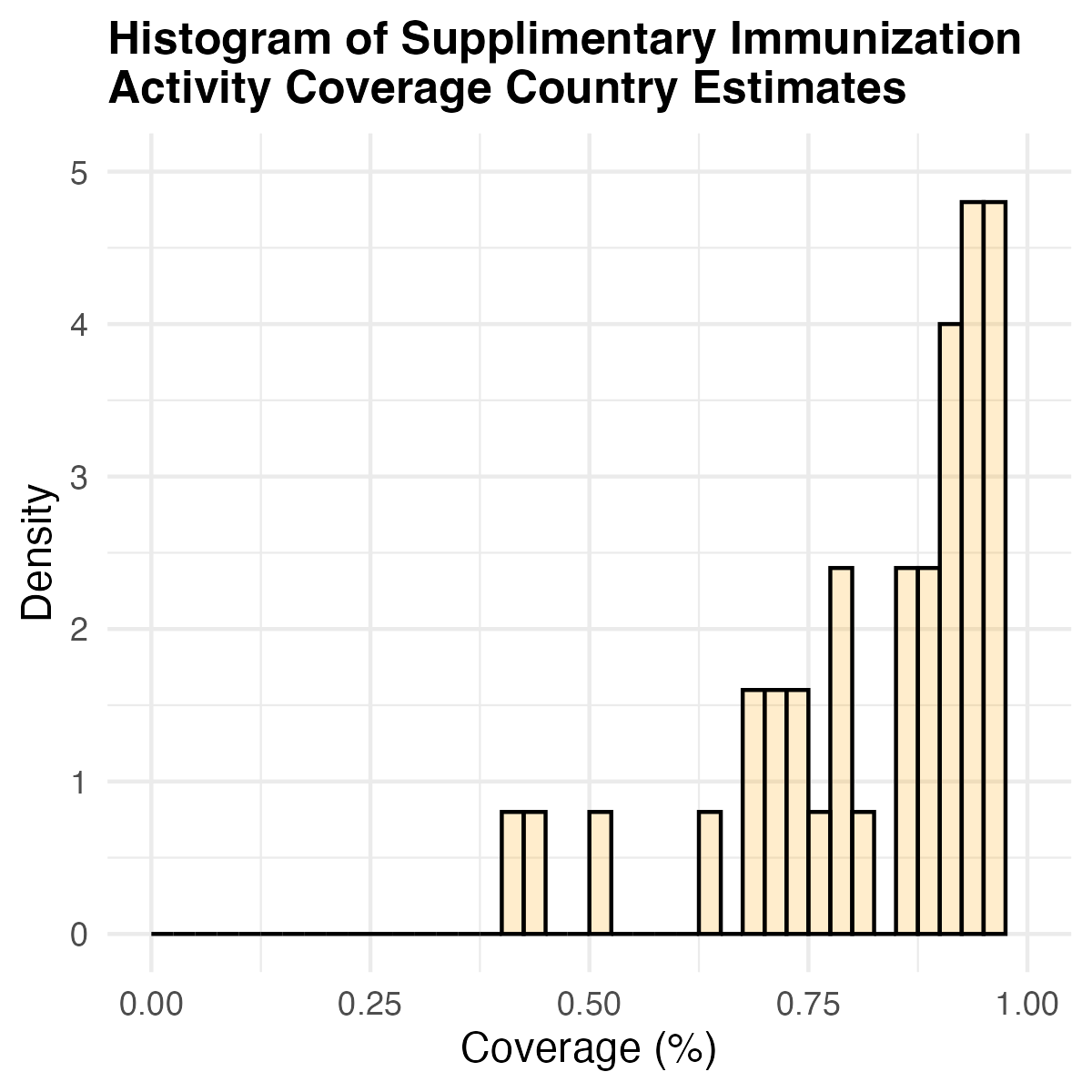

### 5 Model Descriptions

#### 5.1 Model harmonization

For this multi-model comparison, four modeling groups were selected in response to a “Request for Proposals for Modelled Evidence To Support A SAGE WG On Typhoid Conjugate Vaccine (TCV)” [23]; the teams were from the Burnet Institute, the Bill & Melinda Gates Foundation Institute for Disease Modeling (IDM), Stanford University, and Yale University. All four models are age-stratified and simulate typhoid transmission dynamics in a population; two are compartmental (Stanford and Yale) and two are agent-based (Burnet and IDM). We harmonized the models around key features of typhoid natural history while preserving unique model elements where there may be uncertainty about typhoid dynamics. In all models, susceptible individuals become infected based on the force of infection of typhoid in the population at that time step, which is proportional to the prevalence of infectious individuals. Following a period of symptomatic or asymptomatic infection, individuals may recover and have natural immunity or become chronic carriers, asymptotically contributing to the force of infection. Natural immunity is modeled as either a recovered class whose protection wanes until they enter the fully susceptible class or a partially susceptible class with a certain level of protection based on previous infections. We model clinical and subclinical infection based on prior infection history, where individuals may or may not experience symptoms, but both groups contribute to the force of infection.

All models assume individuals can also gain immunity via vaccination. The Yale, IDM, and Burnet models assume fully “leaky” vaccine-induced immunity in which individuals enter the vaccinated class based on vaccine coverage. Upon exposure to typhoid, a fraction of those individuals, equal to the initial vaccine efficacy (*VE*_0,_*_a_*) in age group *a* times the number of vaccinated individuals in age group *a*, remain protected, while the complement can become infected and move to the infected state. The Stanford model assumed a hybrid leaky/all-or-nothing vaccine-induced immunity mechanism, as described in section 5.4. For all models, vaccine-derived immunity is assumed to wane at a constant rate over time (*ω_v,a_*); when protection has fully waned, individuals re-enter a susceptible state.

We first calibrated the models to the harmonized demographic data by fitting the age-specific background mortality, establishing similar, stable population dynamics in each model. Then, each model was fitted to the age-specific incidence data from the three incidence settings. This was achieved through estimating the model-specific transmission parameters (see below). The models also simulate vaccine efficacy and waning of vaccine protection as described in section 3. We assumed booster doses are administered only to individuals who previously received a dose during routine immunization or a catch-up campaign.

#### 5.2 Burnet Institute

**Figure S5.2.1: Model structure for the Burnet agent-based model**

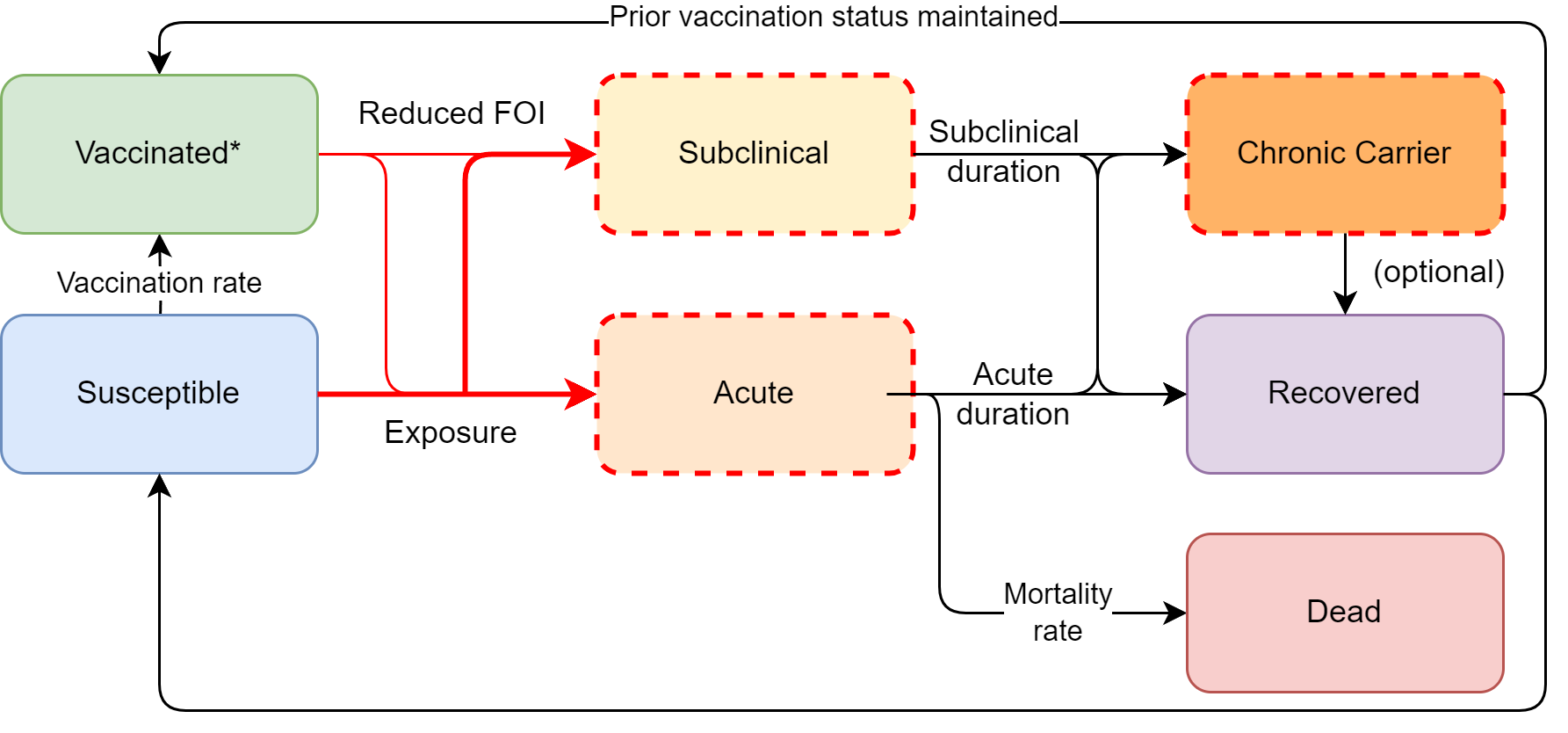

The Burnet group developed an agent-based model using the *Starsim* framework to simulate typhoid disease dynamics. Diffuse direct transmission is modelled as a proxy for all sources of transmission, including environmental transmission. In this way, the overall force of infection is captured without explicitly modelling each individual transmission route or requiring detailed data on specific pathways. Beyond the common features among all the models, infected individuals in this model may become chronic carriers based on age- and sex-specific probabilities and are assumed to have life-long carriage. Natural and vaccine immunity provide leaky protection against both infection and symptoms, and any prior infection is assumed to be fully protective against subsequent acute infections.

To reproduce the incidence of each incidence setting, we estimated the transmission coefficient, duration of natural immunity, relative infectiousness of chronic carriers, age-specific relative risk of infection, and the symptomatic fraction of primary infections. We calibrated the model by adjusting parameters such as the transmission rate or probability of an acute infection to minimize the differences between the model projections and archetypal acute incidence data. For each burden setting, the model was calibrated to a steady state that matched the observed age-specific incidence of typhoid. To introduce a level of uncertainty (10%) into our calibrated set of parameters, we drew samples from distributions centered around the parameter values, with standard deviations set to 10% of their respective means. Because model simulations can yield a broad range of outcomes for a given set of key parameters, we filtered simulations based on how realistically they represented the observed data. Specifically, for each age group, the mean incidence over the analysis period was required to fall within 25% of the observed age-specific incidence, and the trend in incidence (i.e., the slope) over this same period could not exceed 25%. Figure S5.2.2 compares age-specific typhoid incidence per 100,000 person-years across the baseline incidence settings (medium, high, and very high) with the modelled incidence, illustrating the fit of the model to the observed data for each age group.

Additionally, the model was fitted under slow and fast vaccine immunity waning scenarios using data from the TyVAC Malawi [16,17] and TyVOID Bangladesh [18,19] studies, respectively. We estimated waning rates by explicitly simulating the trial cohort structure and vaccine coverage, fitting VE_0_ and waning rates such that the model’s incidence-based estimates of vaccine efficacy matched the observed trial data (Figures S5.2.3 and S5.2.4). In the fast-waning scenario, this adjustment involved age-group-specific efficacy and waning rates. The infectiousness of chronic carriers was calibrated to align with the observed indirect protection in the TyVAC-Bangladesh cluster RCT.

**Table S5.2.1 Fitted and calibrated parameters by incidence setting for the Burnet agent-based model (Median (Q1 – Q3))**

| **Medium** | |
| --- | --- |
| Transmission Rate | 0.2542 (0.2426 – 0.2664) |
| Probability of acute infection | 0.2966 (0.2770 – 0.3168) |
| Duration of natural immunity | 4.9533 (4.6443 – 5.3030) |
| Relative infectiousness of chronic carriers | 0.1970 (0.1854 – 0.2099) |
| Relative susceptibility of 0 to 2-year-olds | 0.3359 (0.3140 – 0.3591) |
| Relative susceptibility of 2 to 5-year-olds | 0.7014 (0.6627 – 0.7440) |
| Relative susceptibility of 5 to 10-year-olds | 0.9230 (0.8658 – 0.9792) |
| Relative susceptibility of 10 to 15-year-olds | 1 (1 – 1) |
| Relative susceptibility of >15-year-olds | 0.5319 (0.5022 – 0.5641) |
| **High** | |
| Transmission Rate | 0.3357 (0.3210 – 0.3493) |
| Probability of acute infection | 0.3509 (0.3292 – 0.3733) |
| Duration of natural immunity | 5.1090 (4.7538 – 5.4584) |
| Relative infectiousness of chronic carriers | 0.1977 (0.1860 – 0.2089) |
| Relative susceptibility of 0 to 2-year-olds | 0.4111 (0.3879 – 0.4341) |
| Relative susceptibility of 2 to 5-year-olds | 1.2661 (1.1902 – 1.3391) |
| Relative susceptibility of 5 to 10-year-olds | 1.0520 (0.9888 – 1.1055) |
| Relative susceptibility of 10 to 15-year-olds | 1 (1 – 1) |
| Relative susceptibility of >15-year-olds | 0.4180 (0.3932 – 0.4420) |
| **Very High** | |
| Transmission Rate | 0.4665 (0.4515 – 0.4814) |
| Probability of acute infection | 0.6380 (0.6011 – 0.6712) |
| Duration of natural immunity | 5.0950 (4.7562 – 5.4612) |
| Relative infectiousness of chronic carriers | 0.1990 (0.1868 – 0.2133) |
| Relative susceptibility of 0 to 2-year-olds | 0.3026 (0.2839 – 0.3211) |
| Relative susceptibility of 2 to 5-year-olds | 1.1031 (1.0309 – 1.1739) |
| Relative susceptibility of 5 to 10-year-olds | 0.9930 (0.9307 – 1.0598) |
| Relative susceptibility of 10 to 15-year-olds | 1 (1 – 1) |
| Relative susceptibility of >15-year-olds | 0.9209 (0.8632 – 0.9842) |

**Figure S5.2.2: Model fit to the incidence data for the Burnet agent-based model** (a) Model-predicted absolute acute incidence (bars including uncertainty ranges) by age group over 20 years, with chronic carrier prevalence overlaid (red line) and (b) Comparison of model-predicted incidence (black crosses including uncertainty ranges) with observed data (bars) for medium (left), high (middle), and very high (right) incidence settings.
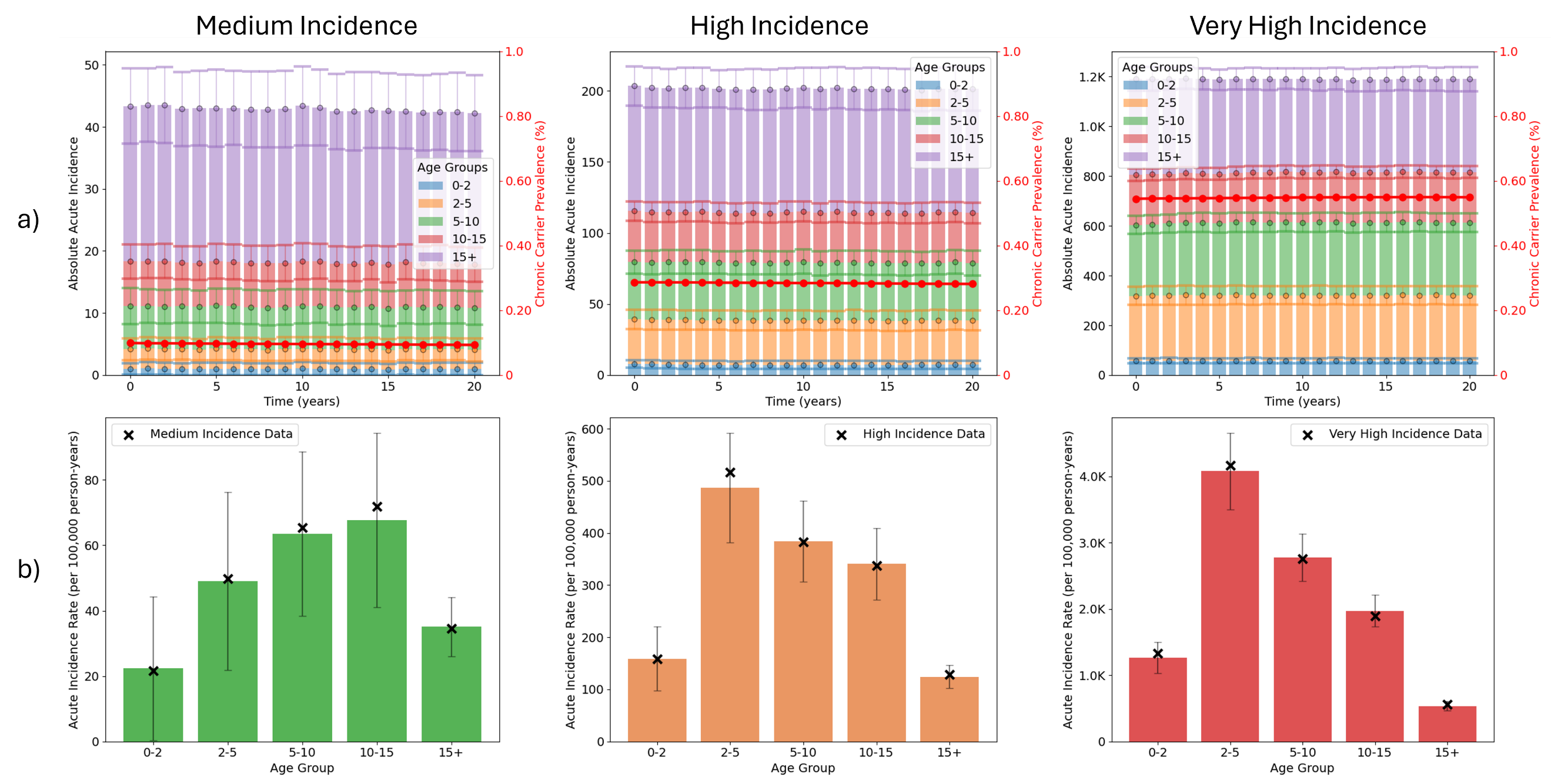

**Figure S5.2.3: Model fit to the Bangladesh VE data for the Burnet agent-based model** Total, overall, and indirect vaccine protection over time after campaign introduction by age group. Points show model predictions (median and 5^th^ – 95^th^ percentile ranges) and observed data, with the dashed line representing the input VE.

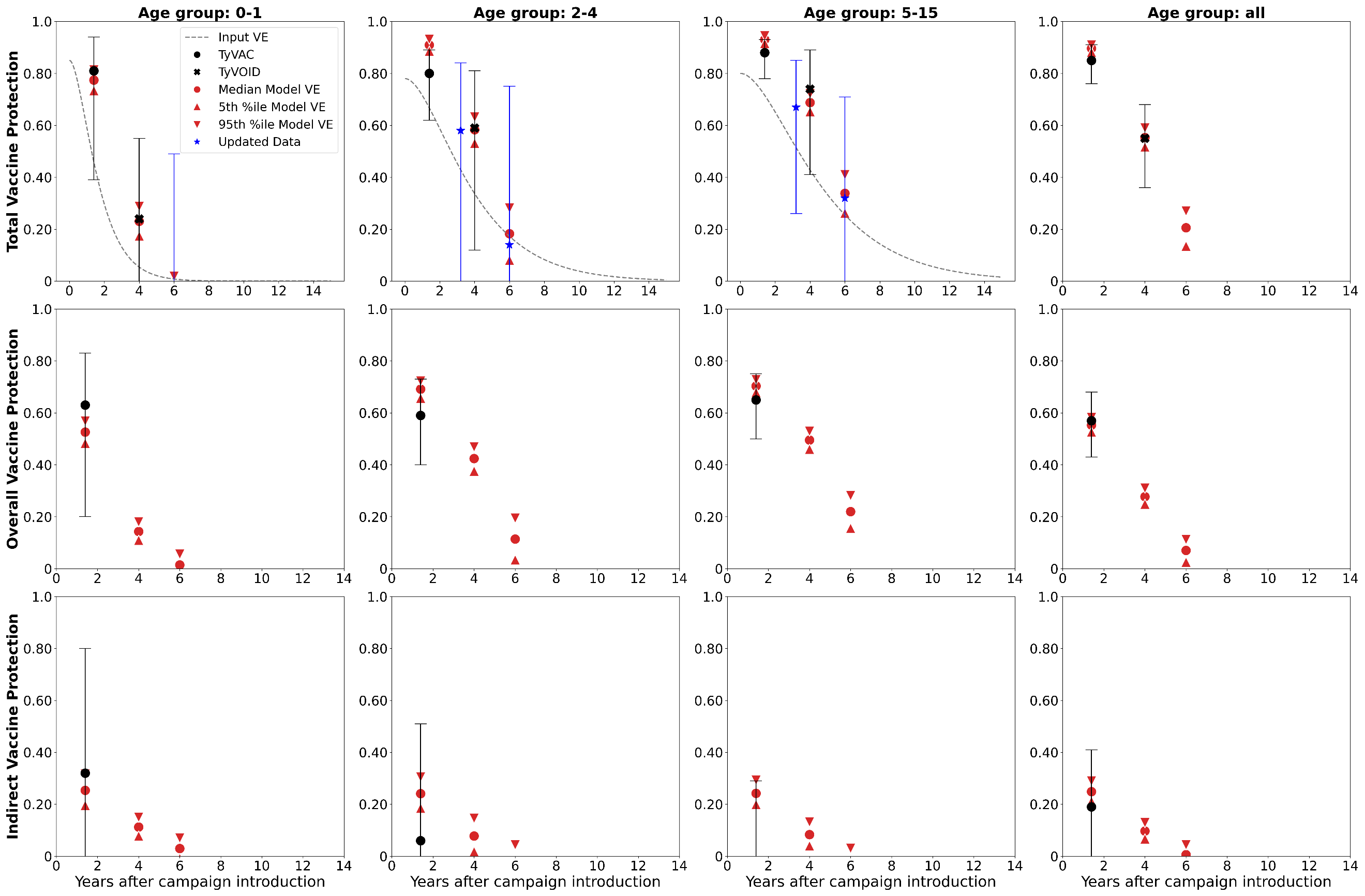

**Figure S5.2.4: Model fit to the Malawi VE data for the Burnet agent-based model**
Vaccine efficacy over time after campaign introduction by age group. Lines show model predictions (median and 5^th^ – 95^th^ percentile ranges) and observed data, with the grey dashed line representing the input VE.

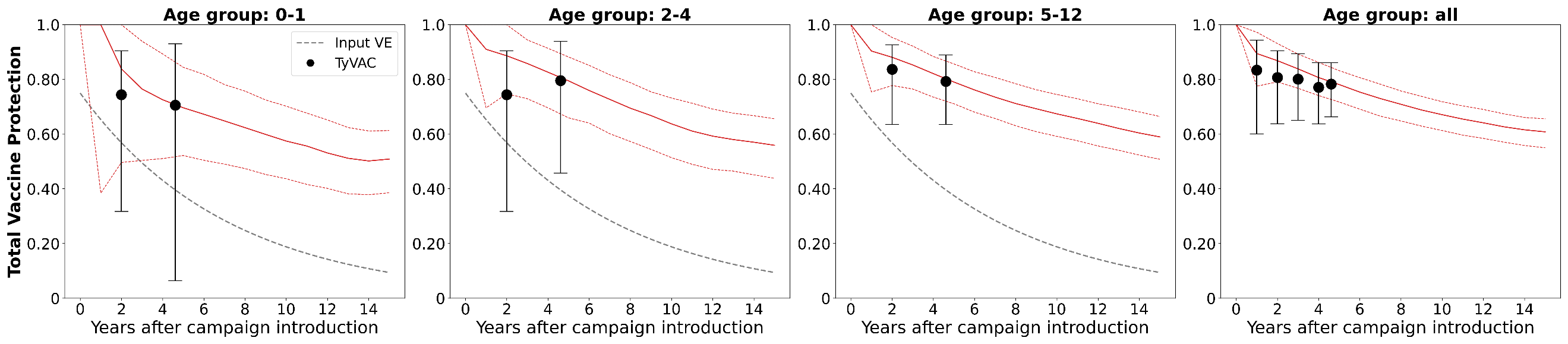

#### 5.3 Institute for Disease Modeling (IDM)

**Figure S5.3.1: Model structure for the IDM agent-based model**

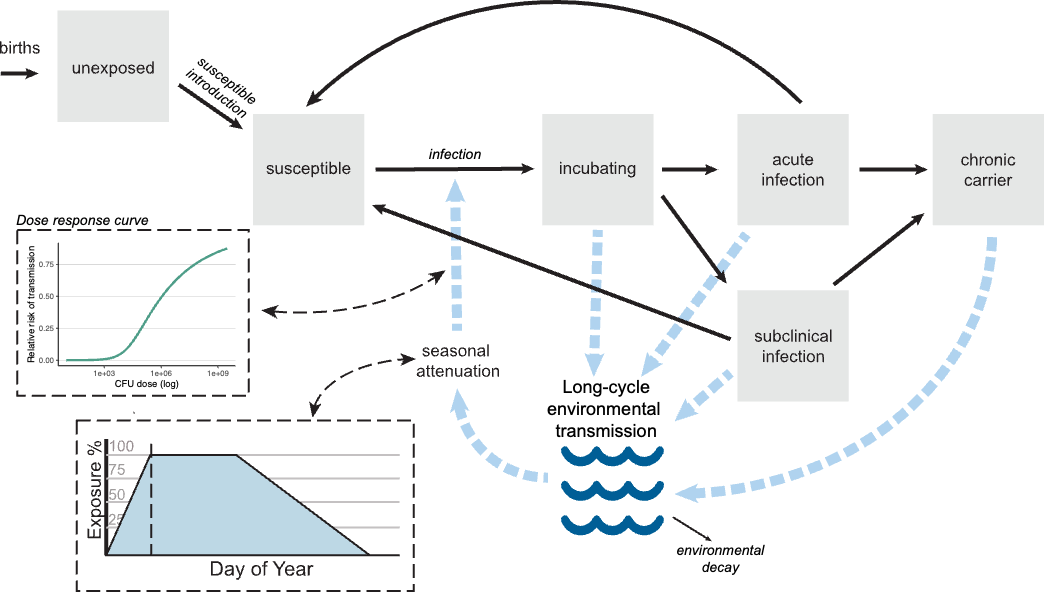

The IDM model, Typhoidsim, is an agent-based model developed in the *Starsim* platform as an update to the previously developed EMOD model [24]. Individuals in this model are immune to infection upon birth due to maternal immunity and/or reduced environmental exposure. Individuals enter the susceptible population with an age-specific probability, governed by an exponential curve whose concavity is a fitted model parameter and a fitted maximum age by which all individuals are considered susceptible.

Individuals can be infected via long-cycle transmission through exposure to environmental reservoirs, which is mediated by a dose-response curve. The prevalence of bacteria in the environment is also subject to seasonal attenuation, which was fit to data from Chile in a previous iteration of the model [24]. Subsequent infections occur at a reduced rate based on the number of prior infections an individual has experienced. After infection, individuals experience a latent period that lasts an average of 2 days before progressing to either subclinical or clinical infection. After infection, individuals may become lifelong chronic carriers at a rate depending on age- and sex-specific risk of gallstone disease. Latent, incubating infections (relative risk, 0.5) and chronic carriers (relative risk, 0.24) are assumed to be less infectious relative to subclinical or clinical infections. Parameters governing acute infectiousness of clinical infections, rate of susceptible introduction, natural immunity, and symptomatic infection were fit to the age distribution and overall magnitude of incidence in each archetypal setting (Table S5.3.1).

Individuals in any model state are eligible for vaccination. Vaccination is modeled as reducing susceptibility to infection by an amount proportional to age-specific vaccine efficacy (“leaky vaccination”). Vaccine protection was modeled as a gamma decay function (shape = 1 for slow waning, shape = 2 for fast waning), with individually-varying, age-specific initial response and duration of protection simulated from posterior samples generated by the statistical model of VE and waning parameters described in section 3. Vaccine coverage varied across stochastic iterations of the model, sampled from country-specific proxy vaccination data described in section 4.

To validate transmission and vaccination dynamics, we simulated routine immunization at 9 months and a one-time vaccination campaign of individuals 9 months to 15 years of age to reflect the conditions of the Bangladesh cluster RCT (cRCT). We also simulated an observation process to match the age-specific person-time distribution of the cRCT. We estimated overall, total, and indirect vaccine protection in this simulated cohort and confirmed that these values were within the cRCT’s confidence intervals.

**Table S5.3.1 Fitted and calibrated parameters by incidence setting for the IDM agent-based model**

| **Medium** | |
| --- | --- |
| Acute infectiousness | 3788.9 |
| Susceptible introduction: concavity | 3.068 |
| Susceptible introduction: maximum age | 15 |
| Protection via natural immunity per prior infection | 0.871 |
| Probability of symptomatic infection | 0.025 |
| **High** | |
| Acute infectiousness | 4925.1 |
| Susceptible introduction: concavity | -0.200 |
| Susceptible introduction: maximum age | 3 |
| Protection via natural immunity per prior infection | 0.815 |
| Probability of symptomatic infection | 0.098 |
| **Very High** | |
| Acute infectiousness | 9,713.6 |
| Susceptible introduction: concavity | -0.259 |
| Susceptible introduction: maximum age | 3 |
| Protection via natural immunity per prior infection | 0.812 |
| Probability of symptomatic infection | 0.398 |

**Figure S5.3.2: Model fit to the incidence data for the IDM agent-based model.**

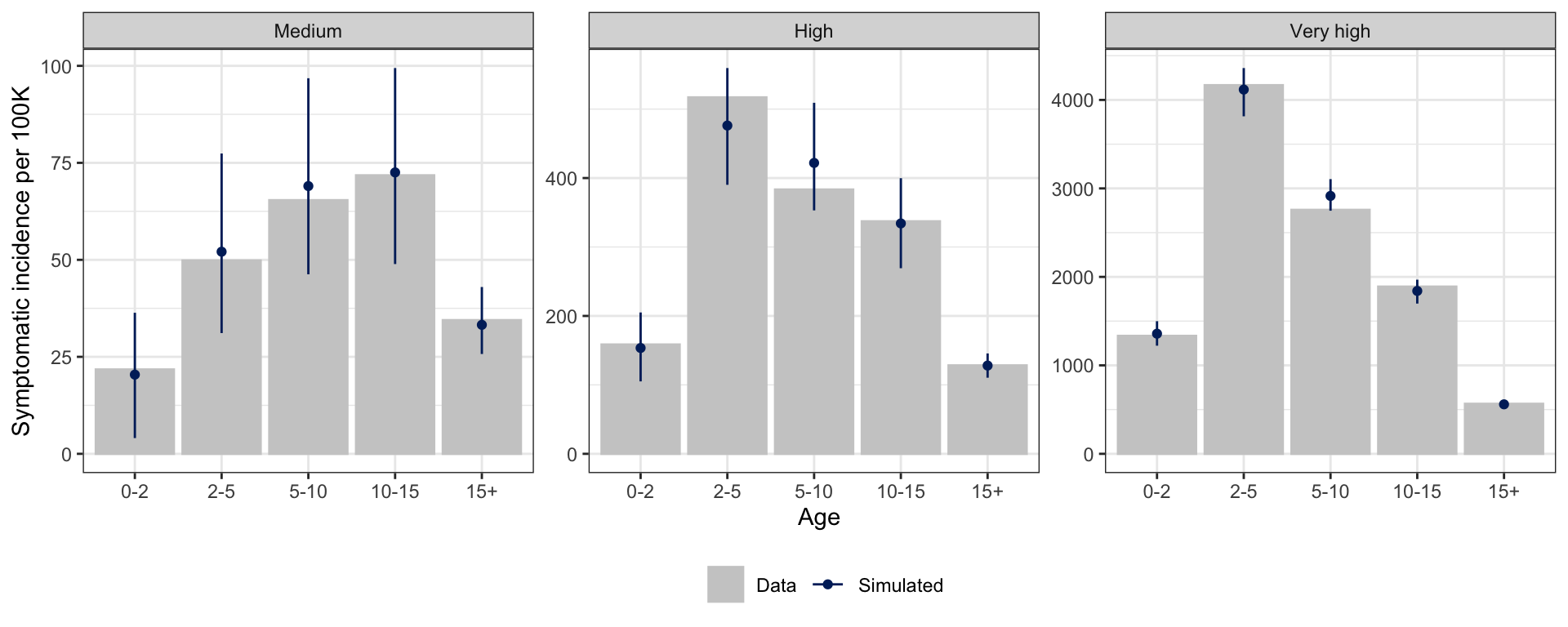

#### 5.4 Stanford University

**Figure S5.4.1: Model structure for the Stanford compartmental model**

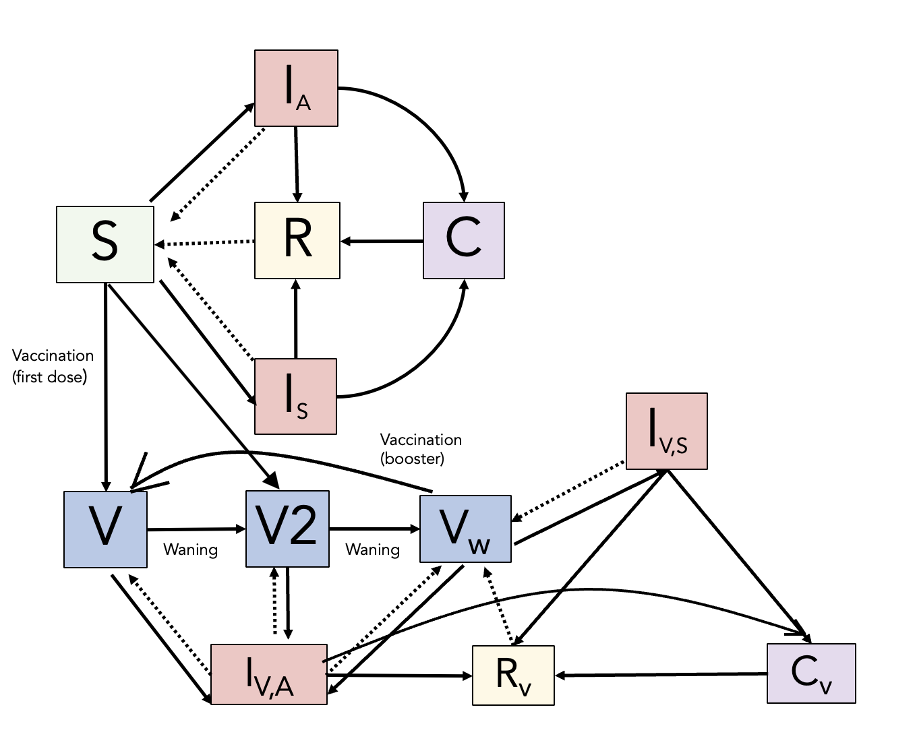

Stanford developed a compartmental model that simulates direct typhoid transmission at the population level, where individuals become infected at a rate related to the number of infectious persons and enter symptomatic and asymptomatic compartments based on an age-specific symptomatic fraction. Based on the robustness of the immune response, individuals may recover with natural immunity for some time (after which they become re-susceptible to infection), return directly to the susceptible class, or become a chronic carrier with a duration of 10 years. Vaccine protection is modeled through an initial all-or-nothing component where vaccinated individuals either obtain protection (V) against symptomatic disease or have no protection (Vw). Individuals who have vaccine protection may become asymptomatically infected (Iv,a), simulating a leaky component of vaccine protection. Protected individuals wane into the Vw compartment upon losing vaccine immunity, where they are susceptible to both symptomatic and asymptomatic infection, but can have their protection restored to initial levels by a booster dose. To ensure demographic stability in the absence of disease transmission, we used Nelder-Mead optimization to calibrate age-specific population sizes, background mortality rates, and the overall birth rate. We also applied Nelder-Mead optimization to calibrate our natural history parameters, i.e., the age-specific transmission rates, symptomatic fractions, and an overall duration of natural immunity.

The vaccine-associated compartments are governed by several key parameters, namely the risk of asymptomatic infection among those in V, the reduction in infectiousness from asymptomatically infected previously vaccinated individuals compared to the vaccine-naïve, and age-specific vaccine effectiveness and waning rates. To appropriately capture waning within our model framework and recreate indirect protection, we calibrated our model using equilibrium incidence data from the very high incidence archetype, to two sets of targets: 1) 19% indirect protection and 2) vaccine effectiveness in the different age groups at their trial-reported time intervals. We did this for the fast- and slow-waning scenarios separately. VE and indirect targets were calibrated concurrently using bounded Black-Box optimization, applying the *BlackBoxOptim* package in Julia. For the fast-waning scenario, we calibrated to VE targets for the youngest and oldest of the three age categories for which VE data were available, and fixed VE to its original value for the middle age group, which consisted of only one age. For the slow-waning scenario, we calibrated to VE targets for the two age categories for which VE data were available.

**Table S5.4.1 Fitted and calibrated parameters by incidence setting for the Stanford compartmental model**

| **Medium** | |
| --- | --- |
| Duration of natural immunity | 18.0 years |
| Transmission rate or beta (age group 1) | 3.60 /month |
| Transmission rate or beta (age group 2) | 3.68 /month |
| Transmission rate or beta (age group 3) | 5.09/month |
| Transmission rate or beta (age group 4) | 5.80/month |
| Transmission rate or beta (age group 5) | 0.845/month |
| Symptomatic fraction (age group 1) | 0.00573 |
| Symptomatic fraction (age group 2) | 0.0128 |
| Symptomatic fraction (age group 3) | 0.0135 |
| Symptomatic fraction (age group 4) | 0.0139 |
| Symptomatic fraction (age group 5) | 0.0404 |
| **High** | |
| Duration of natural immunity | 19.4 years |
| Transmission rate or beta (age group 1) | 4.70/month |
| Transmission rate or beta (age group 2) | 8.52/month |
| Transmission rate or beta (age group 3) | 6.47/month |
| Transmission rate or beta (age group 4) | 3.85/month |
| Transmission rate or beta (age group 5) | 0.691/month |
| Symptomatic fraction (age group 1) | 0.0175 |
| Symptomatic fraction (age group 2) | 0.0346 |
| Symptomatic fraction (age group 3) | 0.0442 |
| Symptomatic fraction (age group 4) | 0.0612 |
| Symptomatic fraction (age group 5) | 0.117 |
| **Very High** | |
| Duration of natural immunity | 6.5 years |
| Transmission rate or beta (age group 1) | 2.52/month |
| Transmission rate or beta (age group 2) | 18.5/month |
| Transmission rate or beta (age group 3) | 12.1/month |
| Transmission rate or beta (age group 4) | 7.52/month |
| Transmission rate or beta (age group 5) | 0.993/month |
| Symptomatic fraction (age group 1) | 0.0475 |
| Symptomatic fraction (age group 2) | 0.0529 |
| Symptomatic fraction (age group 3) | 0.0754 |
| Symptomatic fraction (age group 4) | 0.0729 |
| Symptomatic fraction (age group 5) | 0.0664 |

**Figure S5.4.2: Model fit to the incidence data for the Stanford compartmental model**

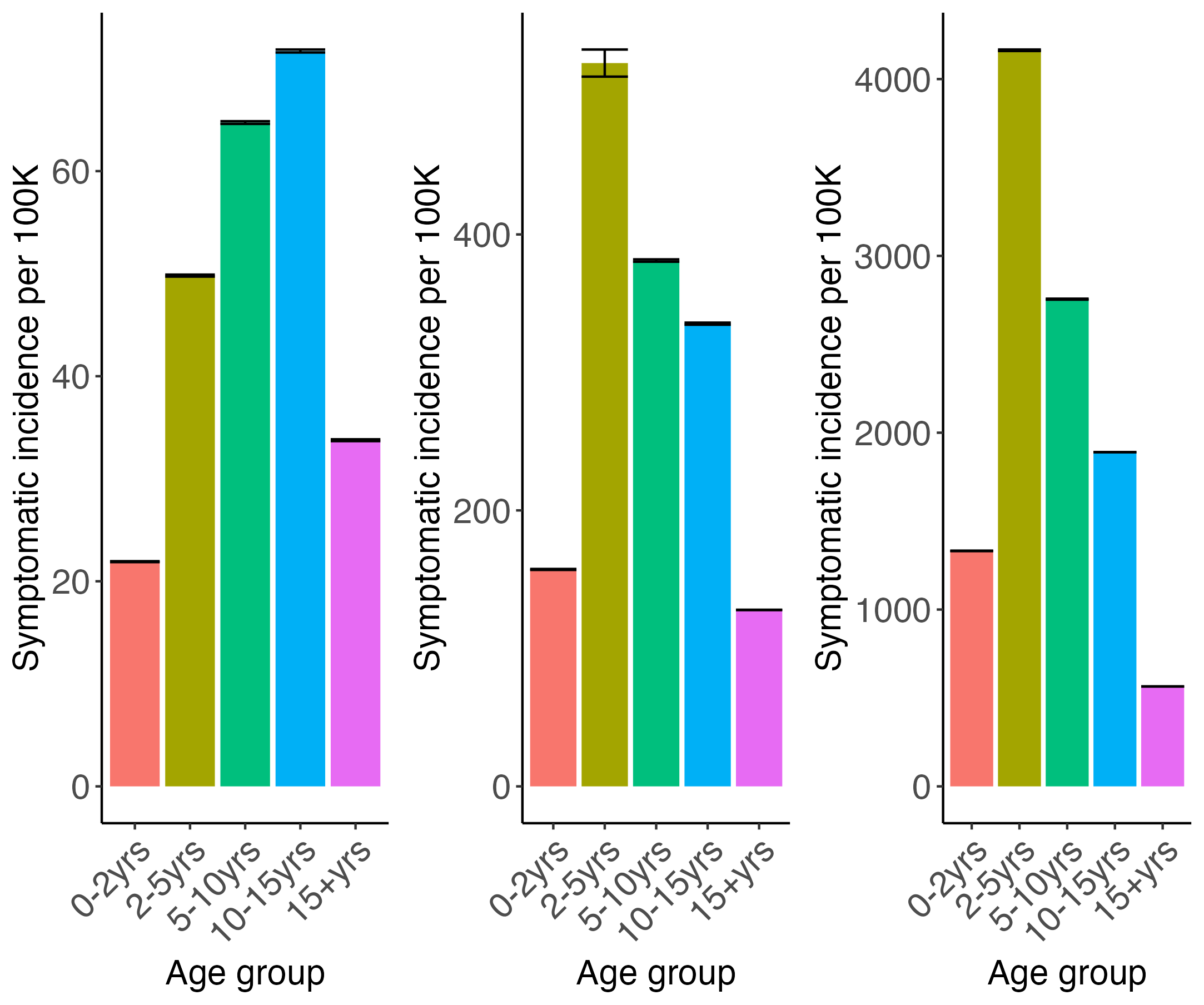

#### 5.5 Yale University

**Figure S5.5.1: Model structure for the Yale compartmental model**

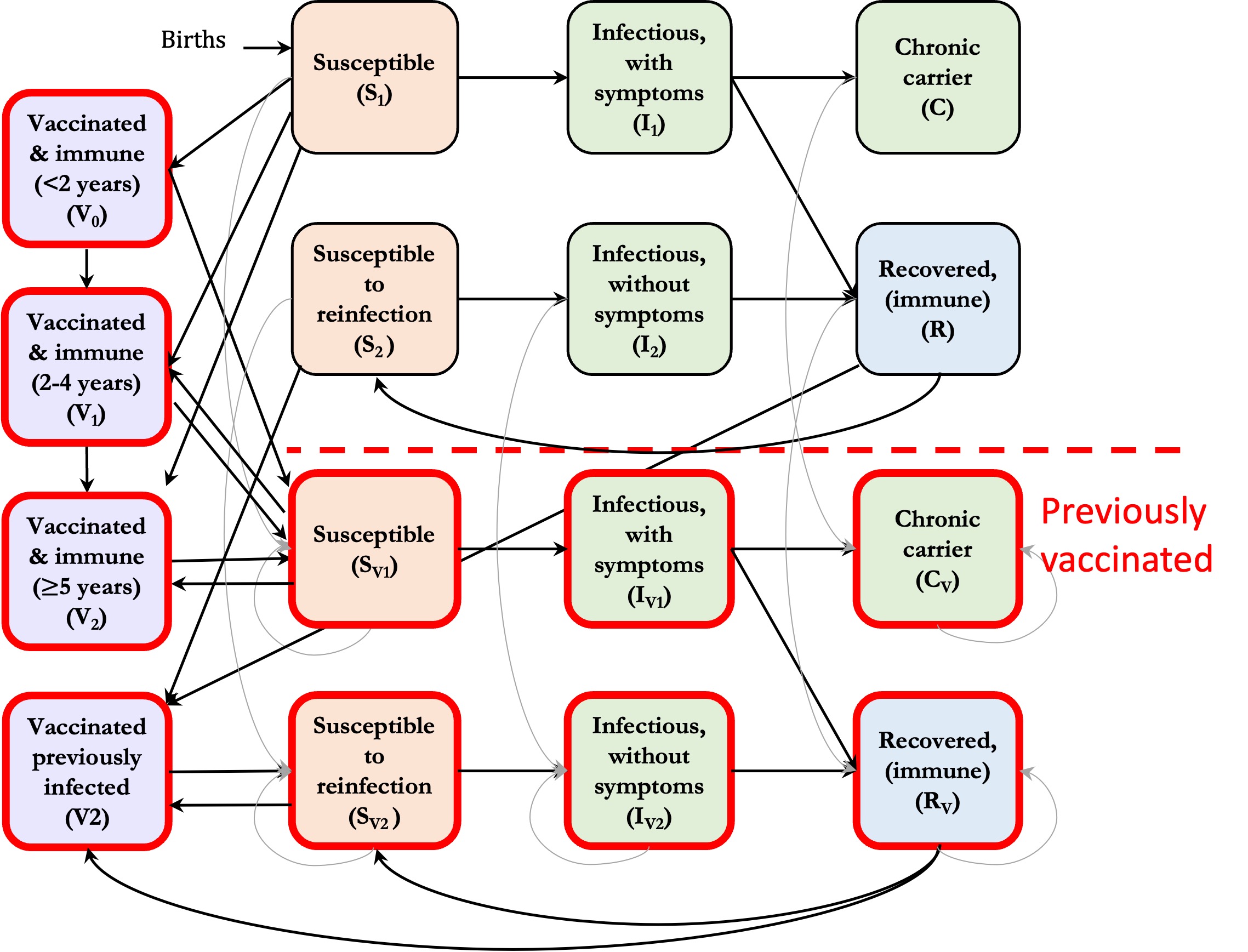

The Yale model is a compartmental transmission dynamic model that models primary and secondary infections, which are mediated by the force of infection and age-specific risk of infection. Primary infections may be clinical or asymptomatic based on the symptomatic fraction. All secondary and subsequent infections are assumed to be asymptomatic but contribute to the force of infection. Infected individuals may recover and be protected by natural immunity for an assumed duration of 2 years or become a life-long chronic carrier that fractionally contributes to the force of infection. Any individual may be vaccinated, but only those in the susceptible and recovered classes receive protection from the vaccine. All vaccinated and protected individuals enter a vaccine class according to their age group, where they are protected until they become infected due to a leaky vaccine assumption or susceptible after their immunity wanes. Those vaccinated while in the infected or carrier compartments (I1, I2, C) move to the corresponding vaccinated compartments (Iv1, Iv2, C). We fit the demographic and incidence data via Maximum Likelihood Estimation using a multinomial distribution for population age structure and a binomial distribution for age-specific incidence. Table S5.1.1 details the fitted incidence parameters, and Figure S5.5.2 shows the model fit to the age-specific incidence data by incidence setting.

**Table S5.5.1 Fitted and calibrated parameters by incidence setting for the Yale compartmental model.**

| **Medium** | |
| --- | --- |
| R0 Equation (dependent on rC) | 2.986 + 18.299rC + -10.847rC^2 |
| Relative risk of infection for 0 to 2-year-olds | 0.23 |
| Relative risk of infection for 2 to 5-year-olds | 0.57 |
| Probability of symptomatic primary infection | 0.03 |
| Relative infectiousness of chronic carriers (rC) | Beta(8.34,36.08) |
| **High** | |
| R0 Equation (dependent on rC) | 2.341 + 34.351rC + -18.846rC^2 |
| Relative risk of infection for 0 to 2-year-olds | 0.25 |
| Relative risk of infection for 2 to 5-year-olds | 0.97 |
| Probability of symptomatic primary infection | 0.11 |
| Relative infectiousness of chronic carriers (rC) | Beta(8.34,36.08) |
| **Very High** | |
| R0 Equation (dependent on rC) | 2.484 + 45.756rC + -20.057rC^2 |
| Relative risk of infection for 0 to 2-year-olds | 0.25 |
| Relative risk of infection for 2 to 5-year-olds | 1.00 |
| Probability of symptomatic primary infection | 0.61 |
| Relative infectiousness of chronic carriers (rC) | Beta(8.34,36.08) |

**Figure S5.5.2: Model fit to the incidence data for the Yale compartmental model**

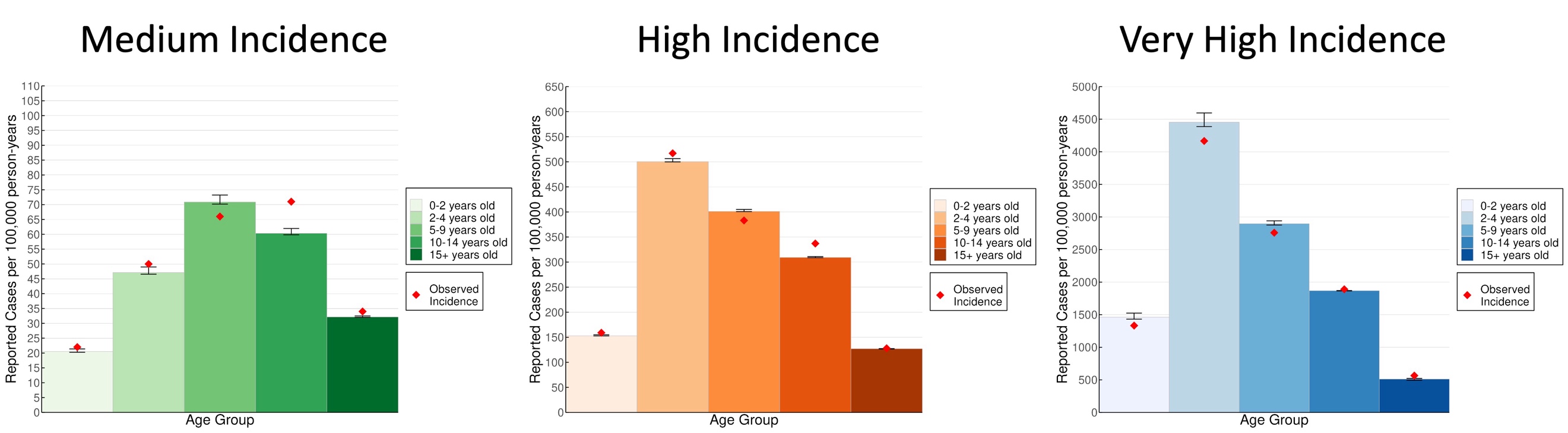

To calibrate the model to the vaccine effects observed in the Bangladesh RCT, we simulated the trial, implementing a vaccine campaign for individuals aged 9 months to 15 years in the high incidence setting. We ran this simulation 1000 times, randomly sampling a value from [0,1] for the relative infectiousness of chronic carriers (rC). We then isolated the sampled rC values that led to simulations where the estimated vaccine effects were within the RCT’s confidence intervals for overall, total, and indirect protection. The samples were used to make a beta distribution Beta(8.34,36.08) for rC that was sampled from for each epidemiological simulation. This model also used 1000 sampled estimates generated by the statistical model for the VE and waning parameters. Vaccine protection was modeled as an exponential decay function for slow waning and a gamma decay function (shape = 2) for fast waning, randomly sampling from the posterior distribution across the epidemiological simulations.

### 6 Cost-Effectiveness Analysis

#### 6.1 Scenarios

**Figure S6.1.1: Scenario combinations for regional cost-effectiveness parameters, incidence setting, and waning of vaccine effectiveness assumptions.** Each set of scenarios was evaluated for a total of 12 analyses.

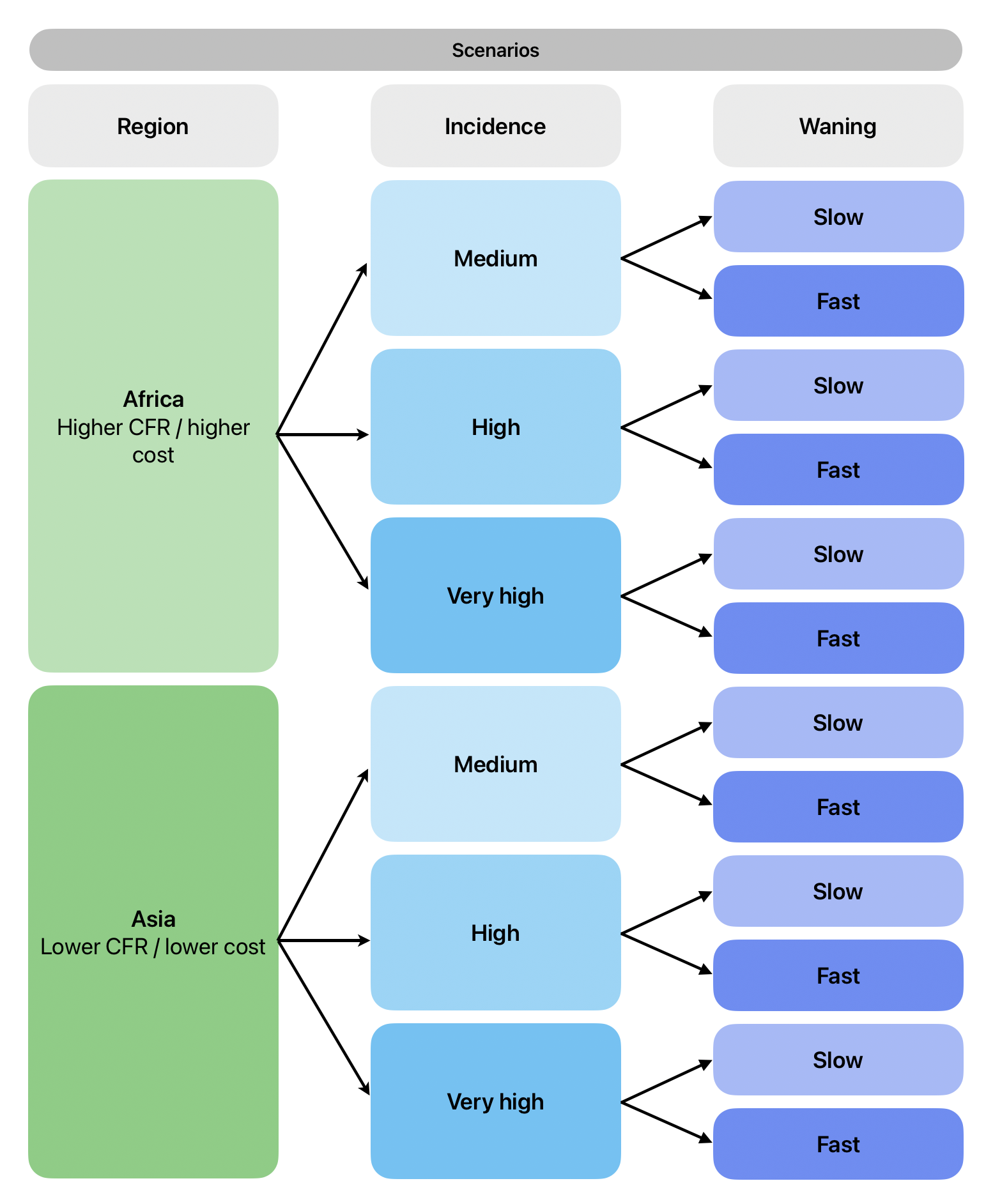

#### 6.2 Cost-effectiveness model structure

**Figure S6.2.1: Probability tree for typhoid treatment and outcomes.** Each branch indicates a unique treatment pathway that individuals in the model may experience. Green boxes indicate states that are associated with age-stratified treatment costs.

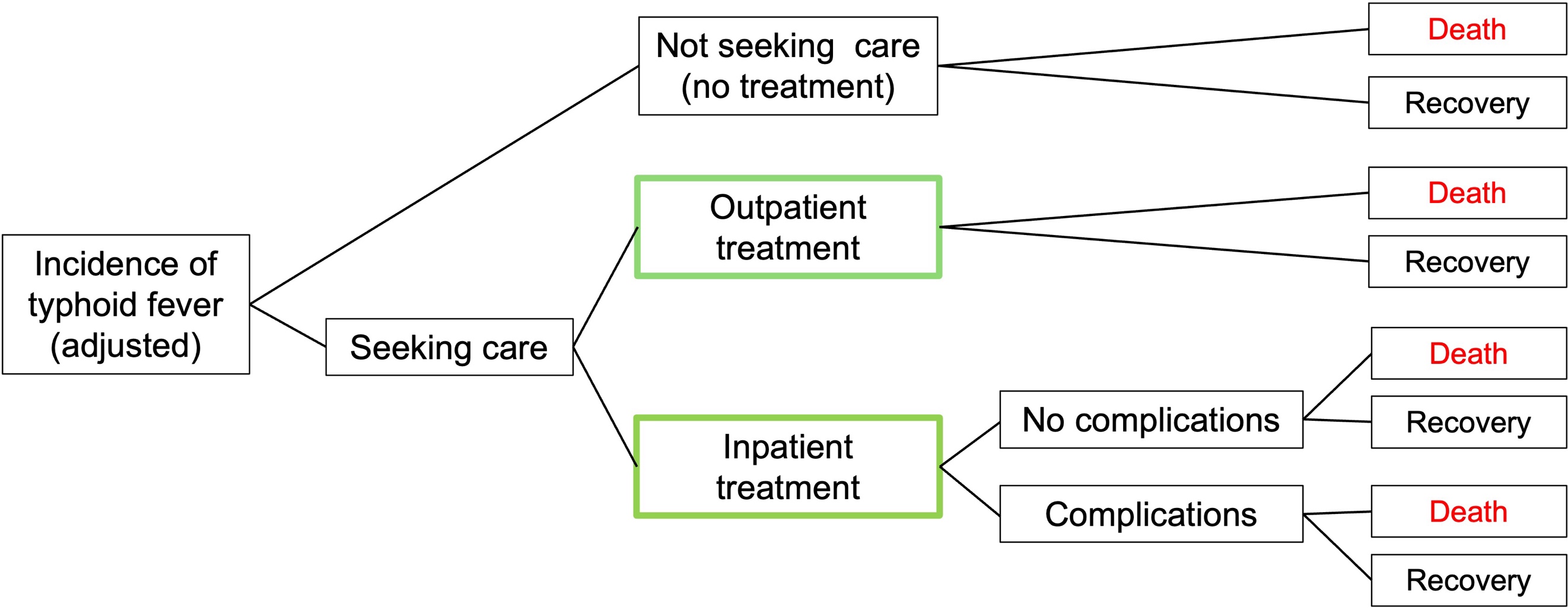

Yearly symptomatic incidence of typhoid fever from the epidemiological simulations of each model were used as inputs for our cost-effectiveness model simulations. Of those with symptoms, a fraction sought care and received either outpatient or inpatient treatment. Costs are age-stratified for outpatient and inpatient treatment; pediatric costs apply to individuals aged 15 years or younger, and adult costs apply to individuals aged 16 years or older. We model inpatient complications (e.g., due to ileal perforation) separately from inpatient treatment with no complications. The costs are the same due to a lack of data on the relative cost of complications compared to no complications, but separate disability weights were applied.

We planned to incorporate a distinct treatment pathway for antimicrobial-resistant (AMR) cases in our model structure; however, there was insufficient data to fully isolate costs and outcome probabilities associated with AMR. Therefore, we assumed that the treatment costs and probabilities and CFR estimates we used encompass both resistant and sensitive cases, as the source studies we identified were primarily conducted in locations with a high prevalence of multidrug-resistant or fluoroquinolone non-susceptible cases.

Each modeling group built a cost-effectiveness model following the same structure and sampled from harmonized uncertainty distributions for each stochastic parameter (informed by estimates from the literature as described below). Three of the modeling groups (Burnet, Stanford, and Yale) used the output from 1,000 epidemiological simulations and ran one cost-effectiveness simulation for each epidemiological simulation, resulting in a total of 1,000 economic simulations. IDM ran 250 economic simulations for each of 250 epidemiological simulations for a total of 62,500 economic simulations.

#### 6.3 Parameters

**Table 6.3.1: Cost-effectiveness parameters**

| **Parameters** | **Africa** | **Asia** | | | **Distribution** | **Source** |
| --- | --- | --- | --- | --- | --- | --- |
| ***Healthcare-seeking parameters*** | | | | | | |
| Probability of seeking care | 0.58 (CI: 0.42, 0.77) | | | | Beta | [25] |
| Probability of inpatient treatment | 0.24 (CI: 0.08, 0.53) | 0.10 (CI: 0.03, 0.28) | | | Beta | [3,26] |
| Probability of outpatient treatment | Compliment of P(inpatient treatment) | | | | Beta | [3,26] |
| Probability of complications | 0.32 (CI: 0.20, 0.47) | 0.02 (CI: 0.001, 0.36) | | | Beta | [3,26] |
| ***Severity parameters*** | | | | | | |
| Duration of severe illness | 20.0 days (IQR: 14.0 - 44.0 days) | | | | Gamma | [27] |
| Duration of moderate illness | 14.0 days (IQR: 7.0 - 25.0 days) | | | | Gamma | [27] |
| Duration of untreated illness | 14.0 days (IQR: 8.0 - 25.0 days) | | | | Gamma | [27] |
| Case fatality risk (CFR) | 0.81%  (CI: 0.10%, 6.22%) | | | 0.16%  (CI: 0.07%-0.40%) | Beta | [28] |
| ***Disability-adjusted life year (DALY) weights*** | | | | | | |
| Disability weight, severe with complications | 0.324 (CI: 0.220,0.442) | | | | Beta | [29] |
| Disability weight, severe | 0.133 (CI: 0.0885, 0.190) | | | | Beta | [29] |
| Disability weight, moderate | 0.0506 (CI: 0.0324, 0.0741) | | | | Beta | [29] |
| Disability weight, untreated | 0.0506 (CI: 0.0324, 0.0741) | | | | Beta | [29] |
| Life expectancy | 64.0 years | | 74.8 years | | Fixed | [1] |
| ***Treatment costs (USD 2025)*** | | | | | | |
| Inpatient pediatric costs | $285.01  (CI: $199.29, $370.74) | $214.45  (CI: $153.75, $326.76) | | | Log Normal | [27,30] |
| Outpatient pediatric costs | $50.41  (CI: $41.16, $59.68) | $42.62  (CI: $26.34, $72.01) | | | Log Normal | [27,30] |
| Inpatient adult costs | $445.65  (CI: $295.27, $596.02) | $214.45  (CI: $153.75, $326.76) | | | Log Normal | [27,30] |
| Outpatient adult costs | $46.77  (CI: $35.18, $58.36) | $42.62  (CI: $26.34, $72.01) | | | Log Normal | [27,30] |
| ***Vaccination costs (USD 2025)*** | | | | | | |
| Vaccine procurement costs (i.e. vaccine price) | $1.50 | | | | Fixed | [31] |
| Syringes & safety equipment costs (unit price) | $0.29 (CI: $0.28, $0.31) | | | | Log Normal | [32] |
| Delivery costs, routine | $2.34  (CI: $1.96, $2.70) | $1.54  (CI: $0.94, $2.14) | | | Log Normal | [25] |
| Delivery costs, school-based | $1.42 (CI: $0.71, $2.14) | | | | Log Normal | [33] |
| Delivery costs, catch-up campaign | $0.50  (CI: $0.44, $0.56) | $0.54  (CI: $0.46, $0.61) | | | Log Normal | [25] |

##### 6.3.1 Healthcare-seeking probabilities

The probability that an individual with typhoid seeks care was based on an estimate of the relative incidence for passive versus active surveillance, as assumed by Bilcke et al. [25]. This serves as a proxy for health-seeking behavior, since passive surveillance studies capture only cases that seek care, whereas active surveillance studies capture people with typhoid who do and do not seek care.

##### 6.3.2 Severity parameters

We assumed the probability that typhoid patients receive inpatient treatment is equivalent to the proportion of severe cases of typhoid among symptomatic infections, and the complement is the probability that patients receive outpatient treatment. These probabilities, as well as the probability of complications, are based on the Severe Typhoid in Africa programme (SETA, 2016-2020), the Surveillance for Enteric Fever in Asia Project (SEAP, 2016-2019), the Strategic Typhoid Alliance across Africa and Asia (STRATAA, 2016-2018), and the National Surveillance System for Enteric Fever in India (NSSEFI) [3,6,25,34]. Specifically, for each of the two regions, we fit a beta-binomial model with random effects for study site to reported hospitalization counts among each of the study populations, using the method of moments to link the posterior of the binomial fit to a corresponding beta distribution.

Due to a lack of regional data for Africa, duration of illness parameters were generalized to both regions based on estimates from Nepal collected during the second phase of the SEAP project (SEAP II) [27]. We used the reported days unable to work for patients who did not report inpatient expenses as the duration of moderate illness due to typhoid. For the duration of severe illness, we used the same metric for patients who reported inpatient expenses. Finally, the duration of illness for untreated patients was assumed to be the days unable to work for both of these groups combined, because we assume all sequelae can occur in untreated illness. These estimates are consistent with the duration of illness used in the 2017 Global Burden of Disease (GBD) study on typhoid, which reported years lived with disability (YLDs) for typhoid [35].

##### 6.3.3 Case fatality risk (CFR)

We calculated case fatality risks (CFRs) for each region based on data from a systematic review and meta-analysis on complications and mortality of typhoid by Murthy et al. [28]. We restricted the studies to those with a median study year of 2010 or later, since the CFR for typhoid has declined since the 1960s due to improved treatment and sanitation [36]. We ran a mixed-effects meta-analysis of proportions, with the CFR as the outcome of interest and study-level random effects, separately for each region. Data on CFRs stratified by sequelae or treatment type were incomplete, so the overall CFR for each region was used uniformly in all treatment pathways.

##### 6.3.4 Disability weights

Disability weights were sourced from the GBD 2021 study’s global estimates for typhoid and were assumed to be the same for both Africa and Asia [29]. We assumed the disability weight for moderate and untreated typhoid was equivalent to the weight for acute typhoid infection. Similarly, we used the weight for intestinal perforation due to typhoid for calculating disability-adjusted life years (DALYs) for severe typhoid cases with complications. The regional life expectancy, sourced from the same data as our demographic calibration parameters, was greater for Asia than for Africa [1].

##### 6.3.5 Treatment costs

Cost estimates for Africa came from a costing cohort study conducted in Malawi that provided direct medical costs of inpatient and outpatient treatment for both children and adults [30]. The cost estimates for Asia come from SEAP II, during which a cost-of-illness study was conducted in Nepal [27]. We assumed that the direct medical costs for patients who did not report any inpatient care expenses were equivalent to outpatient costs. For patients who reported inpatient care expenses, we attributed all of their direct medical costs to inpatient treatment. There was no clear breakdown between pediatric and adult treatment costs, so we used the overall estimates for both age groups. All treatment cost parameters were adjusted for inflation and presented in 2025 US dollars.

##### 6.3.6 Vaccine costs

We do not assume Gavi support in this analysis, so the vaccine procurement cost is the UNICEF-listed cost for a middle-income country when procuring TCV doses from Biologics E or Bharat Biologics in India [31]. Syringe and safety equipment costs increase the total cost per dose to about $1.79 and came from a costing study of vaccination in 94 LMICs [32]. Strategy-specific delivery costs came from estimates used in other TCV cost-effectiveness studies [25,33]. All vaccine cost parameters were adjusted for inflation and presented in 2025 US dollars.

#### 6.4 Net-benefit framework

The primary cost-effectiveness analysis was conducted using the net-benefit framework. This method incorporates a probabilistic sensitivity analysis of the interventions of interest to identify the uncertainty in the predicted optimal strategy. We calculated the net monetary benefit (NMB) of each strategy using the equation:

$$NMB=p*\Delta E- \Delta C$$

where *p* is the willingness-to-pay threshold, $\Delta E$ is the DALYs averted compared to no vaccination, and $\Delta C$ is the incremental cost compared to no vaccination. For each stochastic simulation and vaccination strategy, we varied the WTP threshold from $0 to $2,500. Then, the NMB for each strategy was compared across simulations to calculate the probability that a given strategy had the highest NMB at a given WTP threshold. This was then plotted for each strategy as the cost-effectiveness acceptability curve (CEAC). The cost-effectiveness acceptability frontiers (CEAFs) were derived from the CEACs to highlight only the strategy that yielded the greatest average NMB at each WTP threshold. This approach indicates the optimal strategy across a range of WTP values.

### 7 Supplementary Results

#### 7.1 Model-predicted vaccine impact over time

**Figure S7.1.1 Incidence of symptomatic typhoid for booster dose strategies in the medium incidence setting for all models.**

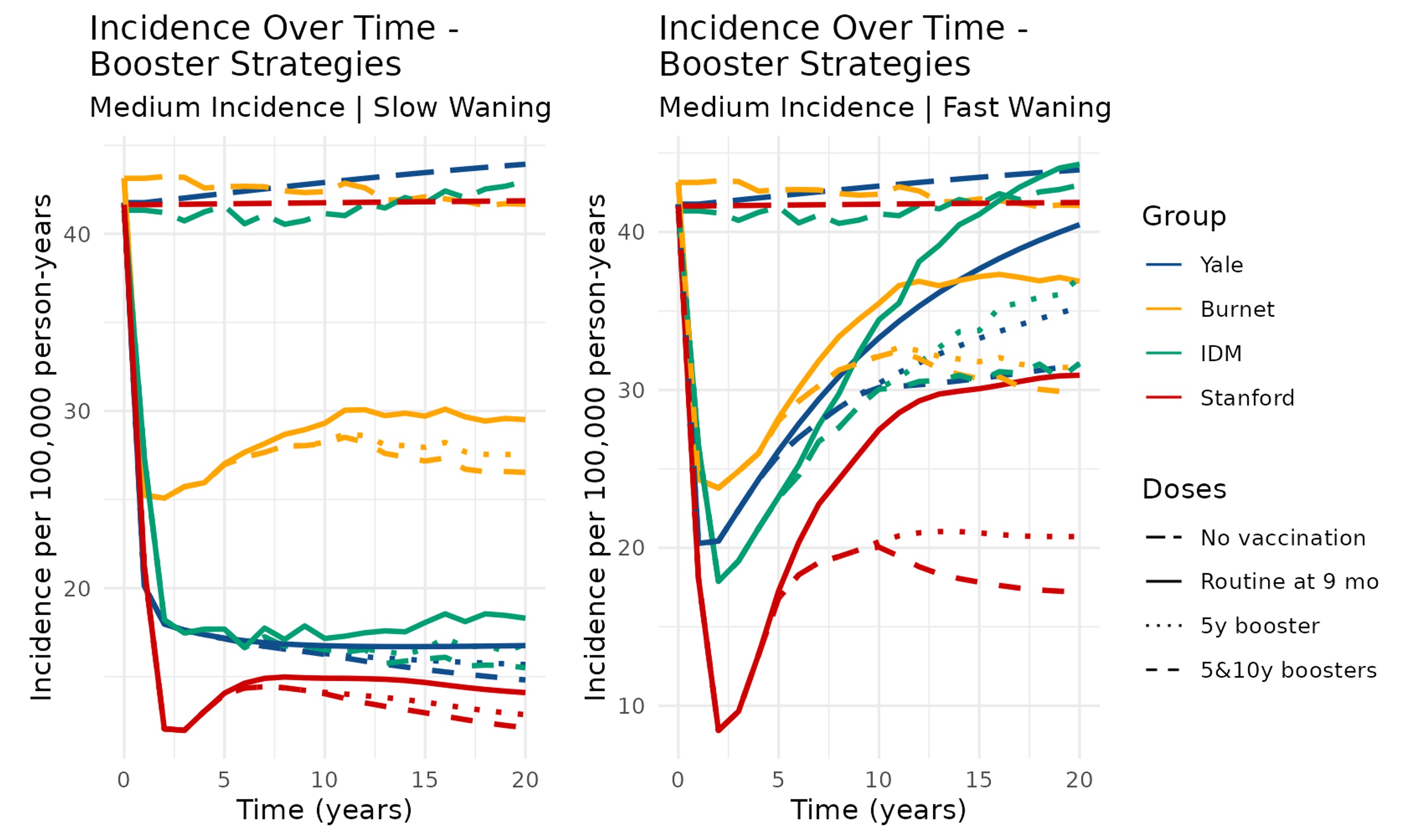

**Figure S7.1.2 Incidence of symptomatic typhoid for booster dose strategies in the high incidence setting for all models.**

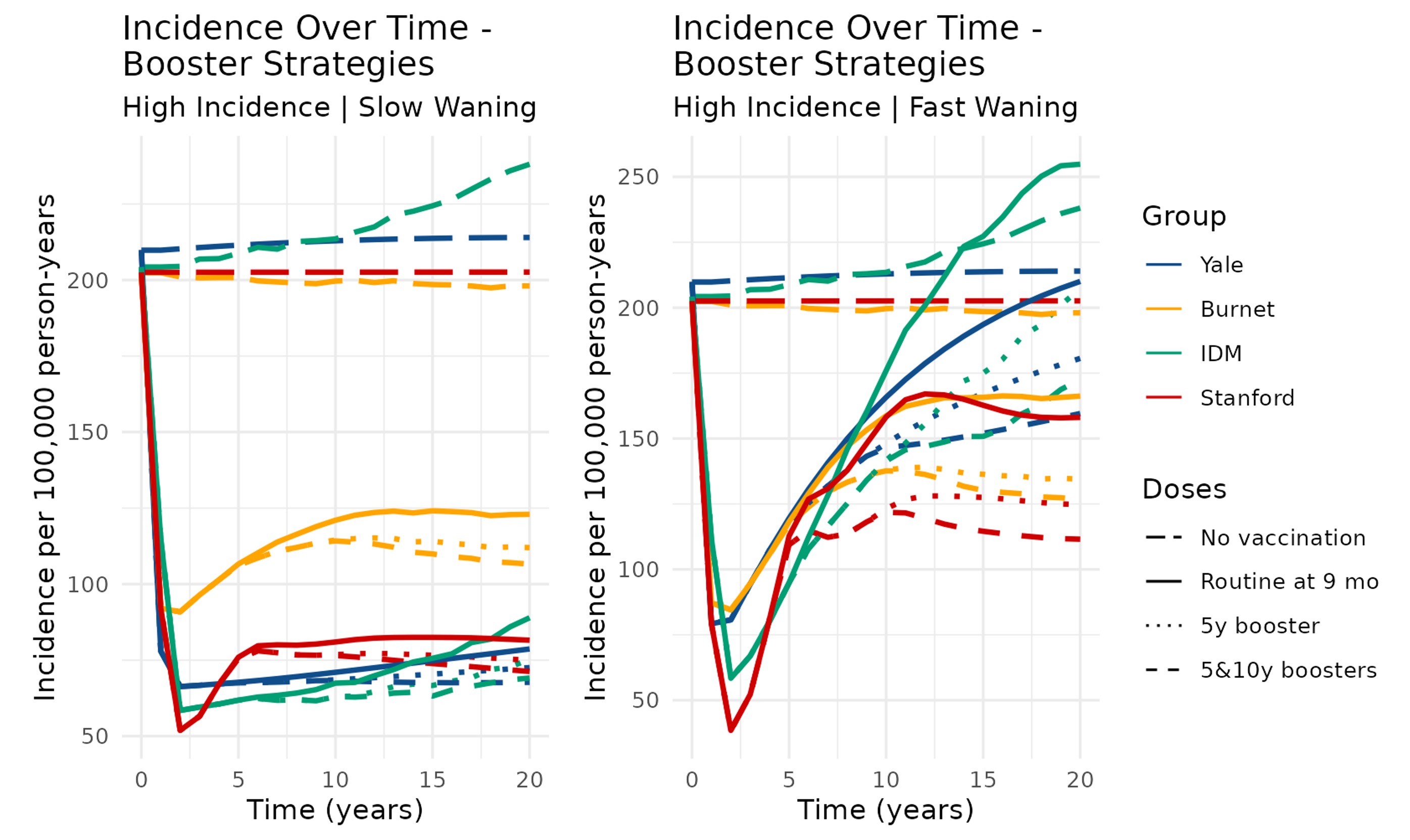

**Figure S7.1.3 Incidence of symptomatic typhoid for booster dose strategies in the very high incidence setting for all models.**

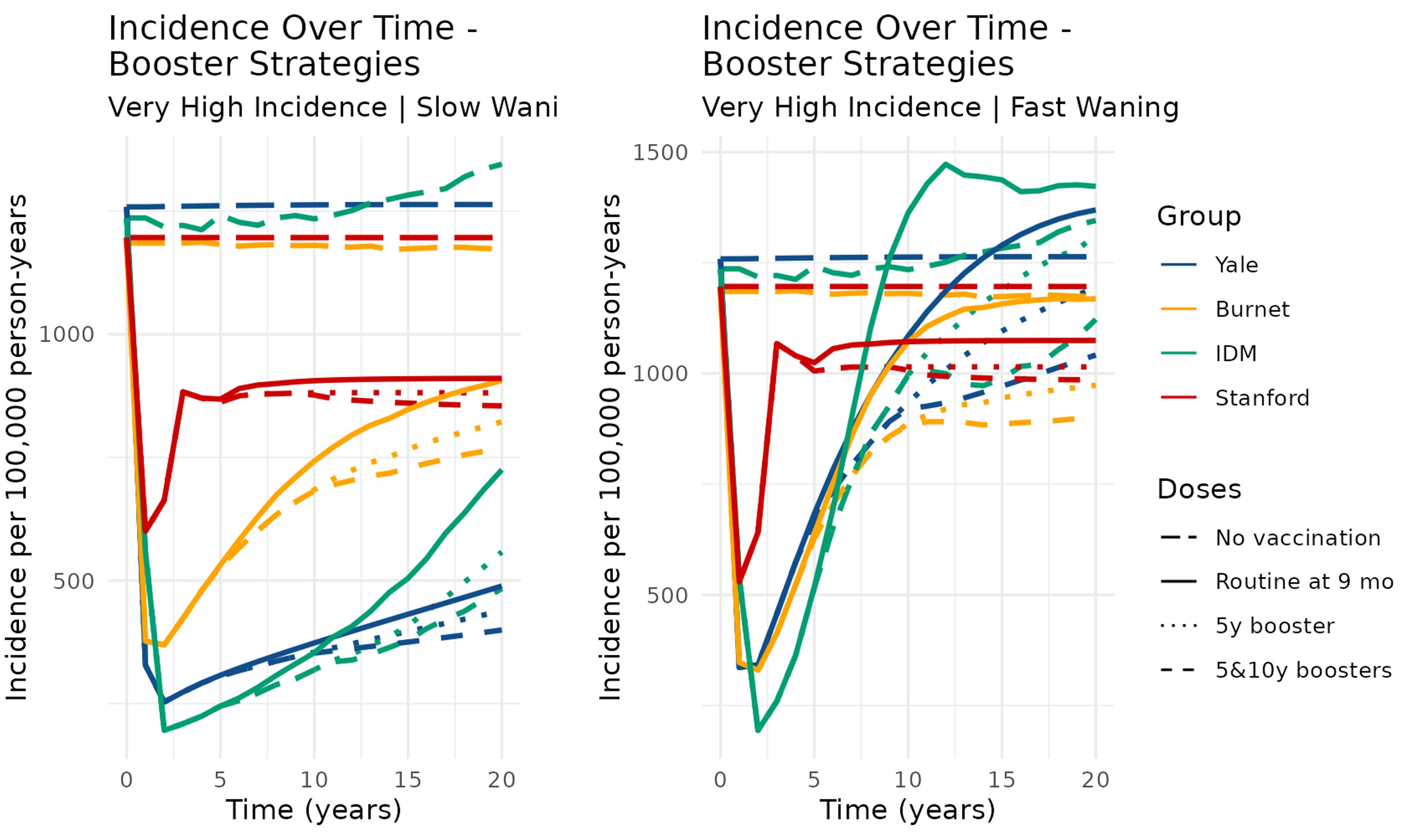

**Figure S7.1.4 Incidence curves of symptomatic typhoid for routine strategies in the medium incidence setting for all models.**

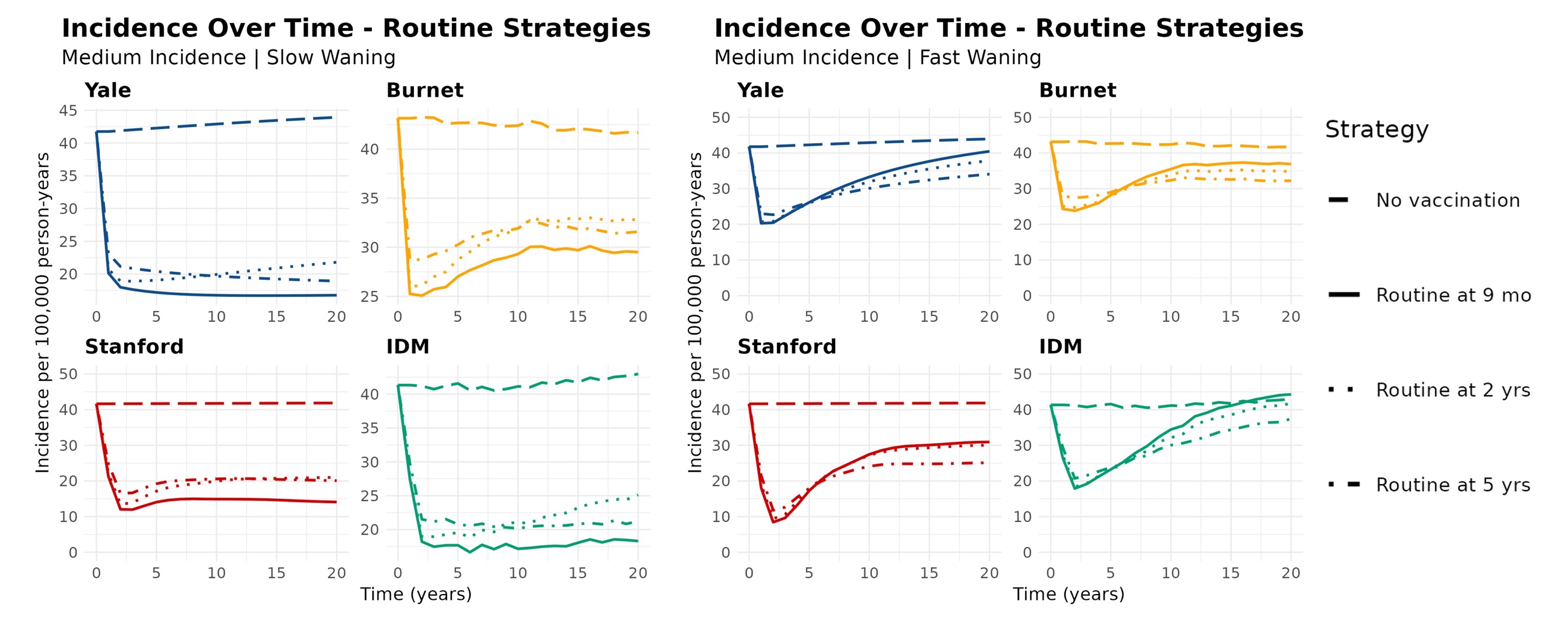

**Figure S7.1.5 Incidence curves of symptomatic typhoid for routine strategies in the high incidence setting for all models.**

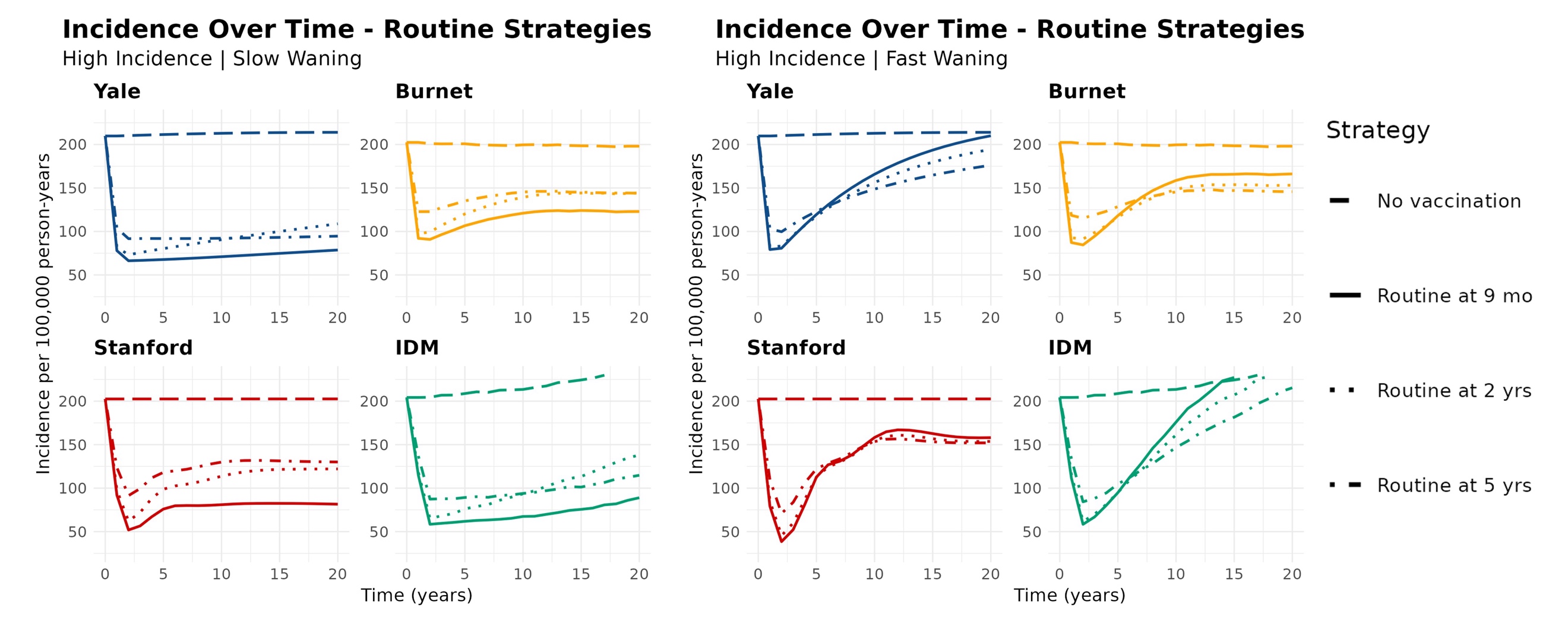

**Figure S7.1.6 Incidence curves of symptomatic typhoid for routine strategies in the very high incidence setting for all models.**

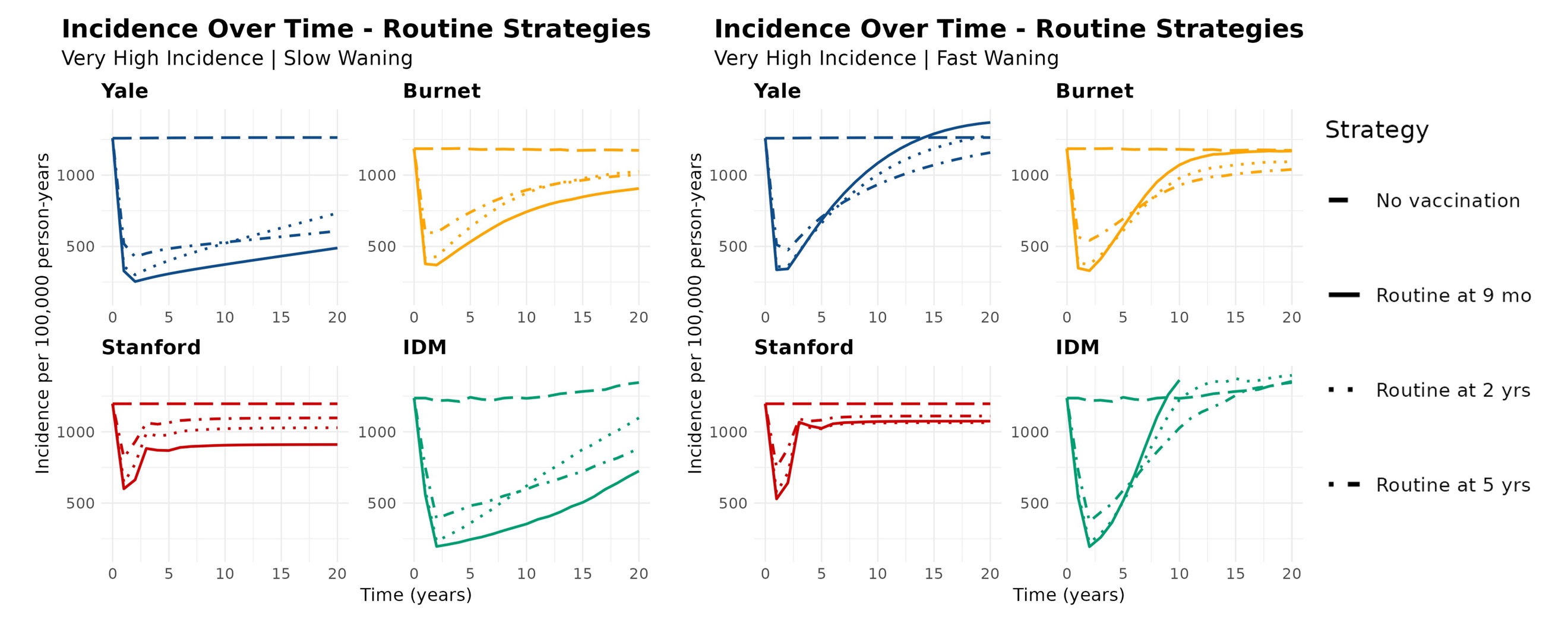

#### 7.2 Vaccine impact tables

**Table S7.2.1 Doses administered per 100,000 people for each vaccination strategy and model over a 10-year time horizon.** Data is presented as the median value from all simulations of that model and a 95% prediction interval.

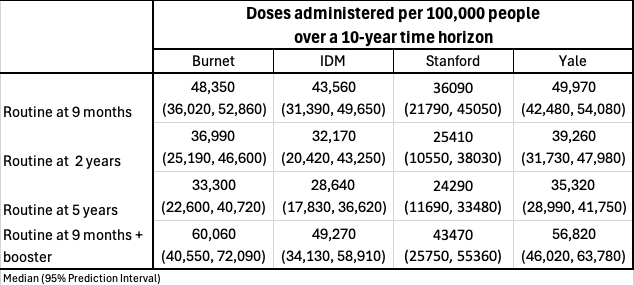

**Table S7.2.2 Vaccine impact results for each vaccination strategy and model in the medium incidence, higher-CFR/higher-cost African scenario over a 10-year time horizon.** Data is presented as the median value from all simulations of that model and a 95% prediction interval. Deaths are rounded to the nearest tenth, and all other values are rounded to the nearest whole number.

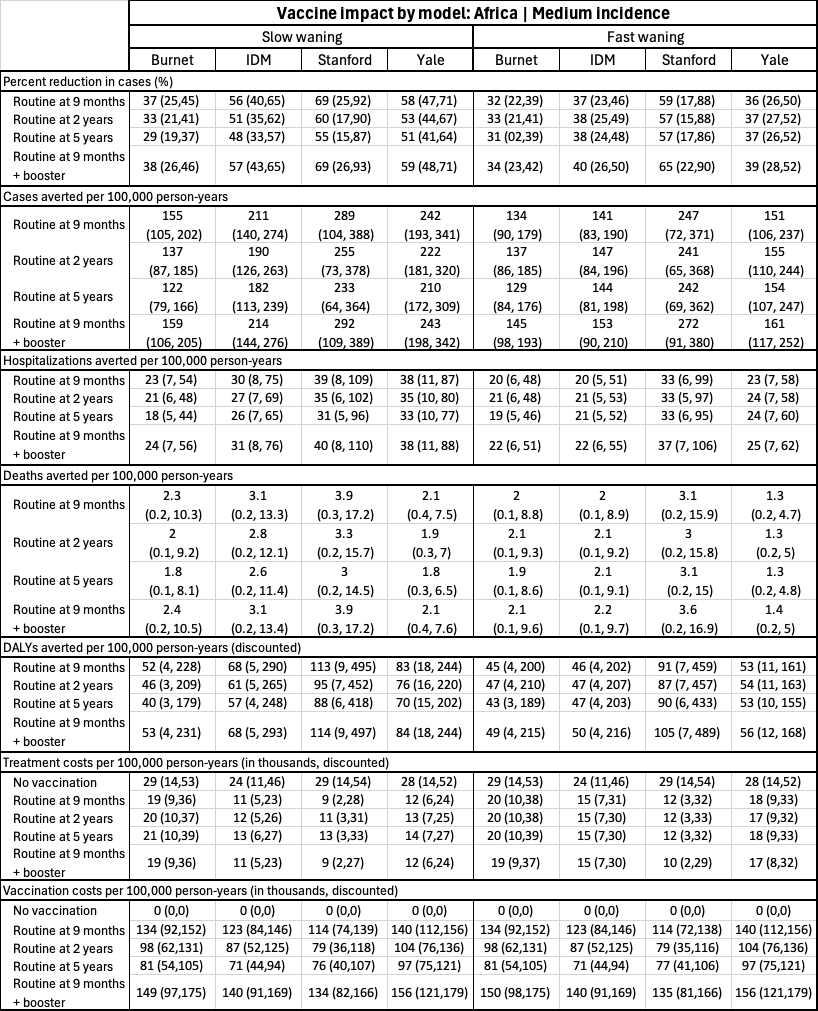

**Table S7.2.3 Vaccine impact results for each vaccination strategy and model in the high incidence, higher-CFR/higher-cost Africa scenario over a 10-year time horizon.** Data is presented as the median value from all simulations of that model and a 95% prediction interval. Deaths are rounded to the nearest tenth, and all other values are rounded to the nearest whole number.

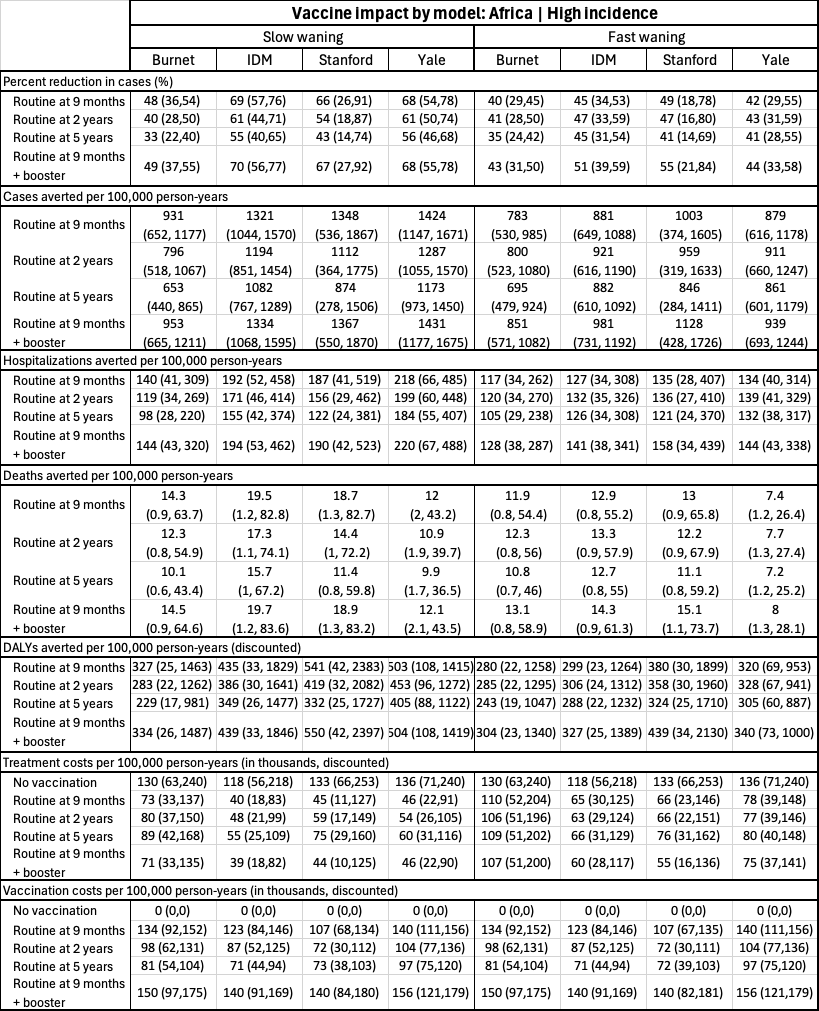

**Table S7.2.4 Vaccine impact results for each vaccination strategy and model in the very high incidence, higher-CFR/higher-cost Africa scenario over a 10-year time horizon.** Data is presented as the median value from all simulations of that model and a 95% prediction interval. Deaths are rounded to the nearest tenth, and all other values are rounded to the nearest whole number.

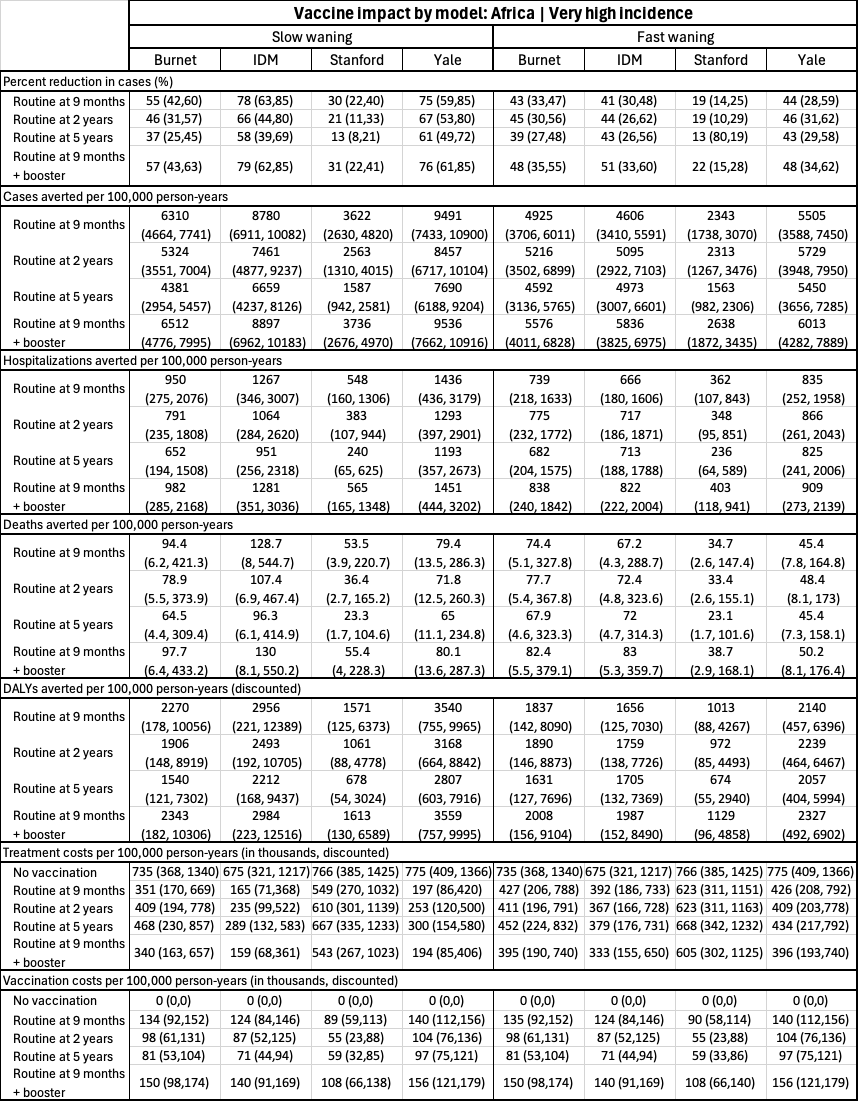

**Table S7.2.5 Vaccine impact results for each vaccination strategy and model in the medium incidence, lower-CFR/lower-cost Asia scenario over a 10-year time horizon.** Data is presented as the median value from all simulations of that model and a 95% prediction interval. Deaths are rounded to the nearest tenth, and all other values are rounded to the nearest whole number.

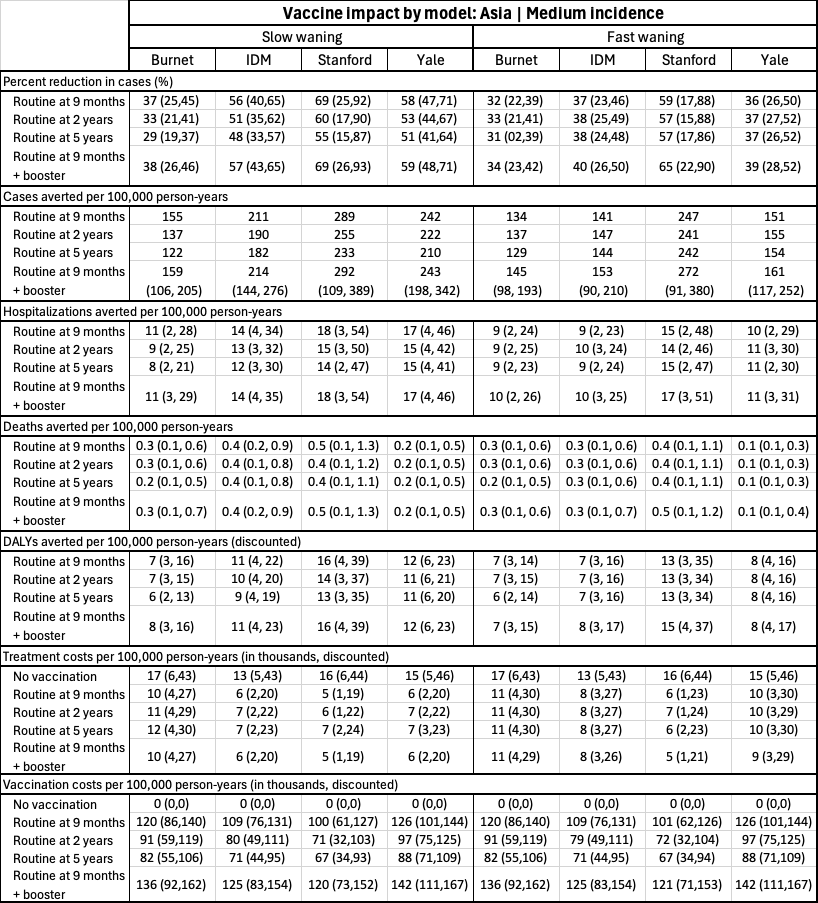

**Table S7.2.6 Vaccine impact results for each vaccination strategy and model in the high incidence, lower-CFR/lower-cost Asia scenario over a 10-year time horizon.** Data is presented as the median value from all simulations of that model and a 95% prediction interval. Deaths are rounded to the nearest tenth, and all other values are rounded to the nearest whole number.

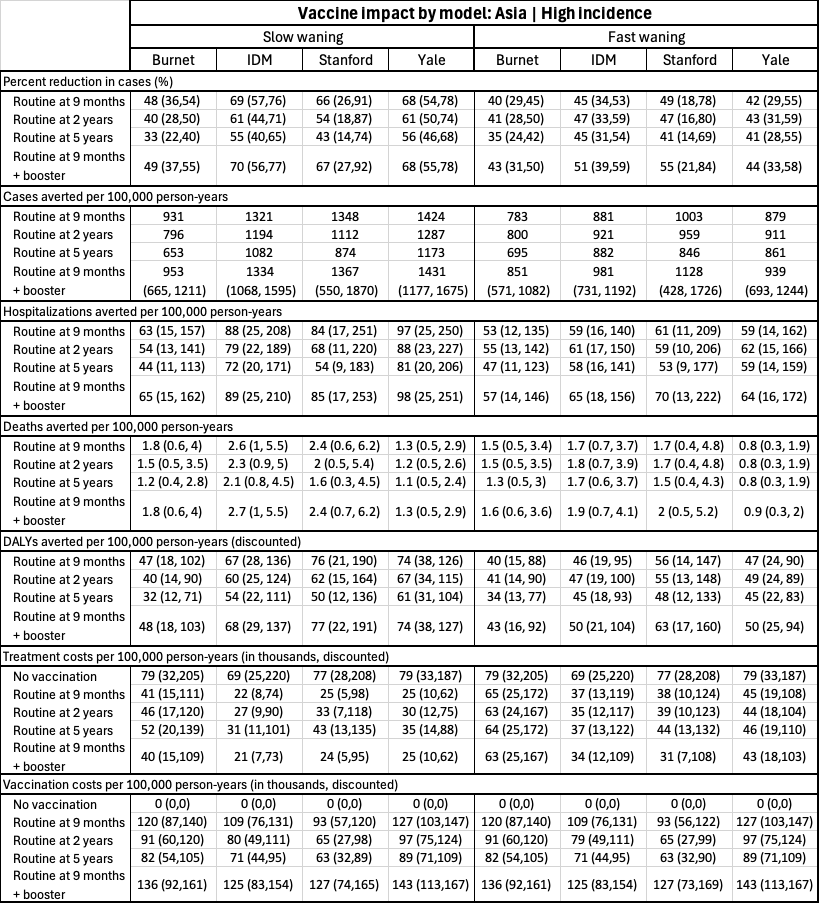

**Table S7.2.7 Vaccine impact results for each vaccination strategy and model in the very high incidence, lower-CFR/lower-cost Asia scenario over a 10-year time horizon.** Data is presented as the median value from all simulations of that model and a 95% prediction interval. Deaths are rounded to the nearest tenth, and all other values are rounded to the nearest whole number.

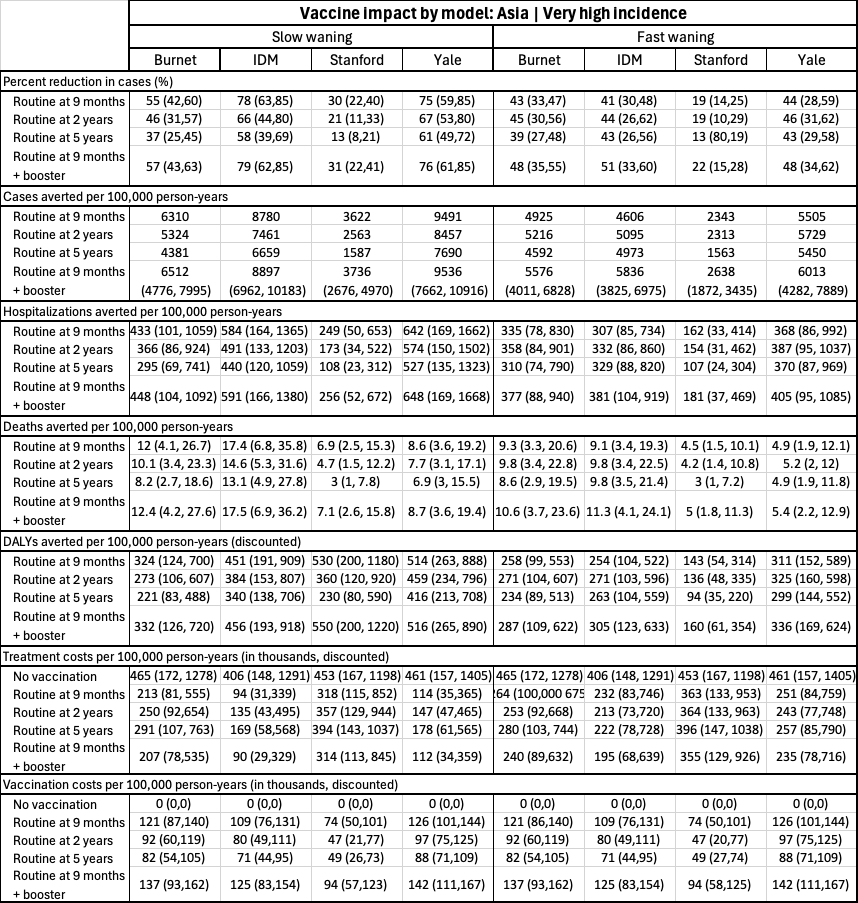

#### 7.3 Cost-effectiveness acceptability curves

**Figure S7.3.1 Cost-effectiveness acceptability curves (CEACs) for the slow-waning, higher-CFR/higher-cost Africa scenario for all incidence settings over a 10-year time horizon.** The lines indicate the probability that each strategy has the highest net-benefit compared to the other strategies across a range of willingness-to-pay thresholds per disability-adjusted life-year averted by the intervention.

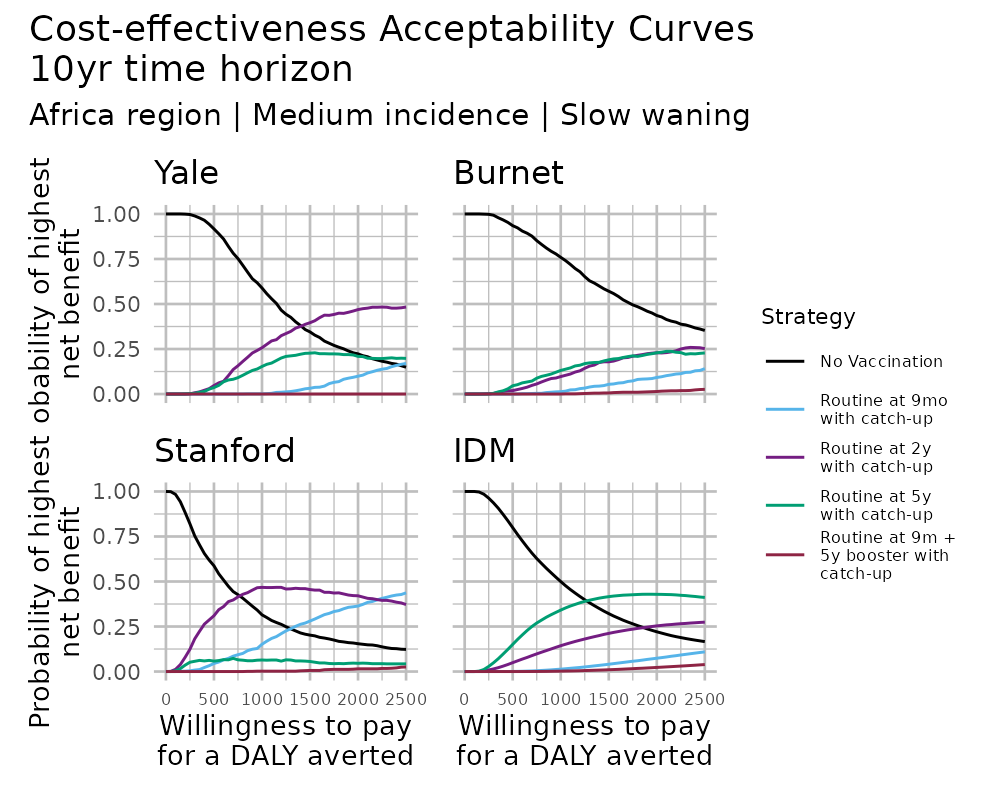

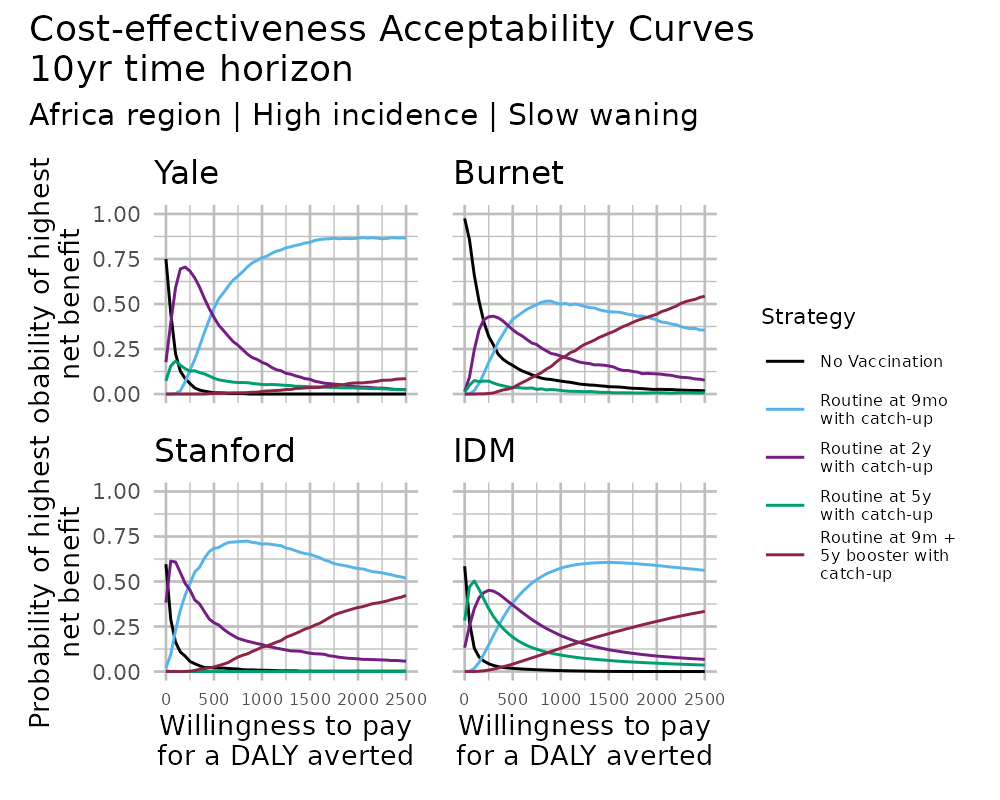

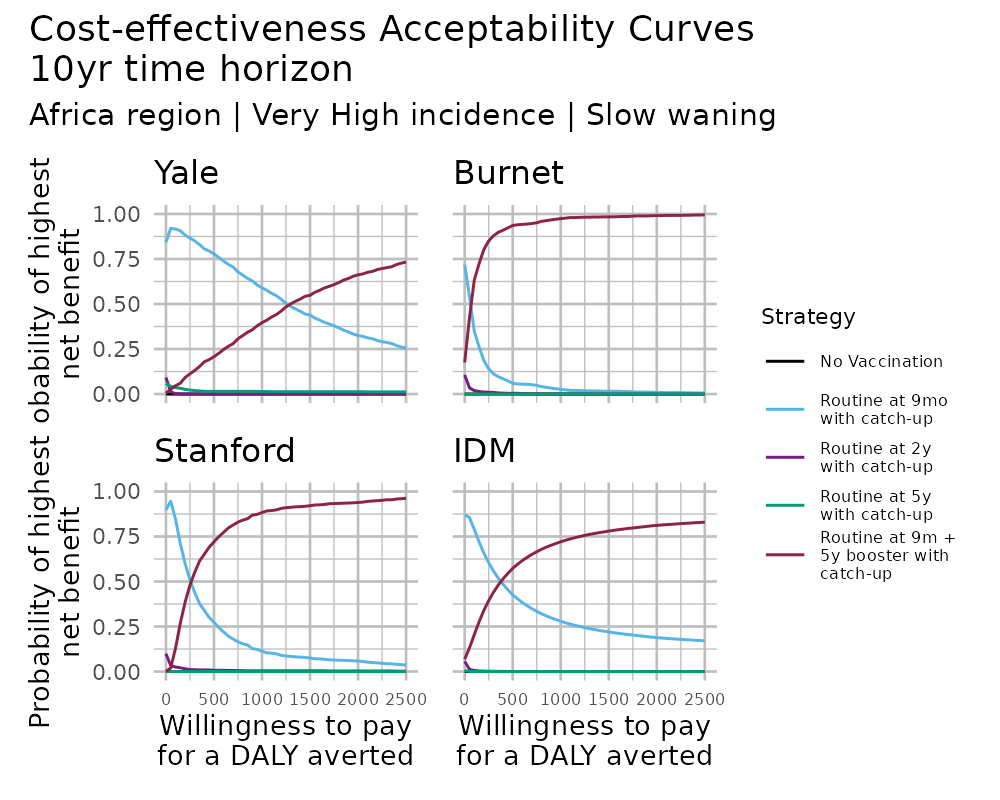

**Figure S7.3.2 Cost-effectiveness acceptability curves (CEACs) for the fast-waning, higher-CFR/higher-cost Africa scenario for all incidence settings over a 10-year time horizon.** The lines indicate the probability that each strategy has the highest net-benefit compared to the other strategies across a range of willingness-to-pay thresholds per disability-adjusted life-year averted by the intervention.

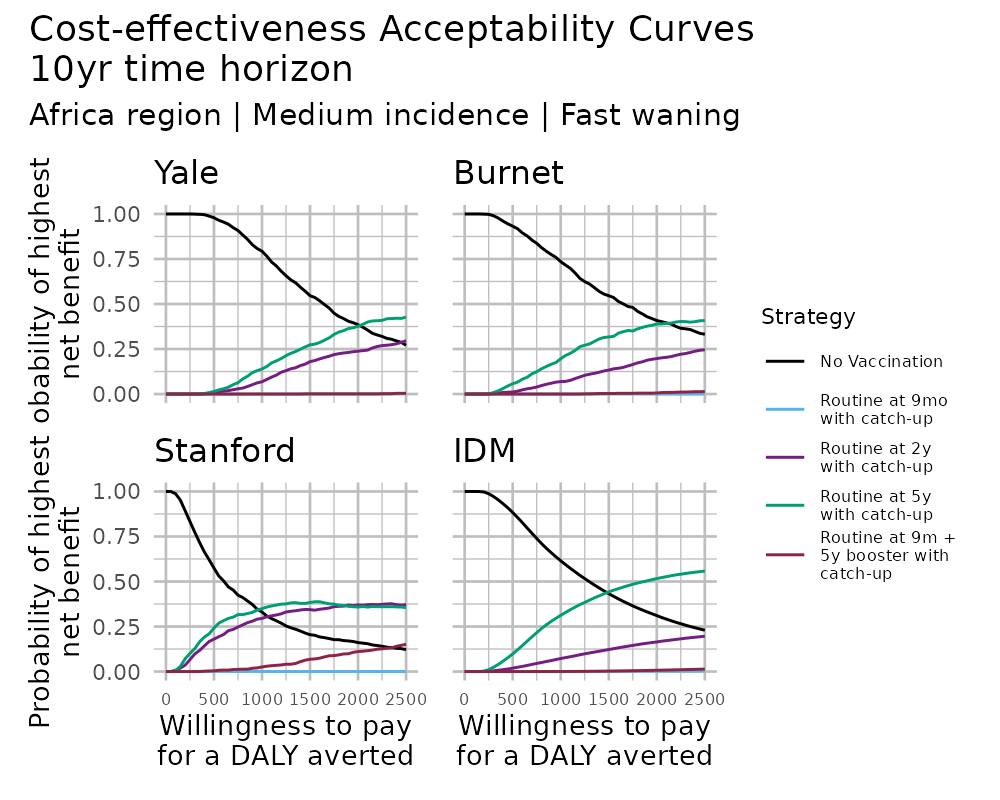

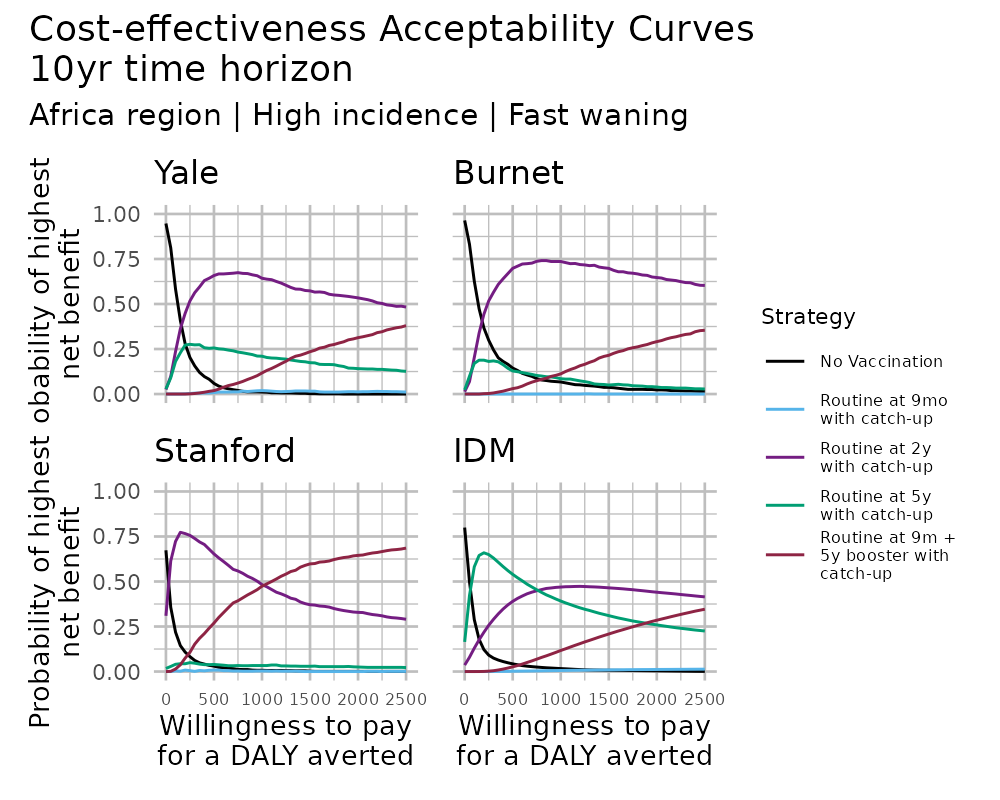

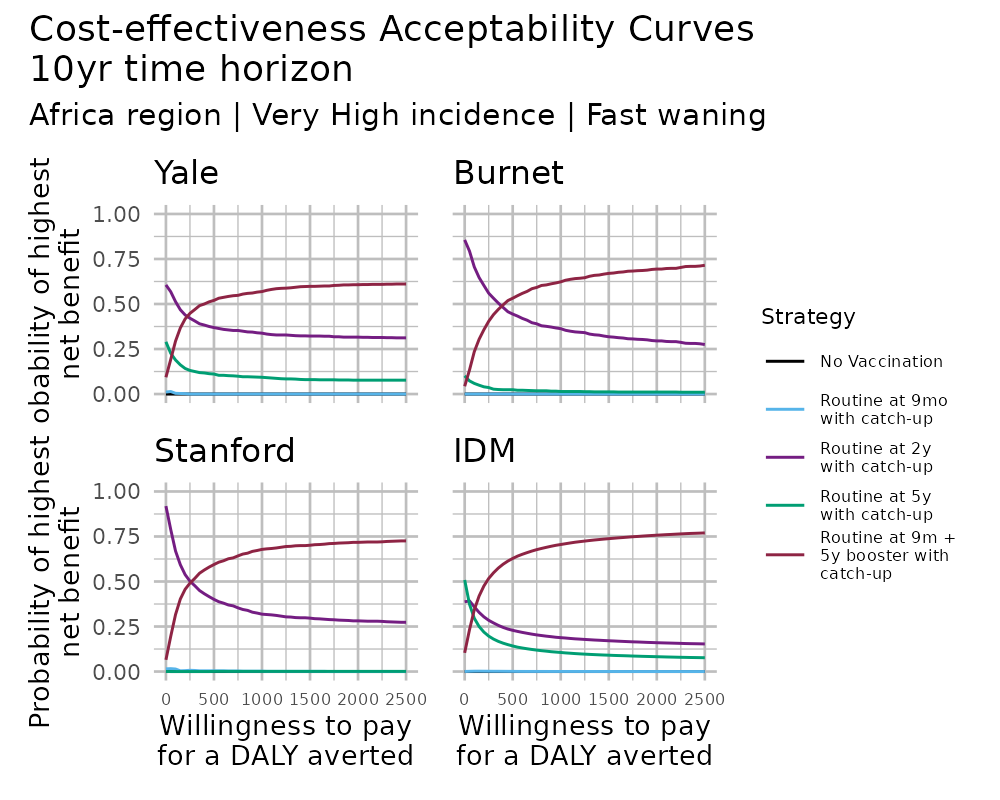

**Figure S7.3.3 Cost-effectiveness acceptability curves (CEACs) for the slow-waning, lower-CFR/lower-cost Asia scenario for all incidence settings over a 10-year time horizon.** The lines indicate the probability that each strategy has the highest net-benefit compared to the other strategies across a range of willingness-to-pay thresholds per disability-adjusted life-year averted by the intervention.

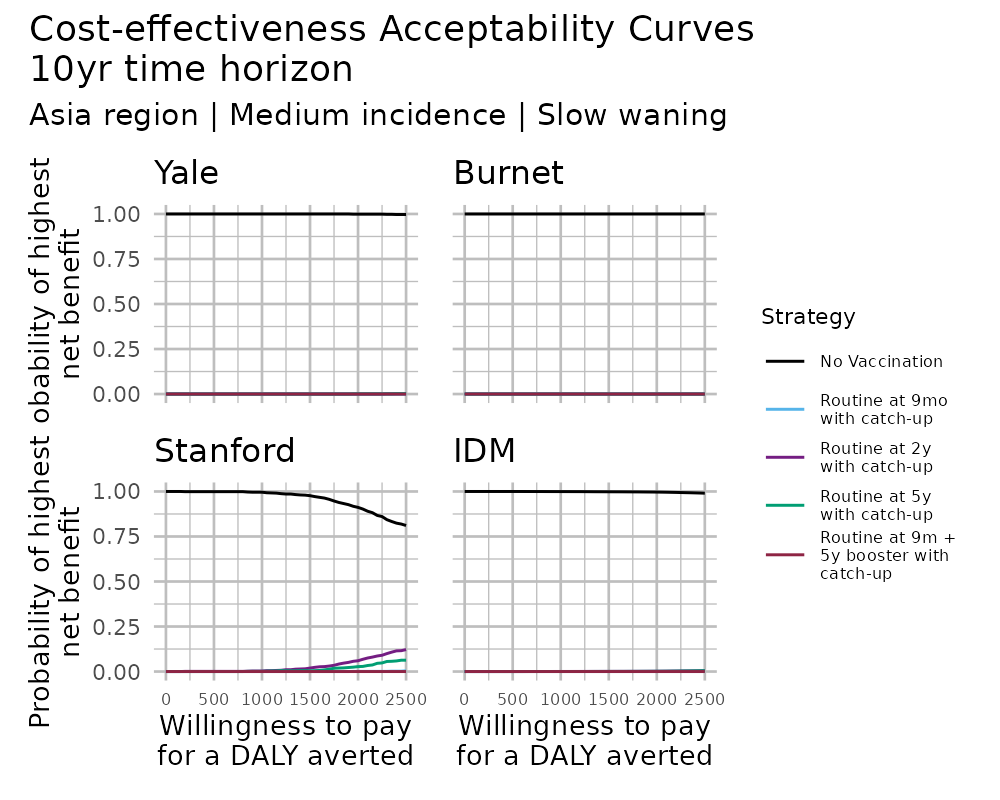

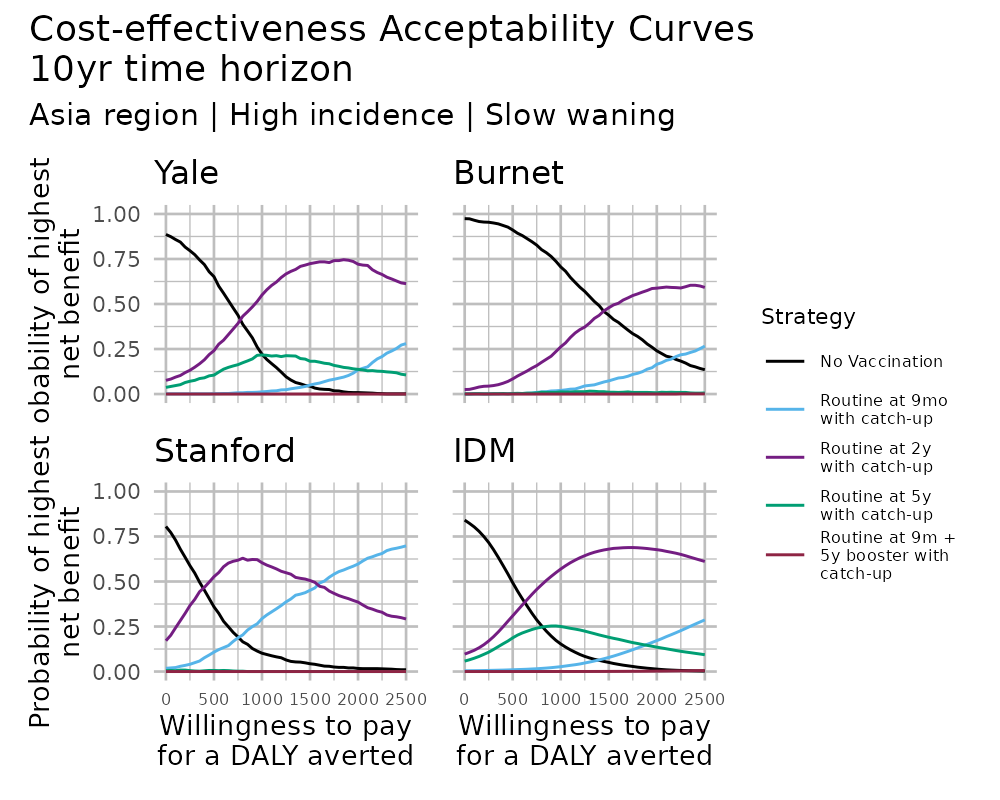

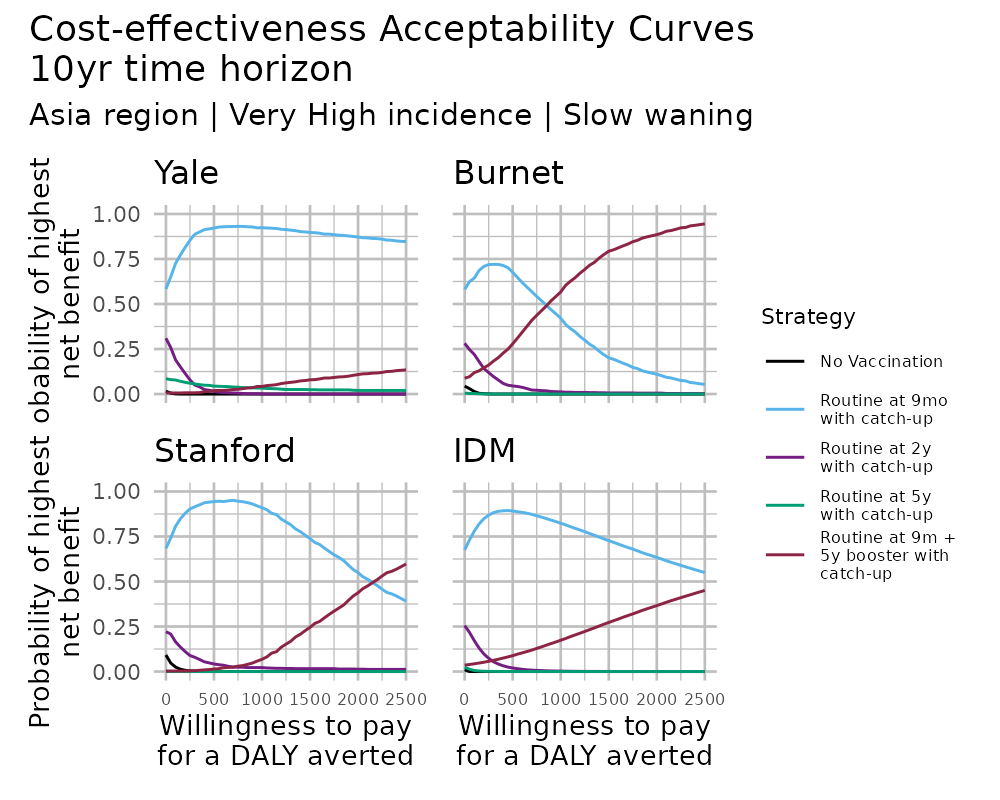

**Figure S7.3.4 Cost-effectiveness acceptability curves (CEACs) for the fast-waning, lower-CFR/lower-cost Asia scenario for all incidence settings over a 10-year time horizon.** The lines indicate the probability that each strategy has the highest net-benefit compared to the other strategies across a range of willingness-to-pay thresholds per disability-adjusted life-year averted by the intervention.

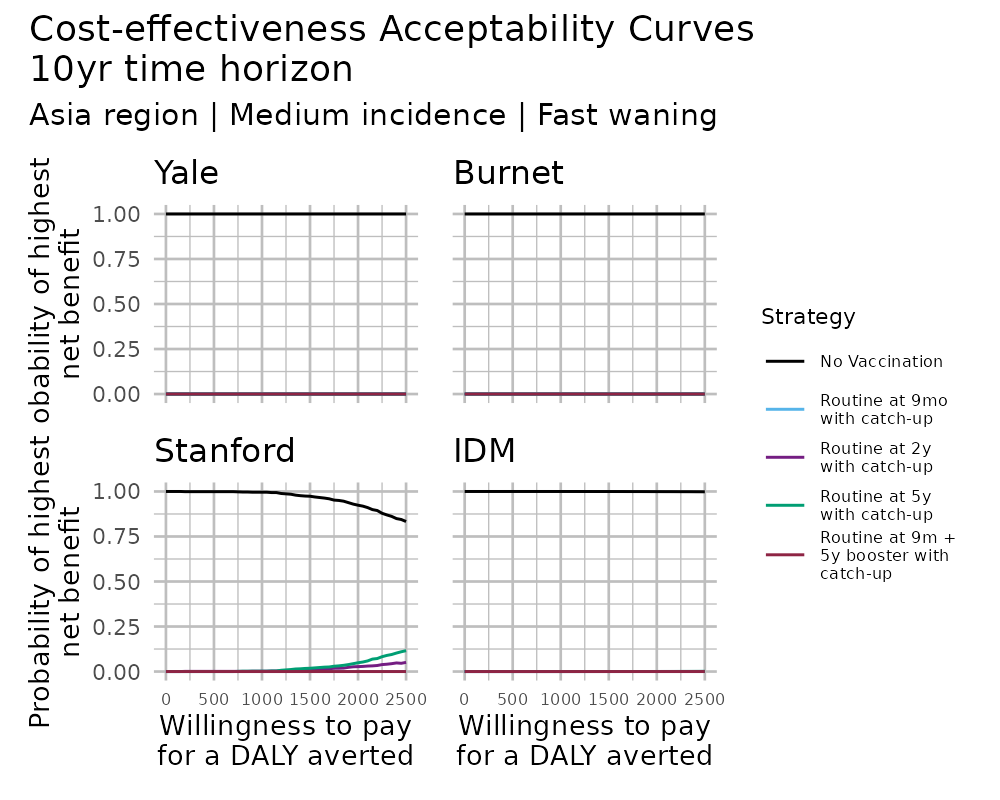

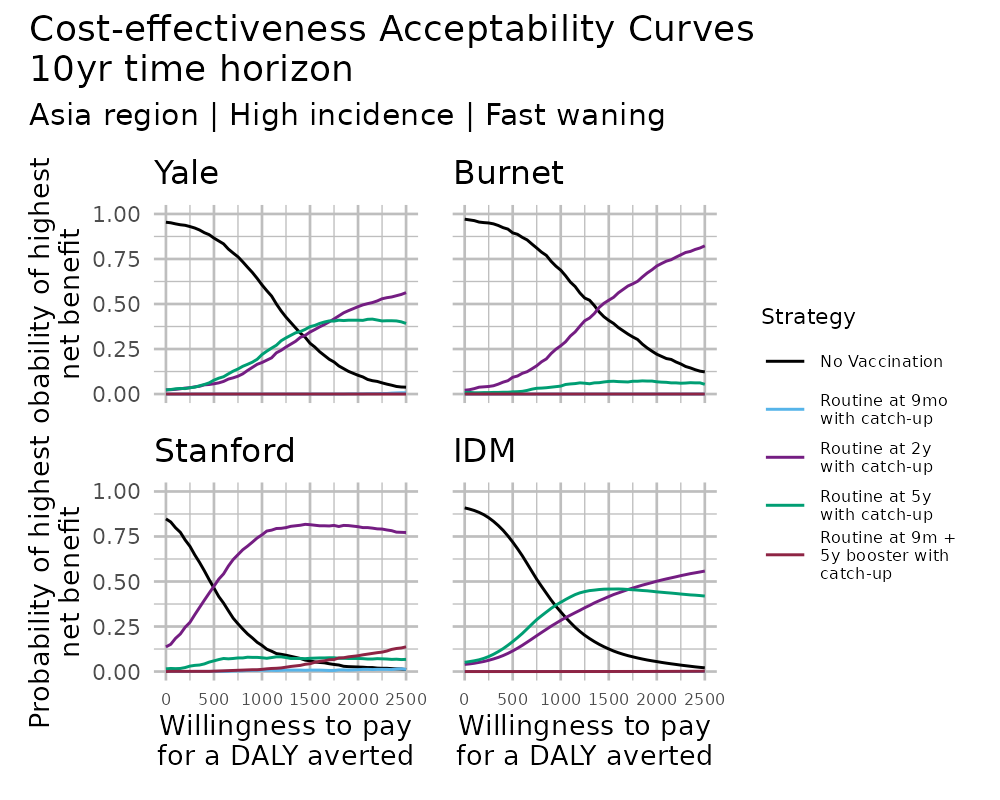

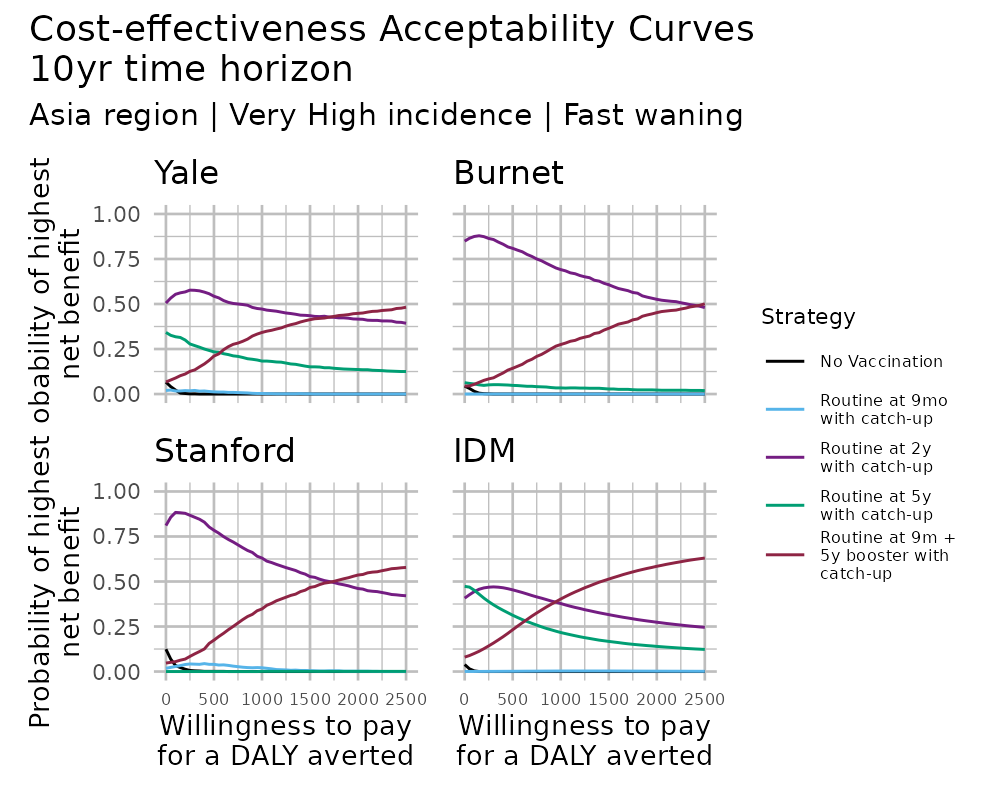

#### 7.4 Cost-effectiveness acceptability frontiers for the 20-year time horizon

**Figure S7.4.1 Cost-effectiveness acceptability frontier (CEAF) heatmap for the slow-waning, higher-CFR/higher-cost Africa scenario for all incidence settings over a 20-year time horizon.** The optimal strategy predicted by each model is indicated by the color plotted at a given willingness-to-pay threshold per disability-adjusted life-year averted by the intervention. The shading indicates the probability that the strategy will provide the highest net benefit, with darker shades representing a higher probability. This analysis includes a two-booster strategy with routine vaccination at 9 months and boosters administered at 5 and 10 years of age.

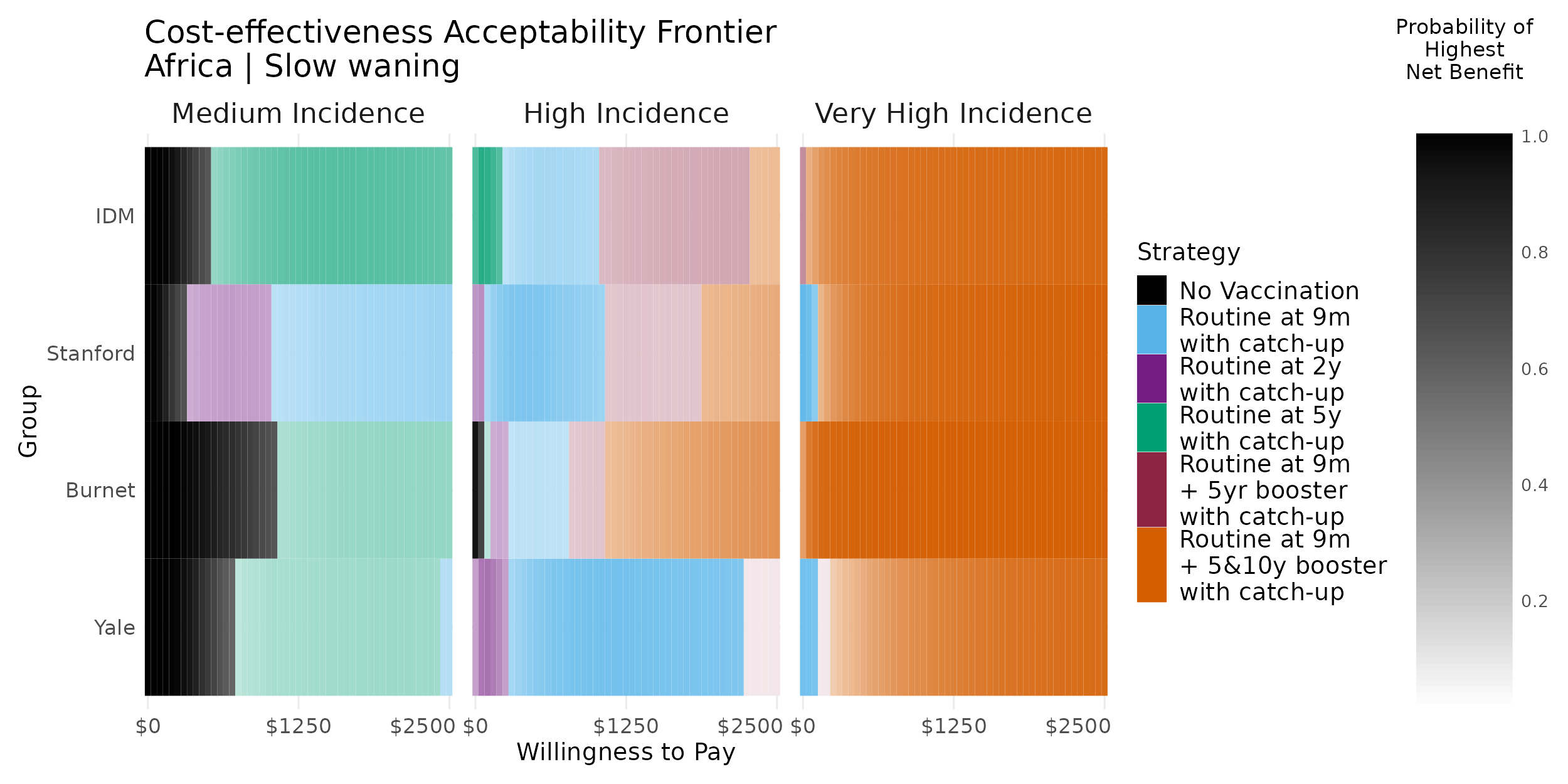

**Figure S7.4.2 Cost-effectiveness acceptability frontier (CEAF) heatmap for the fast-waning, higher-CFR/higher-cost Africa scenario for all incidence settings over a 20-year time horizon.** The optimal strategy predicted by each model is indicated by the color plotted at a given willingness-to-pay threshold per disability-adjusted life-year averted by the intervention. The shading indicates the probability that the strategy will provide the highest net benefit, with darker shades representing a higher probability. This analysis includes a two-booster strategy with routine vaccination at 9 months and boosters administered at 5 and 10 years of age.

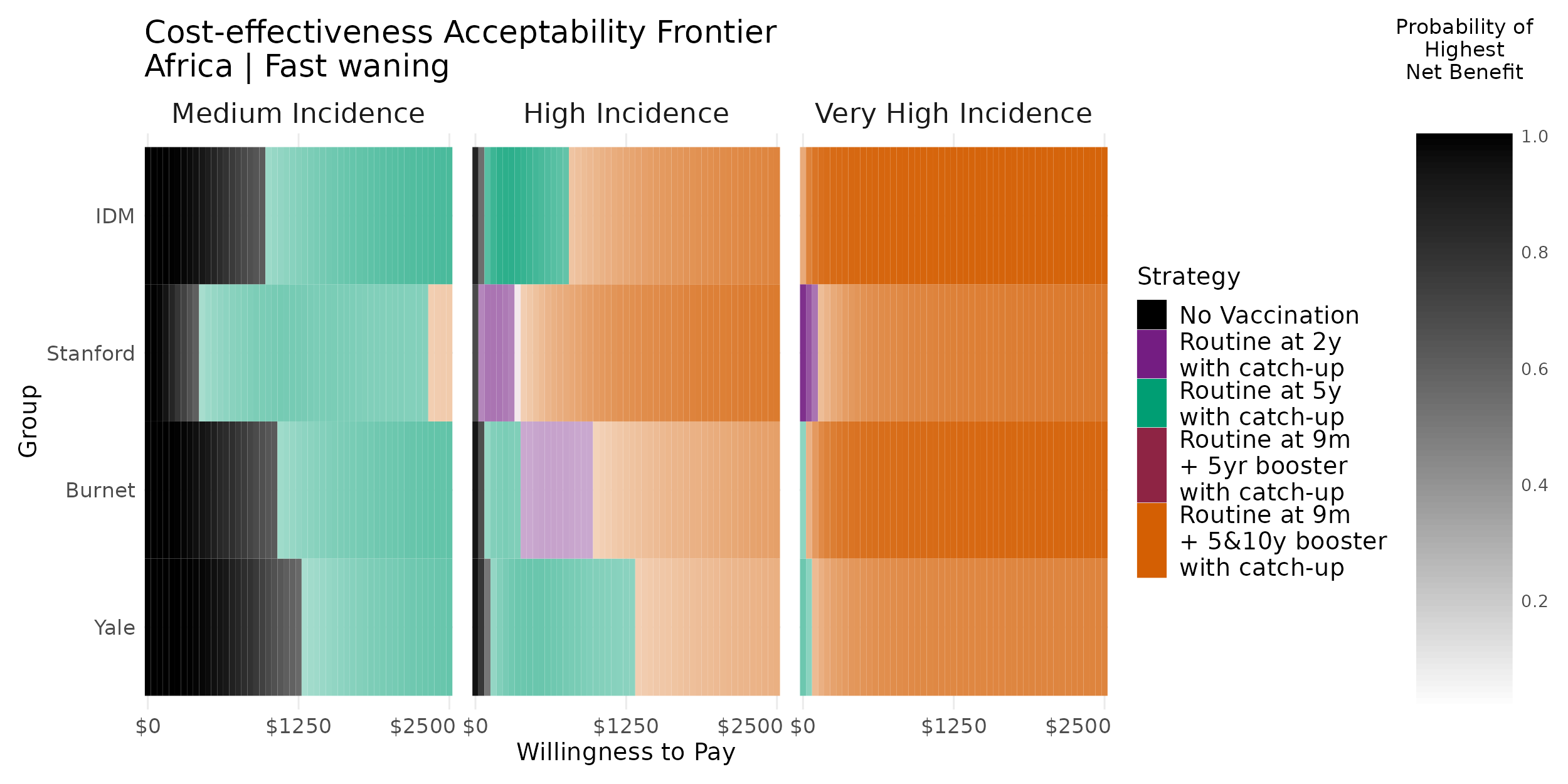

**Figure S7.4.3 Cost-effectiveness acceptability frontier (CEAF) heatmap for the slow-waning, lower-CFR/lower-cost Asia scenario for all incidence settings over a 20-year time horizon.** The optimal strategy predicted by each model is indicated by the color plotted at a given willingness-to-pay threshold per disability-adjusted life-year averted by the intervention. The shading indicates the probability that the strategy will provide the highest net benefit, with darker shades representing a higher probability. This analysis includes a two-booster strategy with routine vaccination at 9 months and boosters administered at 5 and 10 years of age.

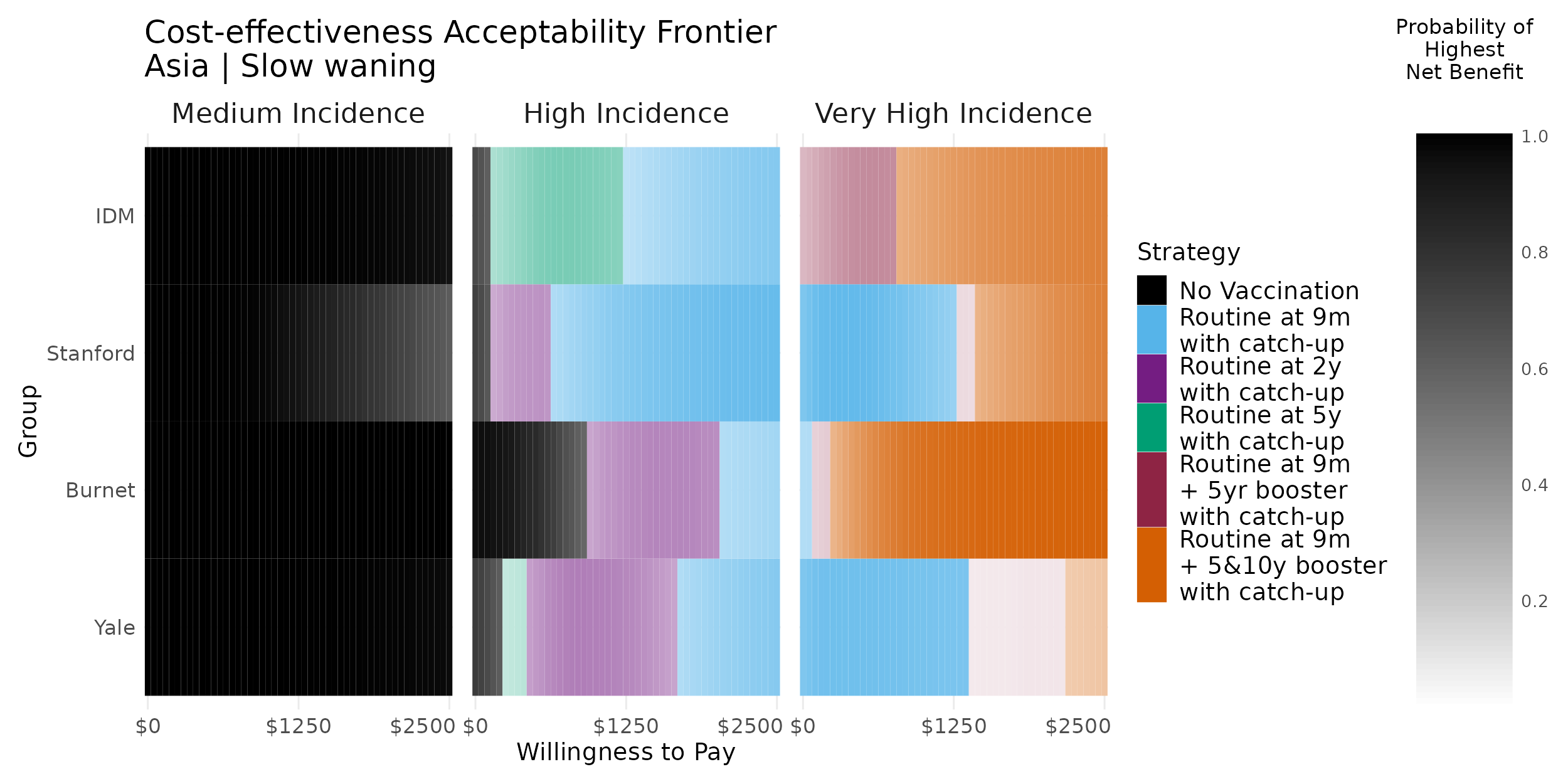

**Figure S7.4.4 Cost-effectiveness acceptability frontier (CEAF) heatmap for the fast-waning, lower-CFR/lower-cost Asia scenario for all incidence settings over a 20-year time horizon.** The optimal strategy predicted by each model is indicated by the color plotted at a given willingness-to-pay threshold per disability-adjusted life-year averted by the intervention. The shading indicates the probability that the strategy will provide the highest net benefit, with darker shades representing a higher probability. This analysis includes a two-booster strategy with routine vaccination at 9 months and boosters administered at 5 and 10 years of age.

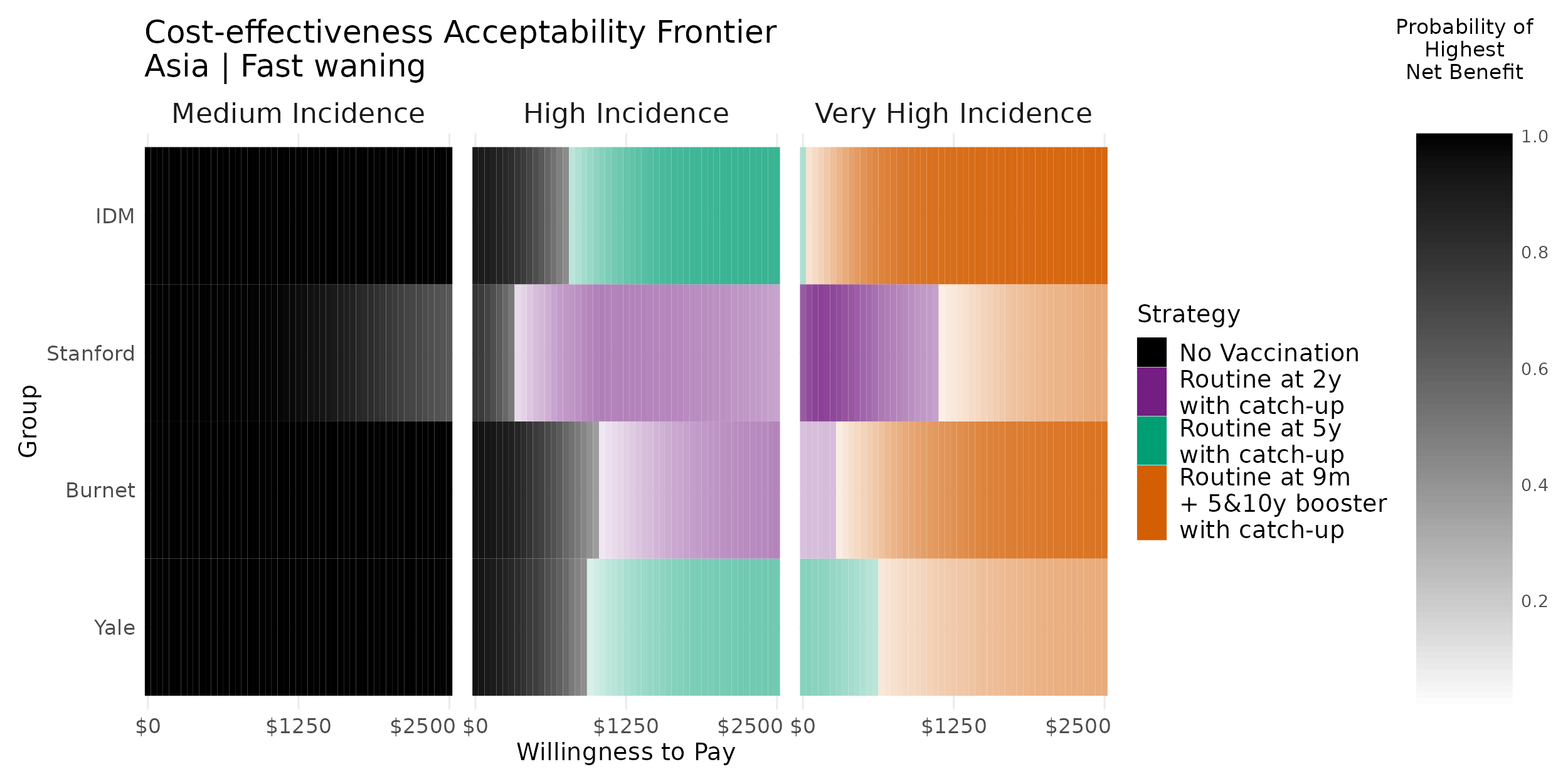

#### 7.5 Scenario analysis: youngest age of vaccination at 15 months

**Figure S7.5.1 Cost-effectiveness acceptability frontier (CEAF) heatmap considering the youngest age of vaccination at 15 months for the slow-waning, higher-CFR/higher-cost Africa scenario for all incidence settings over a 10-year time horizon.** The optimal strategy predicted by each model is indicated by the color plotted at a given willingness-to-pay threshold per disability-adjusted life-year averted by the intervention. The shading indicates the probability that the strategy will provide the highest net benefit, with darker shades representing a higher probability.

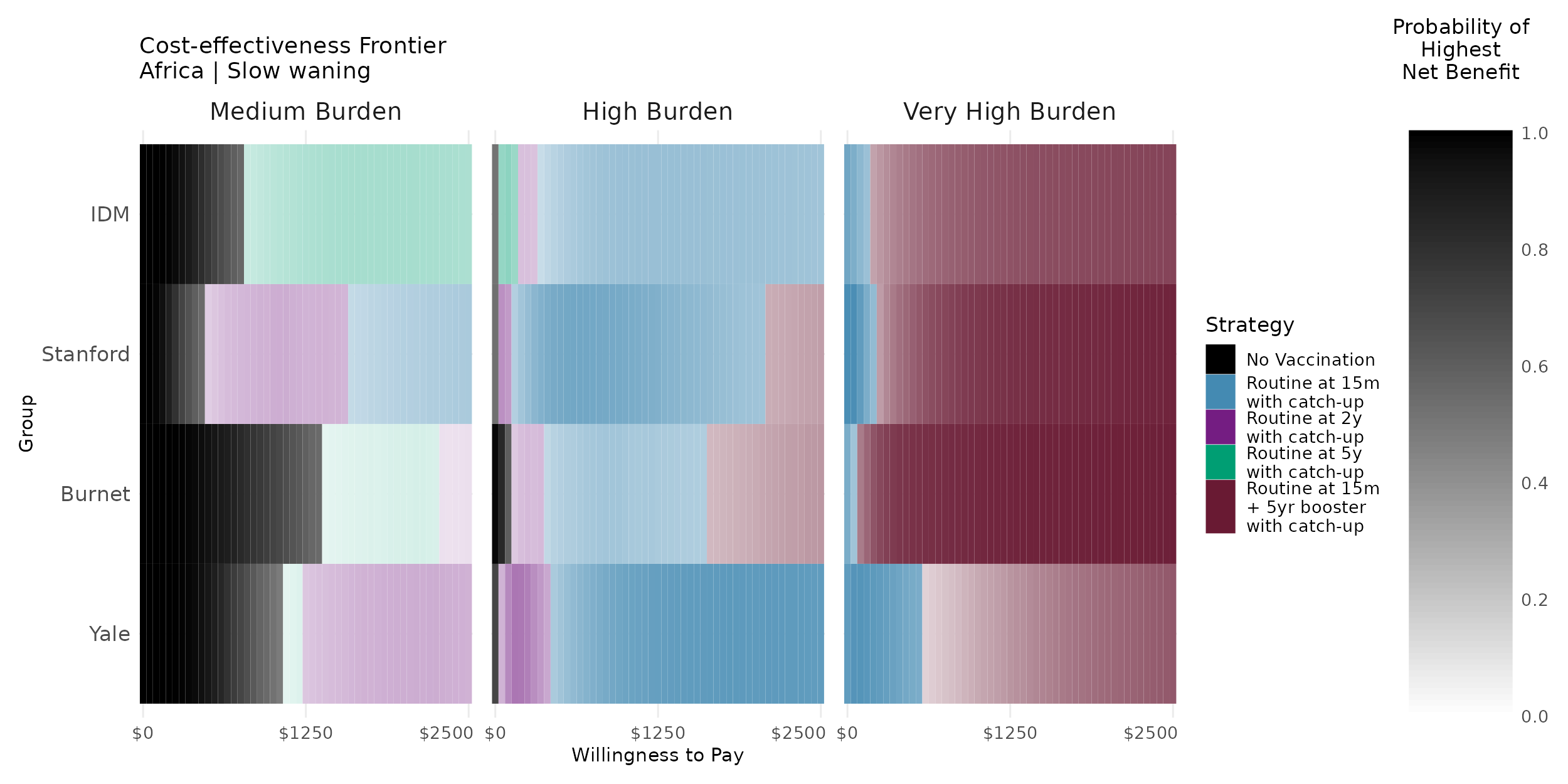

**Figure S7.5.2 Cost-effectiveness acceptability frontier (CEAF) heatmap considering the youngest age of vaccination at 15 months for the fast-waning, higher-CFR/higher-cost Africa scenario for all incidence settings over a 10-year time horizon.** The optimal strategy predicted by each model is indicated by the color plotted at a given willingness-to-pay threshold per disability-adjusted life-year averted by the intervention. The shading indicates the probability that the strategy will provide the highest net benefit, with darker shades representing a higher probability.

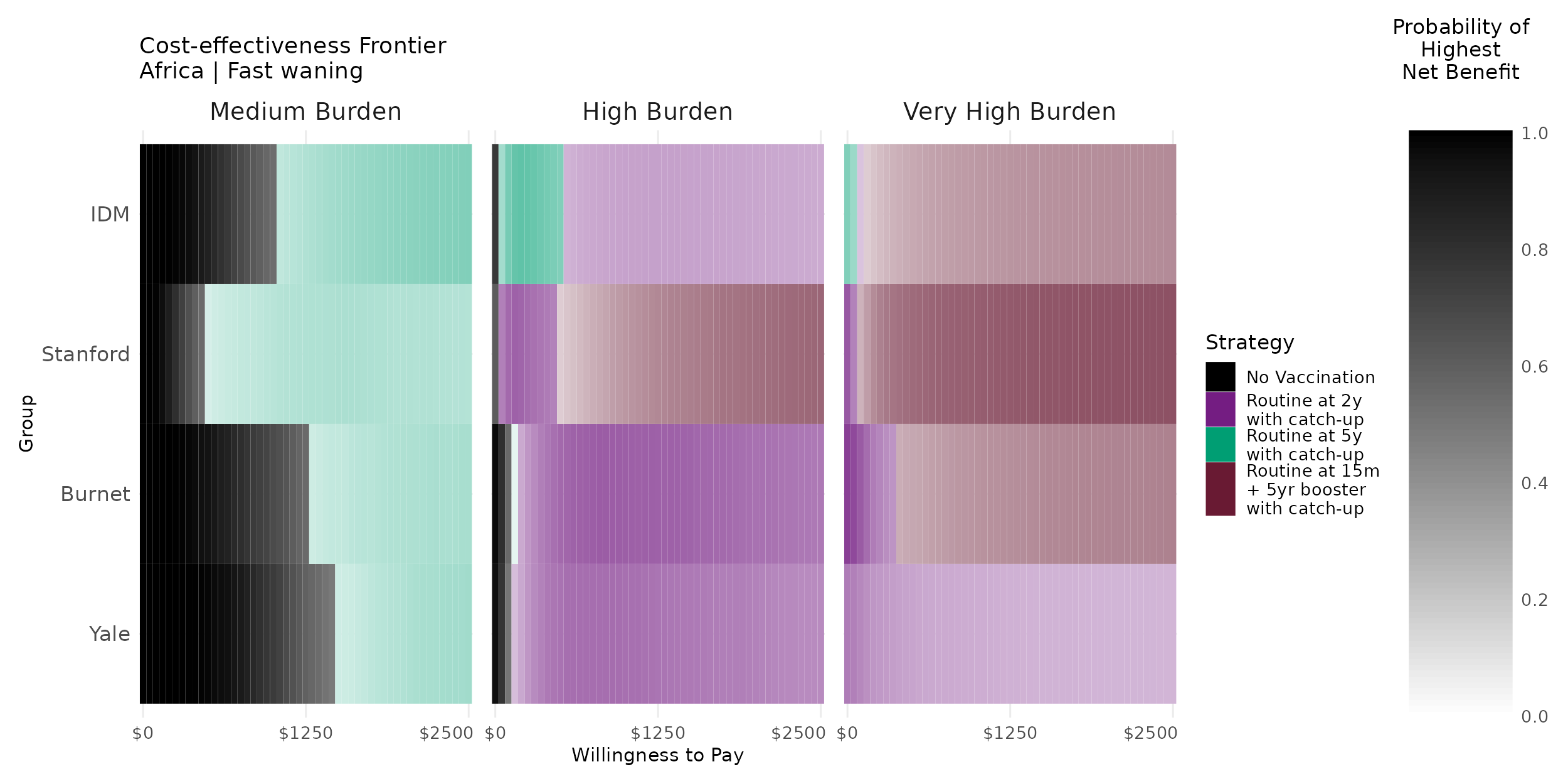

**Figure S7.5.3 Cost-effectiveness acceptability frontier (CEAF) heatmap considering the youngest age of vaccination at 15 months for the slow-waning, lower-CFR/lower-cost Asia scenario for all incidence settings over a 10-year time horizon.** The optimal strategy predicted by each model is indicated by the color plotted at a given willingness-to-pay threshold per disability-adjusted life-year averted by the intervention. The shading indicates the probability that the strategy will provide the highest net benefit, with darker shades representing a higher probability.

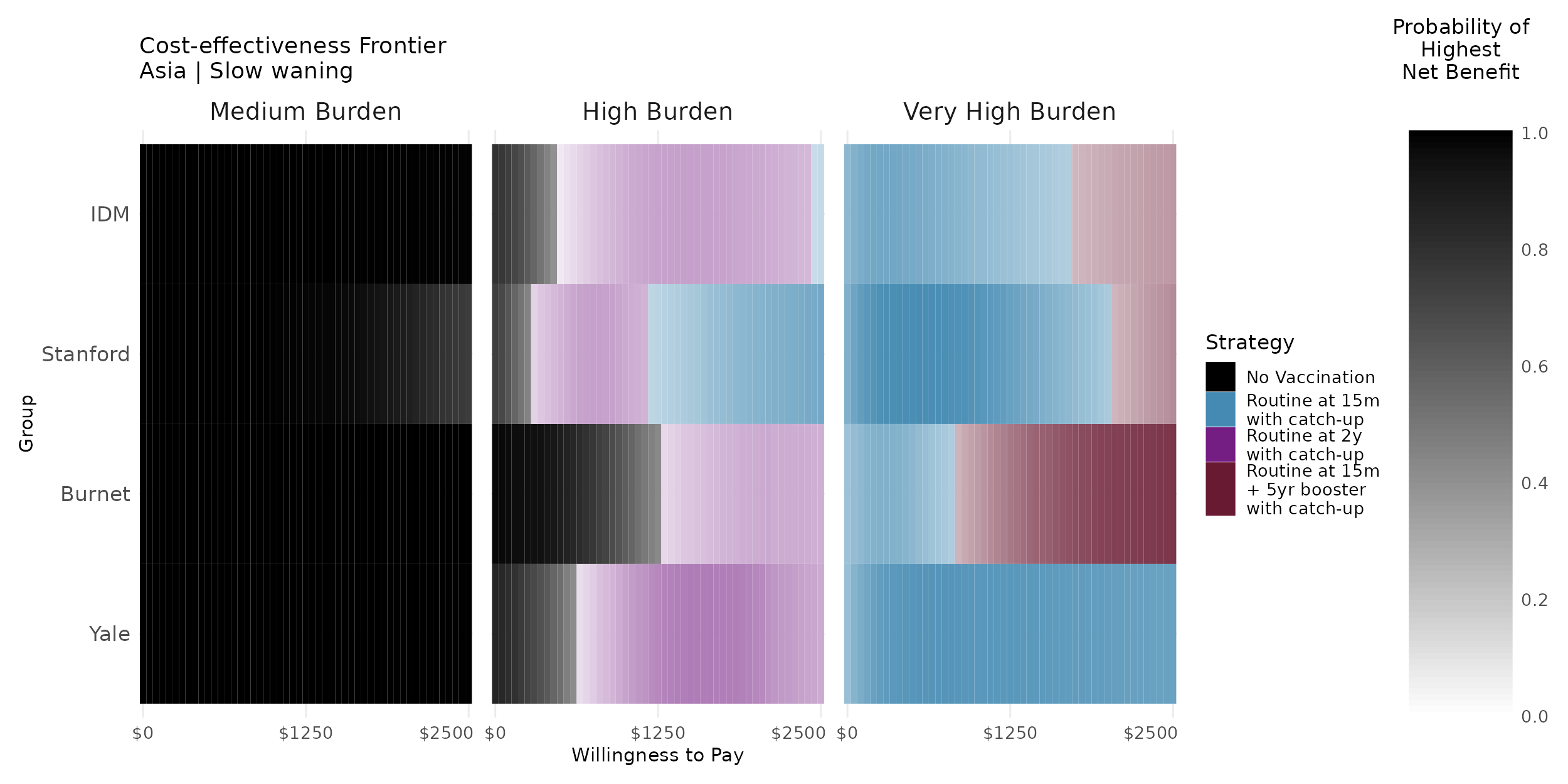

**Figure S7.5.4 Cost-effectiveness acceptability frontier (CEAF) heatmap considering the youngest age of vaccination at 15 months for the fast-waning, lower-CFR/lower-cost Asia scenario for all incidence settings over a 10-year time horizon.** The optimal strategy predicted by each model is indicated by the color plotted at a given willingness-to-pay threshold per disability-adjusted life-year averted by the intervention. The shading indicates the probability that the strategy will provide the highest net benefit, with darker shades representing a higher probability.

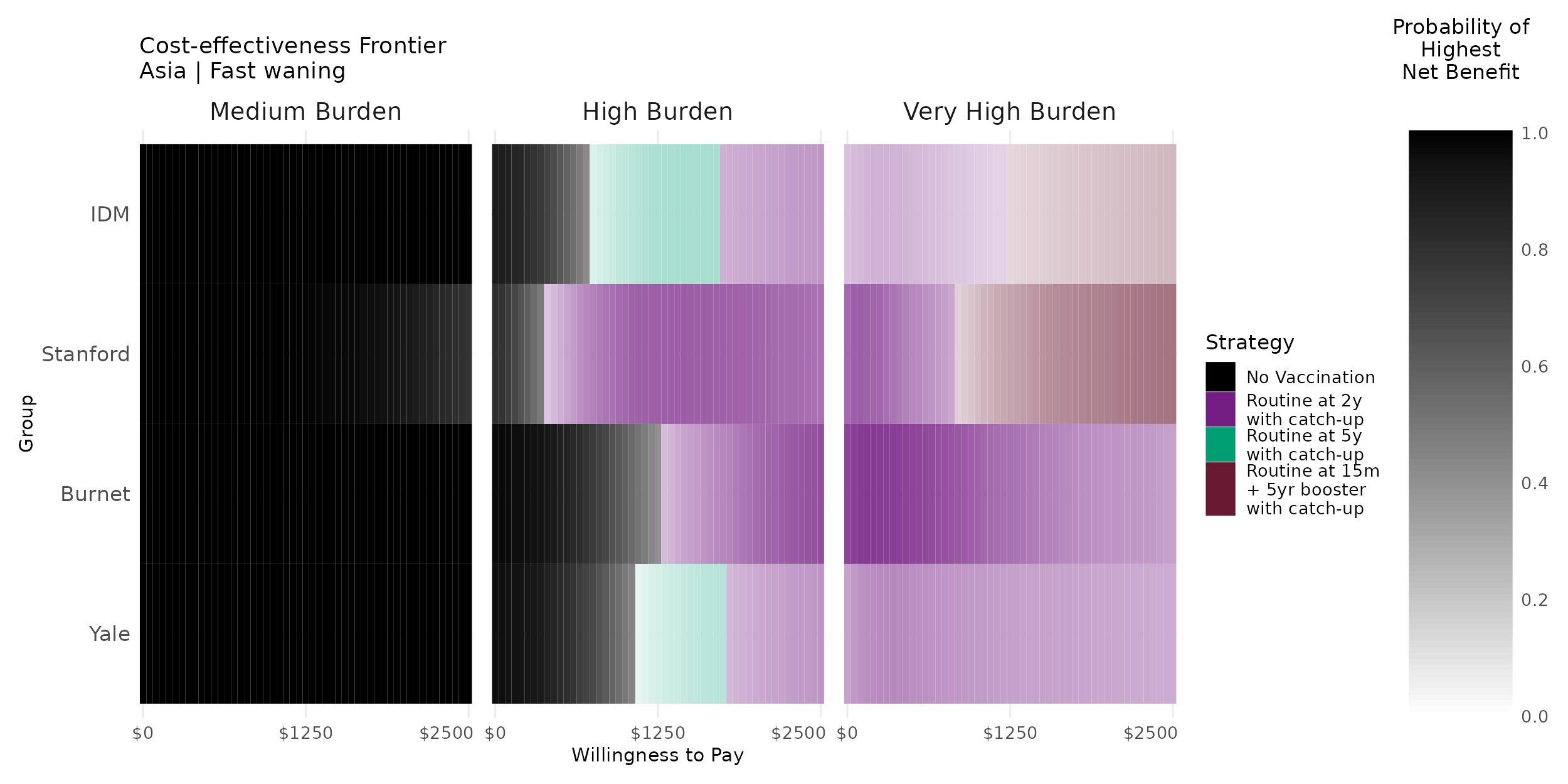

#### 7.6 Scenario analysis: vaccination without catch-up campaigns

**Figure S7.6.1 Cost-effectiveness acceptability frontier (CEAF) heatmap considering strategies without catch-up campaigns only for the slow-waning, higher-CFR/higher-cost Africa scenario for all incidence settings over a 10-year time horizon.** The optimal strategy predicted by each model is indicated by the color plotted at a given willingness-to-pay threshold per disability-adjusted life-year averted by the intervention. The shading indicates the probability that the strategy will provide the highest net benefit, with darker shades representing a higher probability.

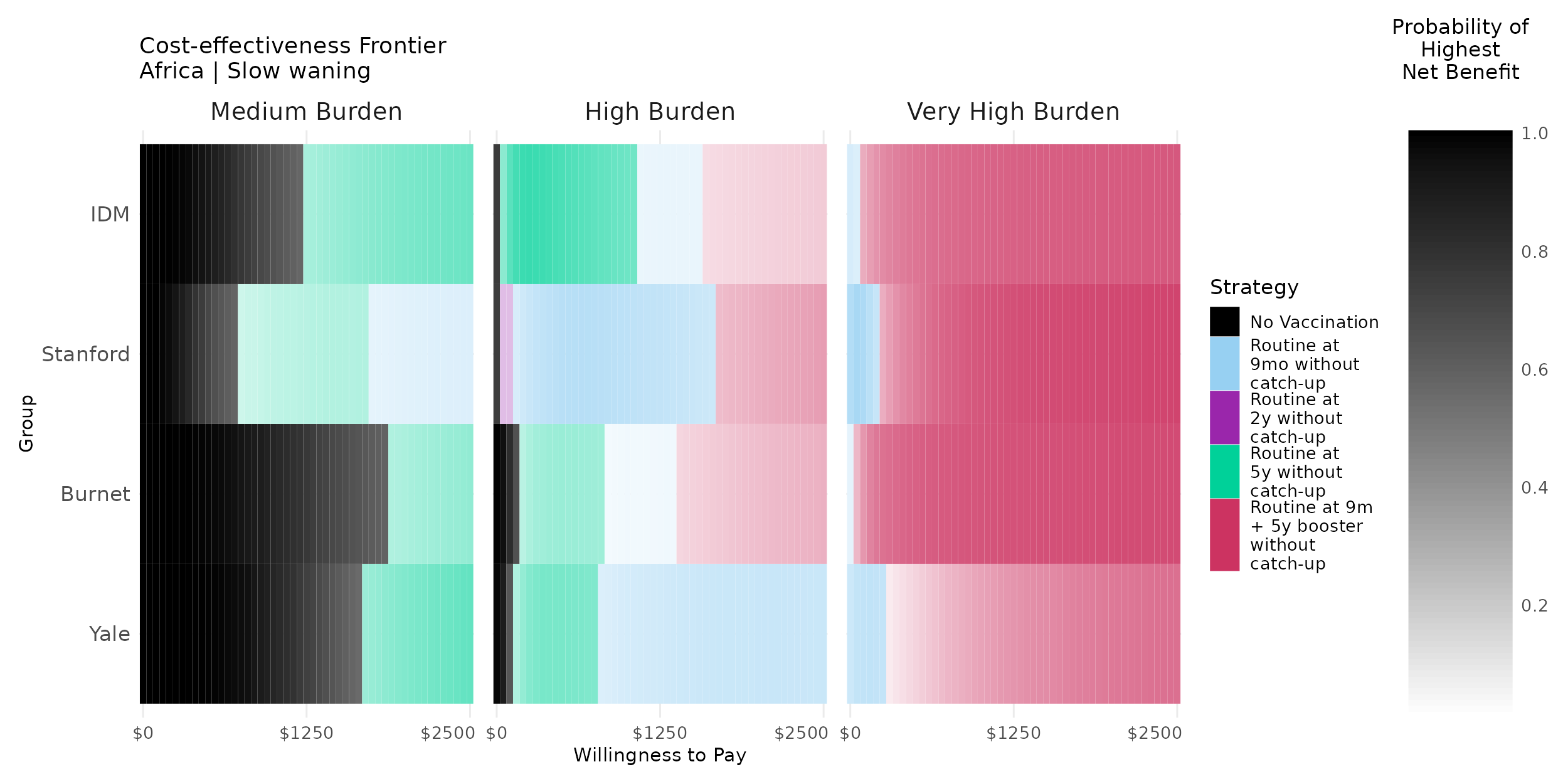

**Figure S7.6.2 Cost-effectiveness acceptability frontier (CEAF) heatmap considering strategies without catch-up campaigns only for the fast-waning, higher-CFR/higher-cost Africa scenario for all incidence settings over a 10-year time horizon.** The optimal strategy predicted by each model is indicated by the color plotted at a given willingness-to-pay threshold per disability-adjusted life-year averted by the intervention. The shading indicates the probability that the strategy will provide the highest net benefit, with darker shades representing a higher probability.

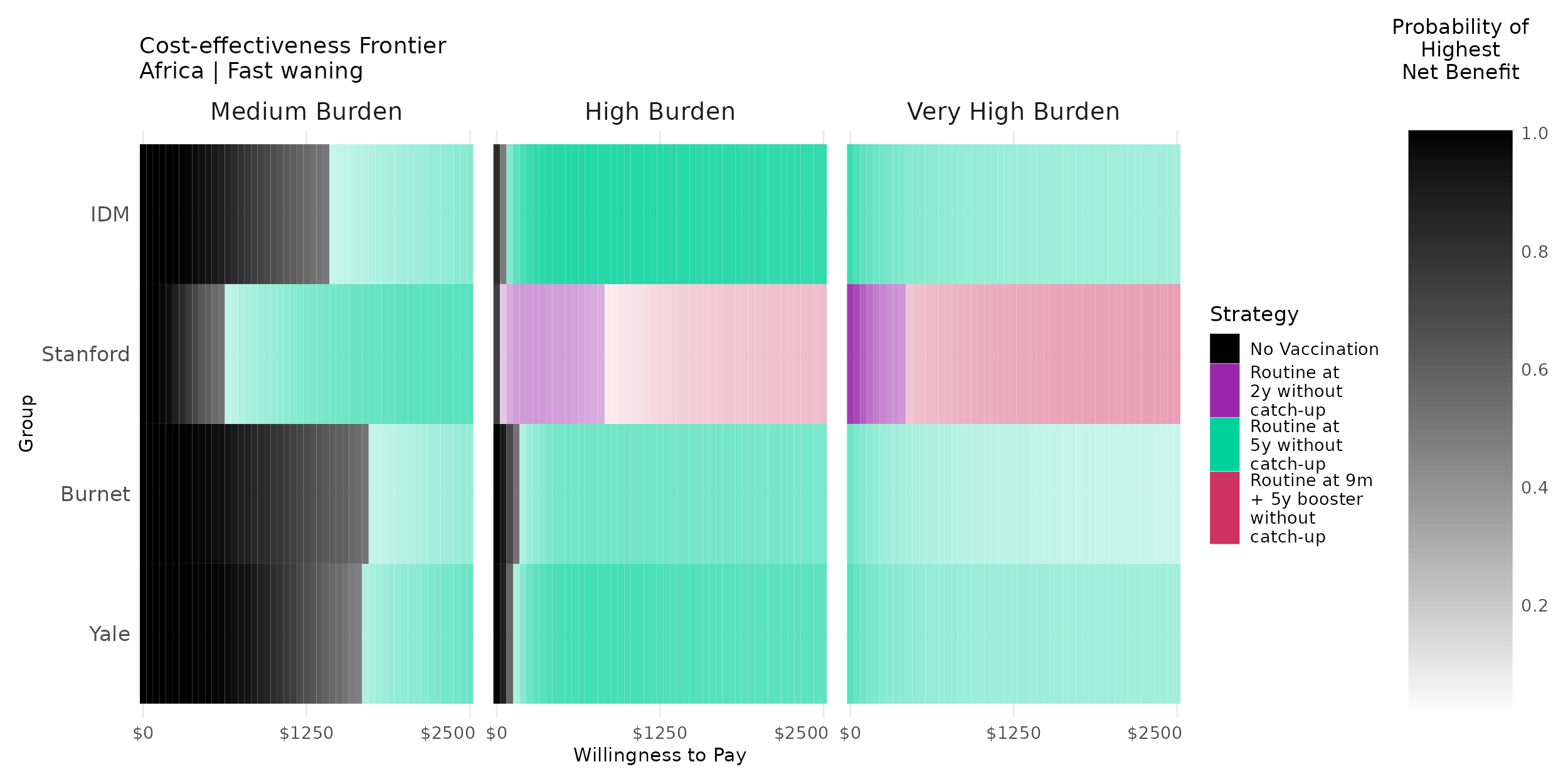

**Figure S7.6.3 Cost-effectiveness acceptability frontier (CEAF) heatmap considering strategies without catch-up campaigns only for the slow-waning, lower-CFR/lower-cost Asia scenario for all incidence settings over a 10-year time horizon.** The optimal strategy predicted by each model is indicated by the color plotted at a given willingness-to-pay threshold per disability-adjusted life-year averted by the intervention. The shading indicates the probability that the strategy will provide the highest net benefit, with darker shades representing a higher probability.

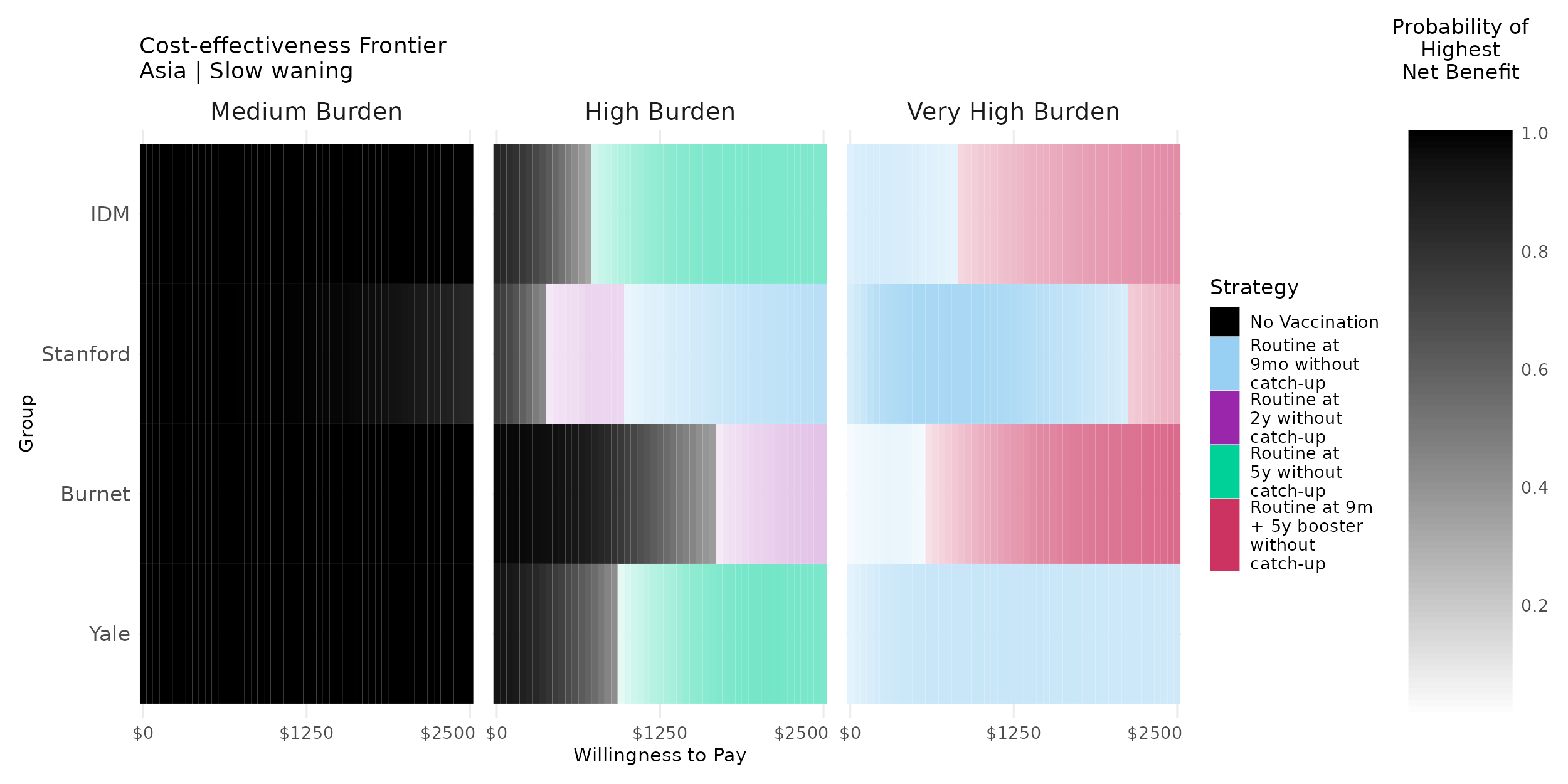

**Figure S7.6.4 Cost-effectiveness acceptability frontier (CEAF) heatmap considering strategies without catch-up campaigns only for the fast-waning, lower-CFR/lower-cost Asia scenario for all incidence settings over a 10-year time horizon.** The optimal strategy predicted by each model is indicated by the color plotted at a given willingness-to-pay threshold per disability-adjusted life-year averted by the intervention. The shading indicates the probability that the strategy will provide the highest net benefit, with darker shades representing a higher probability.

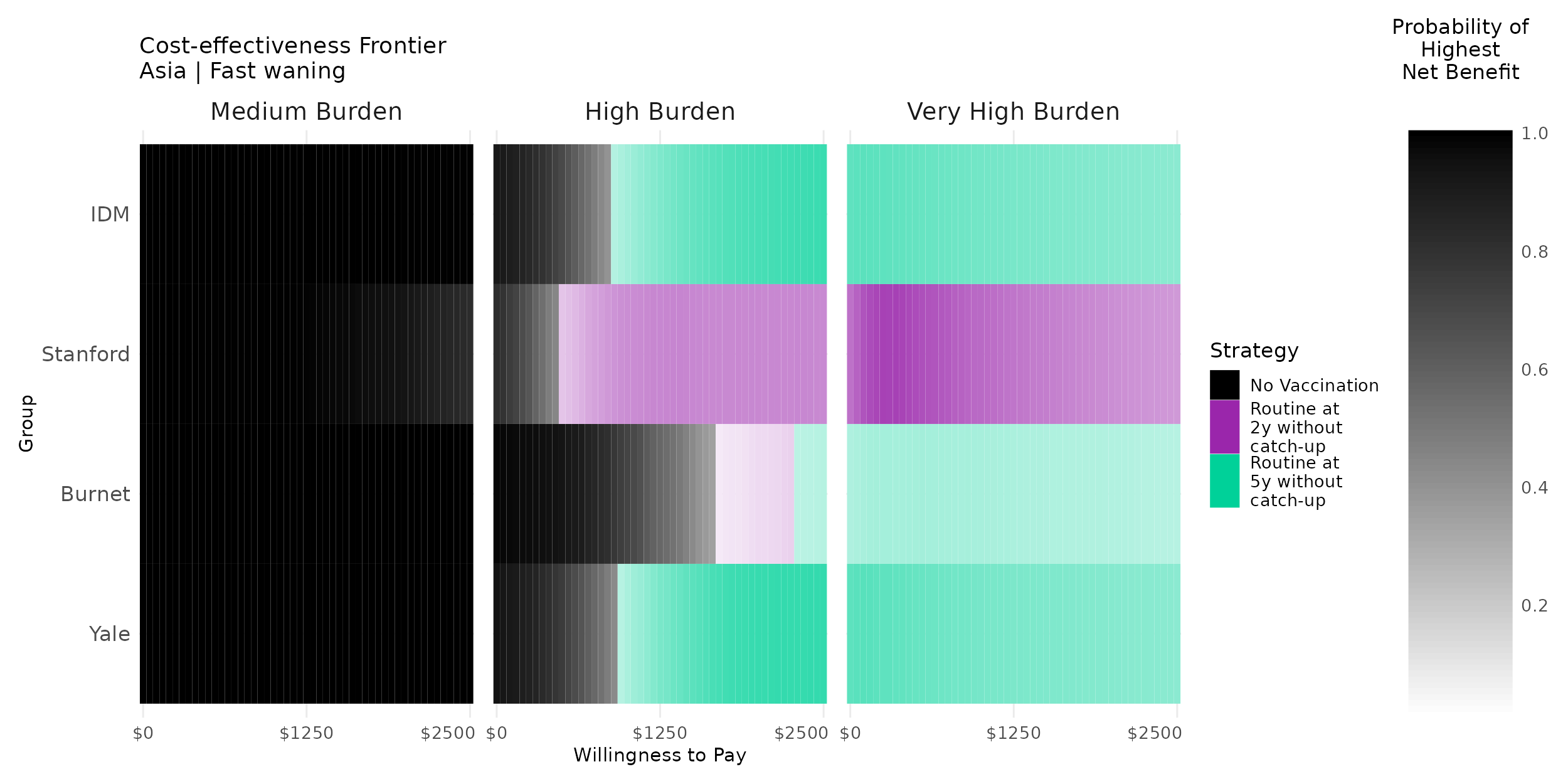

#### 7.7 Threshold analysis

**Figure S7.7.1 Threshold analysis of the preferred vaccination strategy across different baseline incidence values with peak incidence in the 10- to 15-year age group.** The optimal strategy predicted by the model (from the cost-effectiveness acceptability frontier) is indicated by the color plotted for different baseline typhoid incidence rates (from 10 to 500 cases per 100,000 person-years) at a given willingness-to-pay per disability-adjusted life-year averted (from $0 to $5,000, x-axis). Results are plotted for a 10-year time horizon under the slow-waning scenario (top) and fast-waning scenario (bottom) from the higher-CFR/higher-cost African setting (left) and lower-CFR/lower-cost Asian setting (right). The red lines indicate the calibrated incidence used in the main analysis for medium incidence settings. These plots present the results for the Yale model; results for all models are included below.

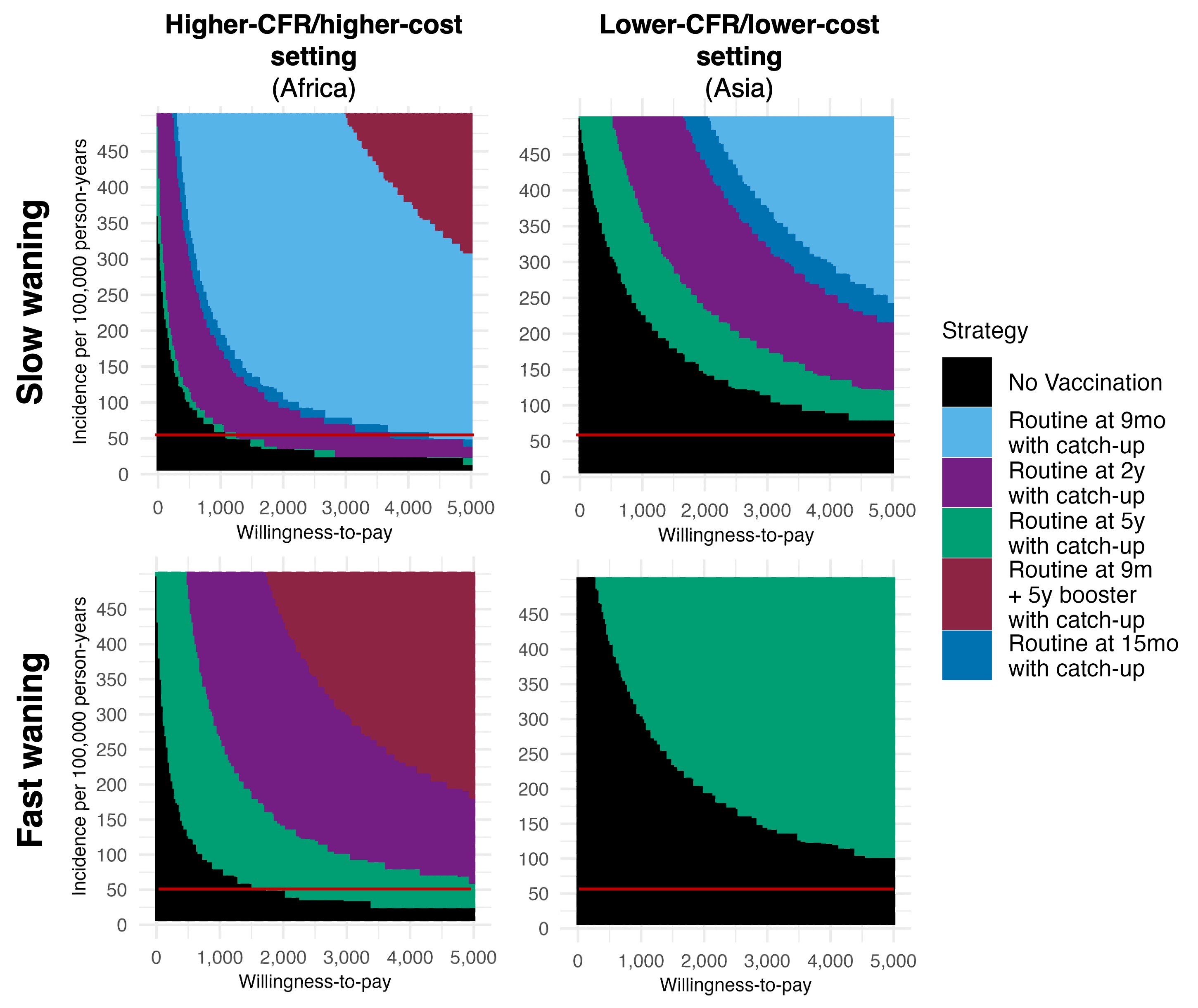

**Figure S7.7.2 Cost-effectiveness acceptability frontier across willingness-to-pay thresholds per disability-adjusted life years averted and incidence ranges for the slow-waning, higher-CFR/higher-cost Africa setting over a 10-year time horizon.** The optimal strategy predicted by each model is indicated by the color plotted at a given willingness-to-pay threshold per disability-adjusted life-year averted by the intervention.

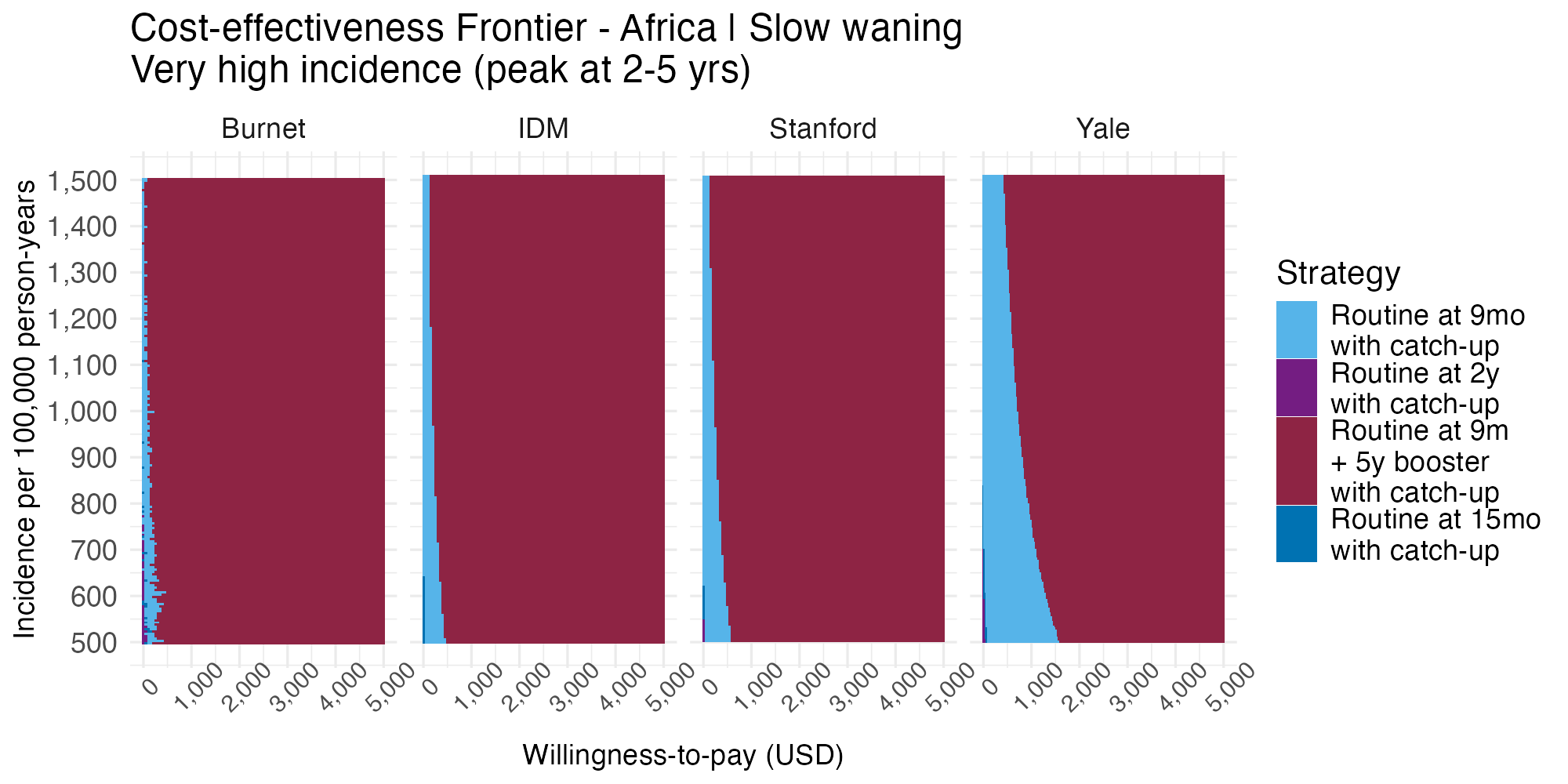

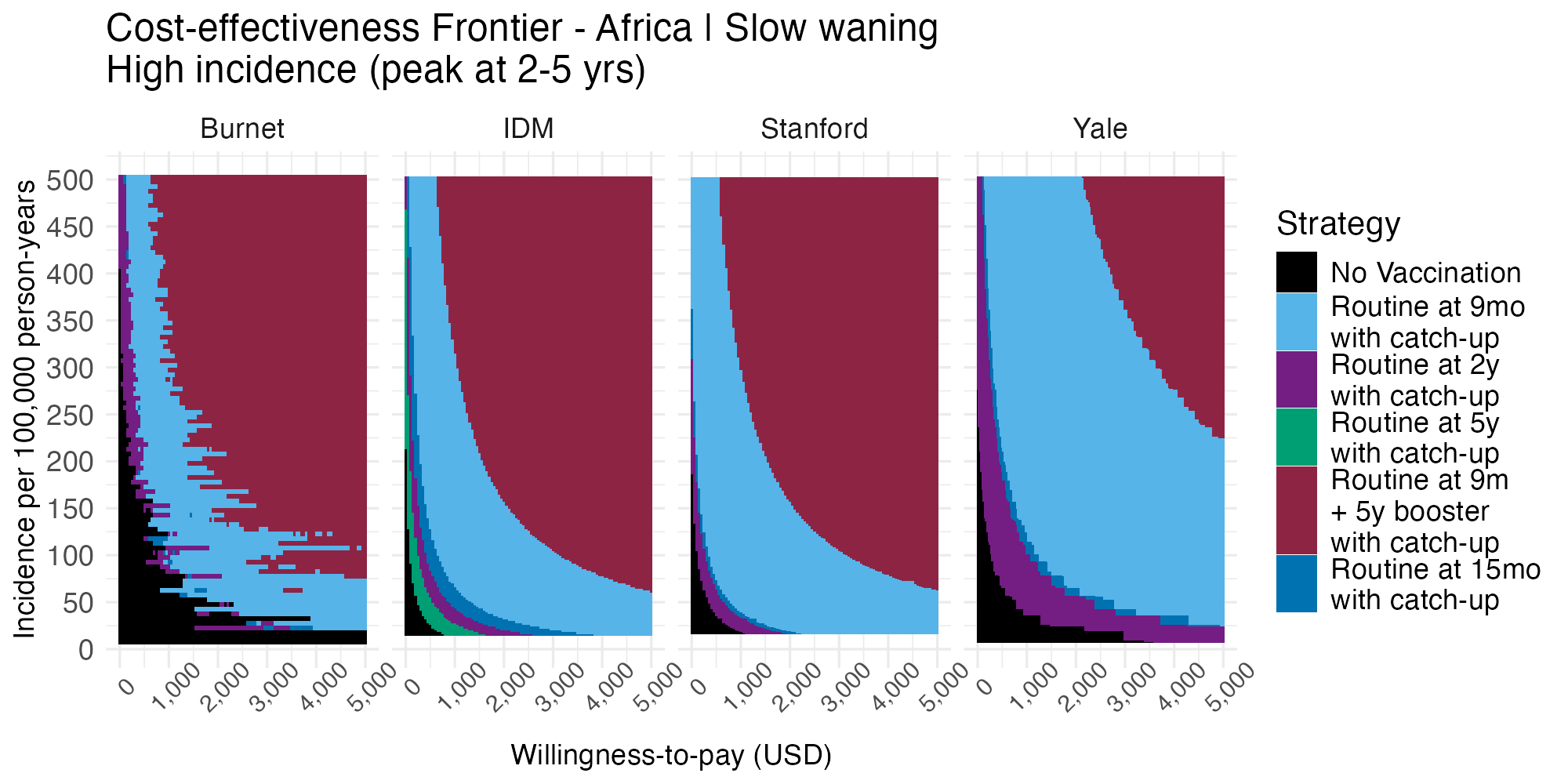

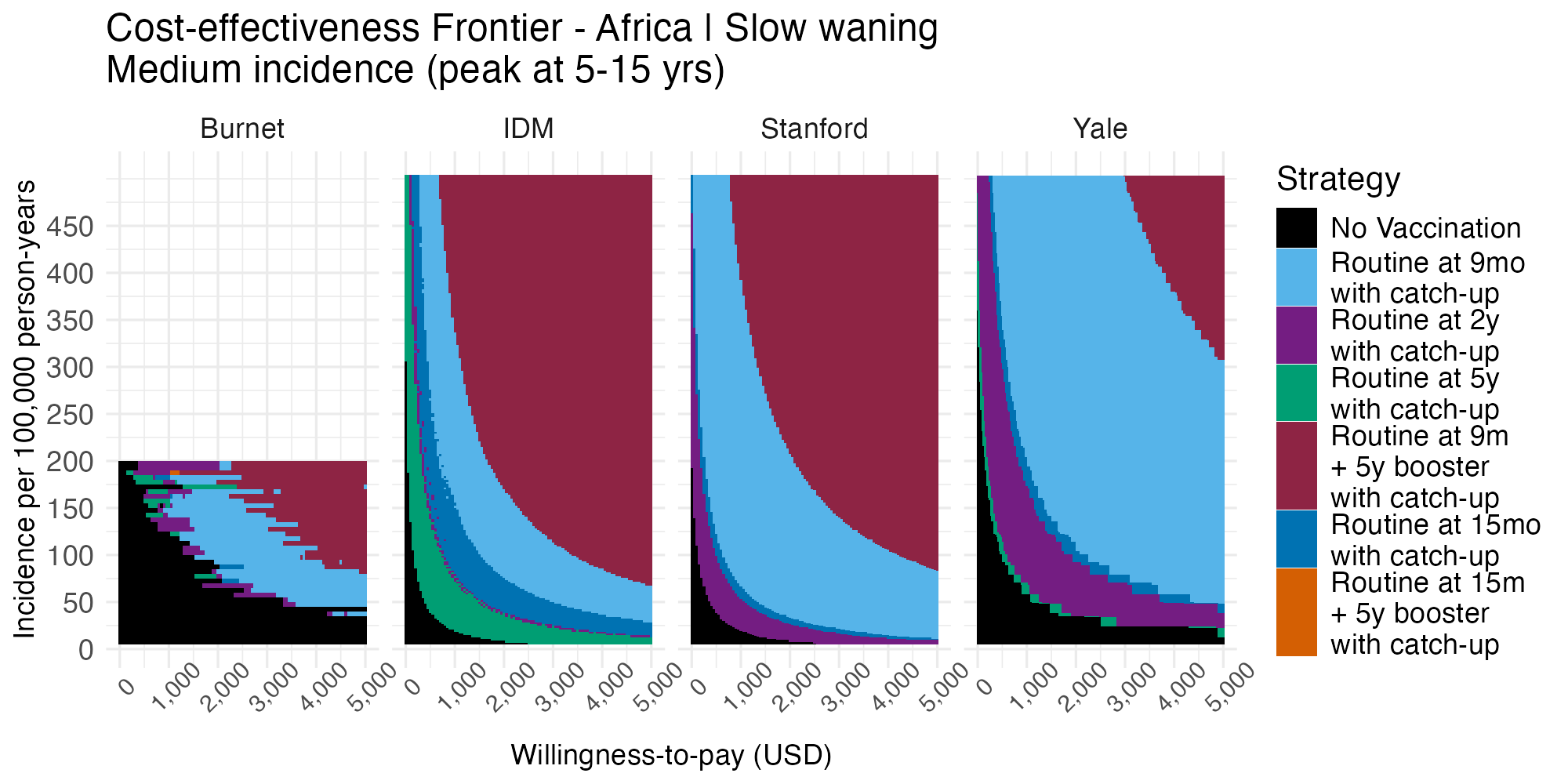

**Figure S7.7.3 Cost-effectiveness acceptability frontier across willingness-to-pay thresholds per disability-adjusted life years averted and incidence ranges for the fast-waning, higher-CFR/higher-cost Africa setting over a 10-year time horizon.** The optimal strategy predicted by each model is indicated by the color plotted at a given willingness-to-pay threshold per disability-adjusted life-year averted by the intervention.

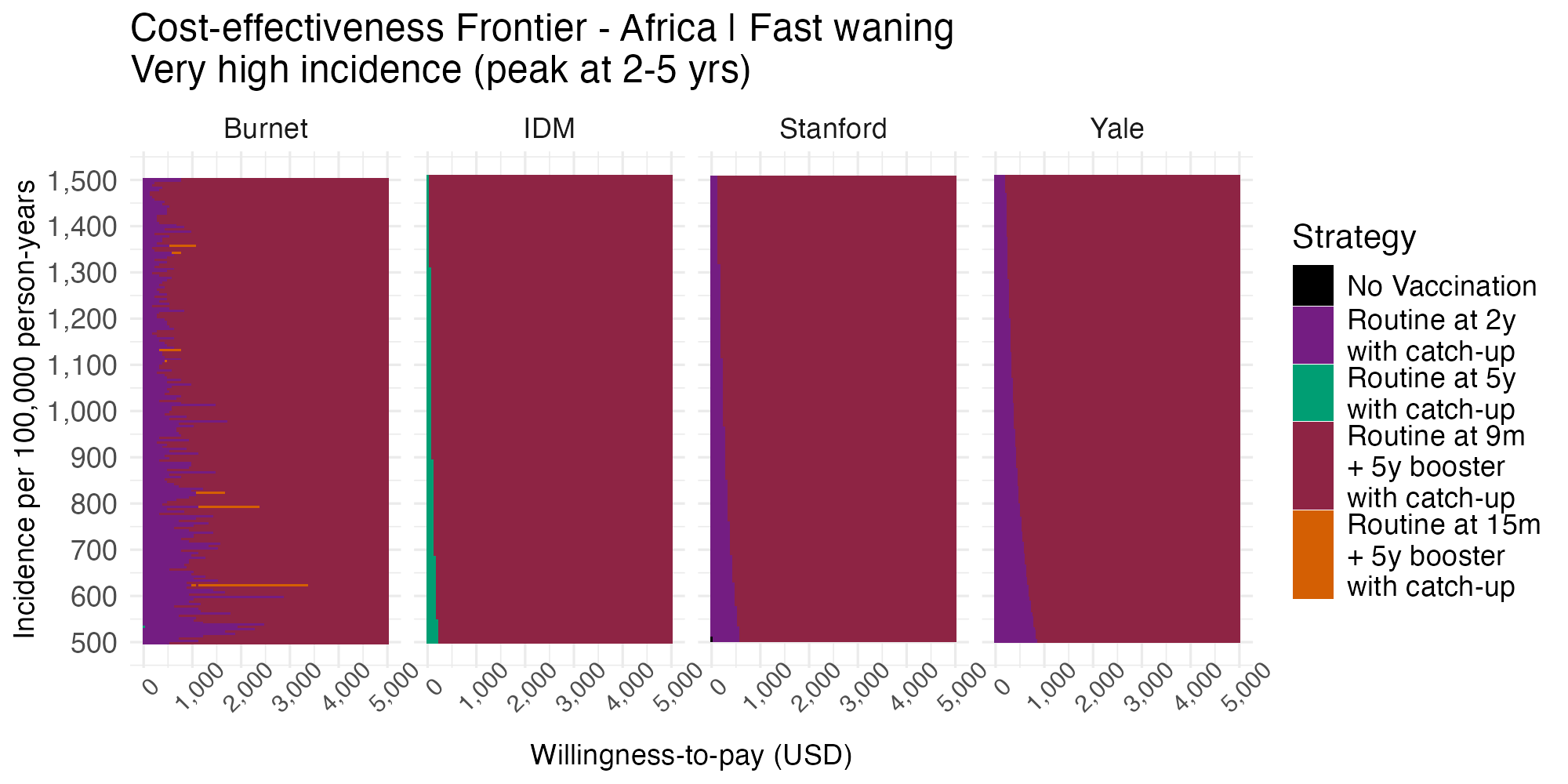

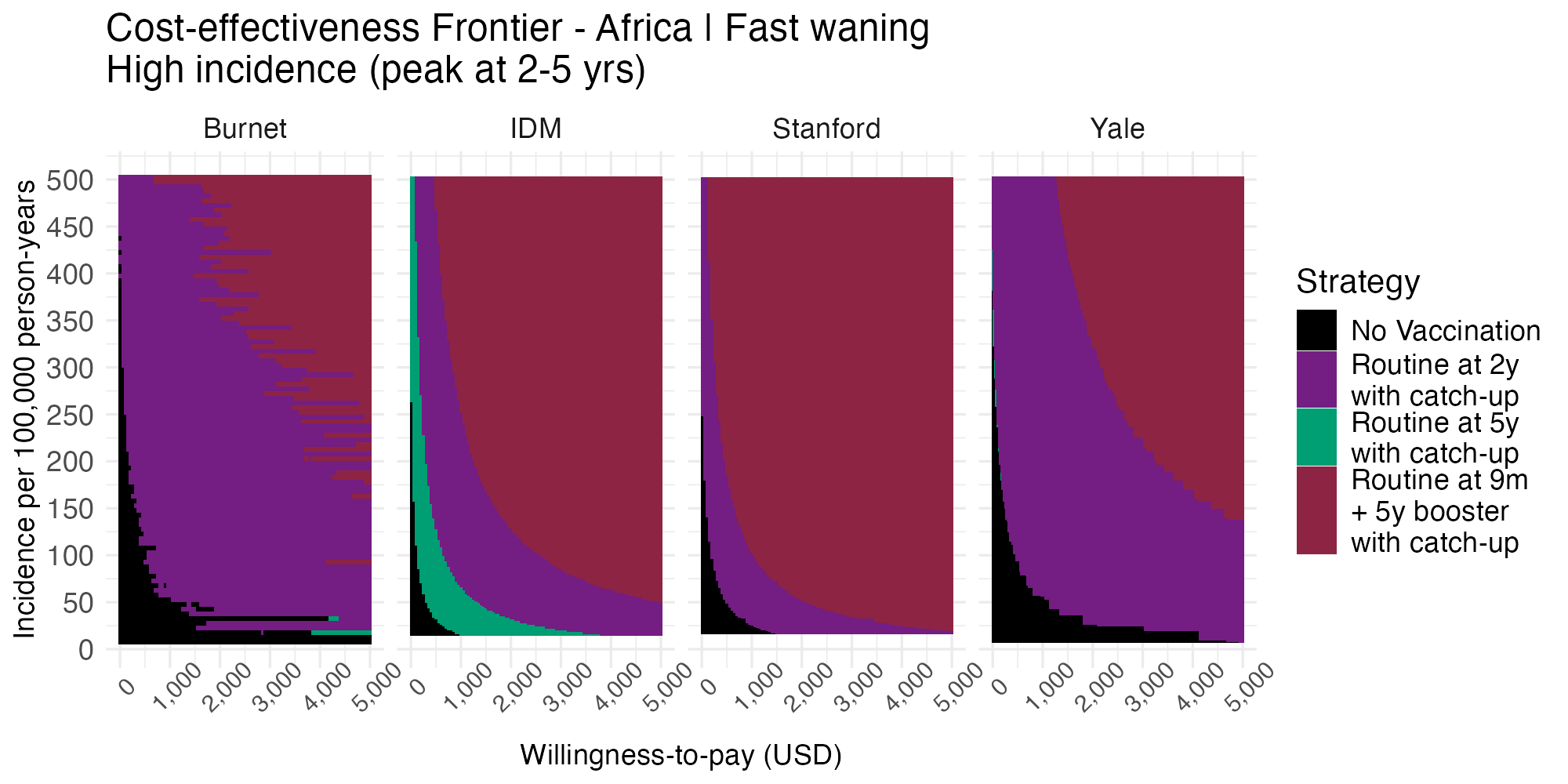

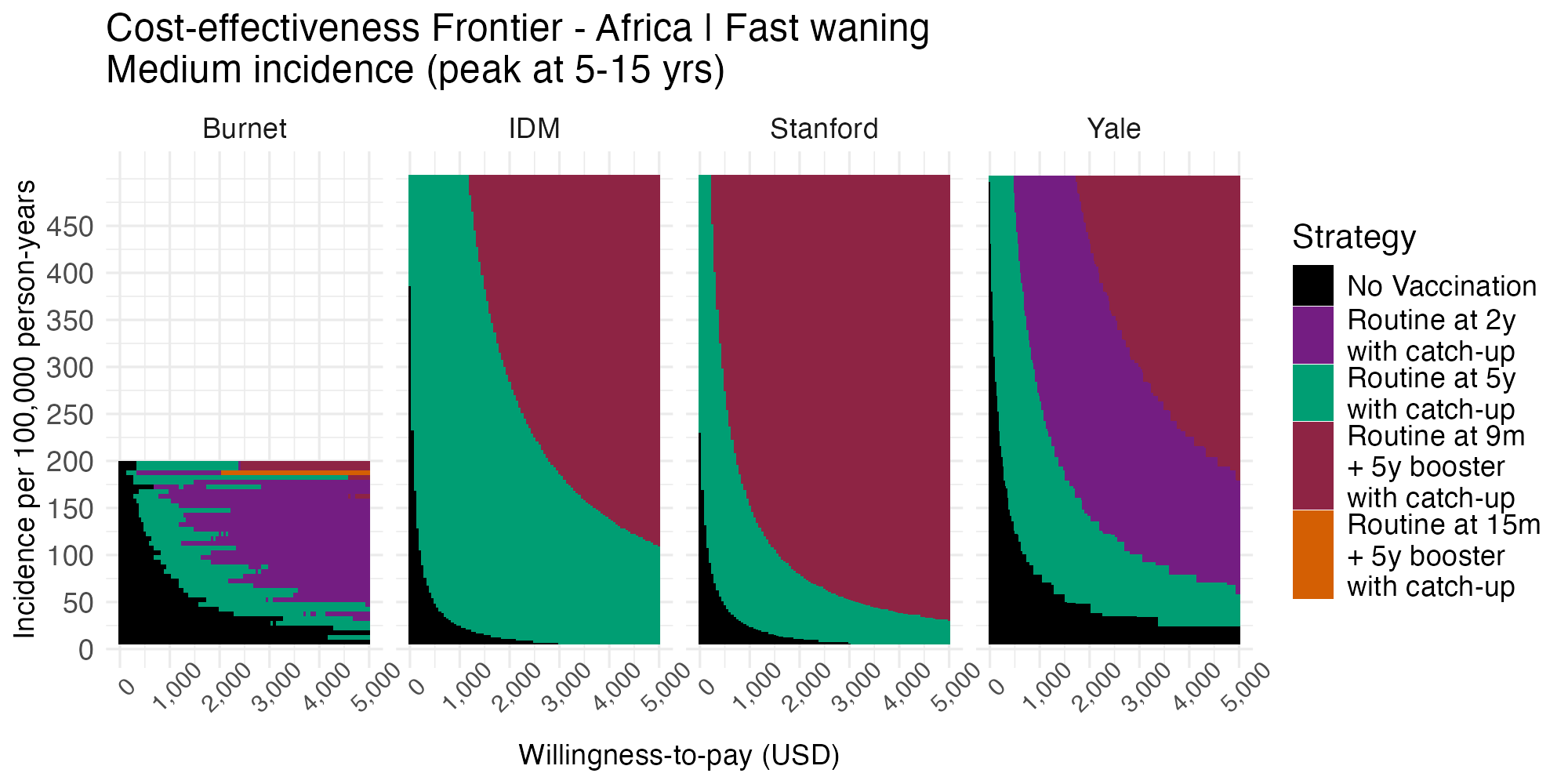

**Figure S7.7.4 Cost-effectiveness acceptability frontier across willingness-to-pay thresholds per disability-adjusted life years averted and incidence ranges for the slow-waning, lower-CFR/lower-cost Asia setting over a 10-year time horizon.** The optimal strategy predicted by each model is indicated by the color plotted at a given willingness-to-pay threshold per disability-adjusted life-year averted by the intervention.

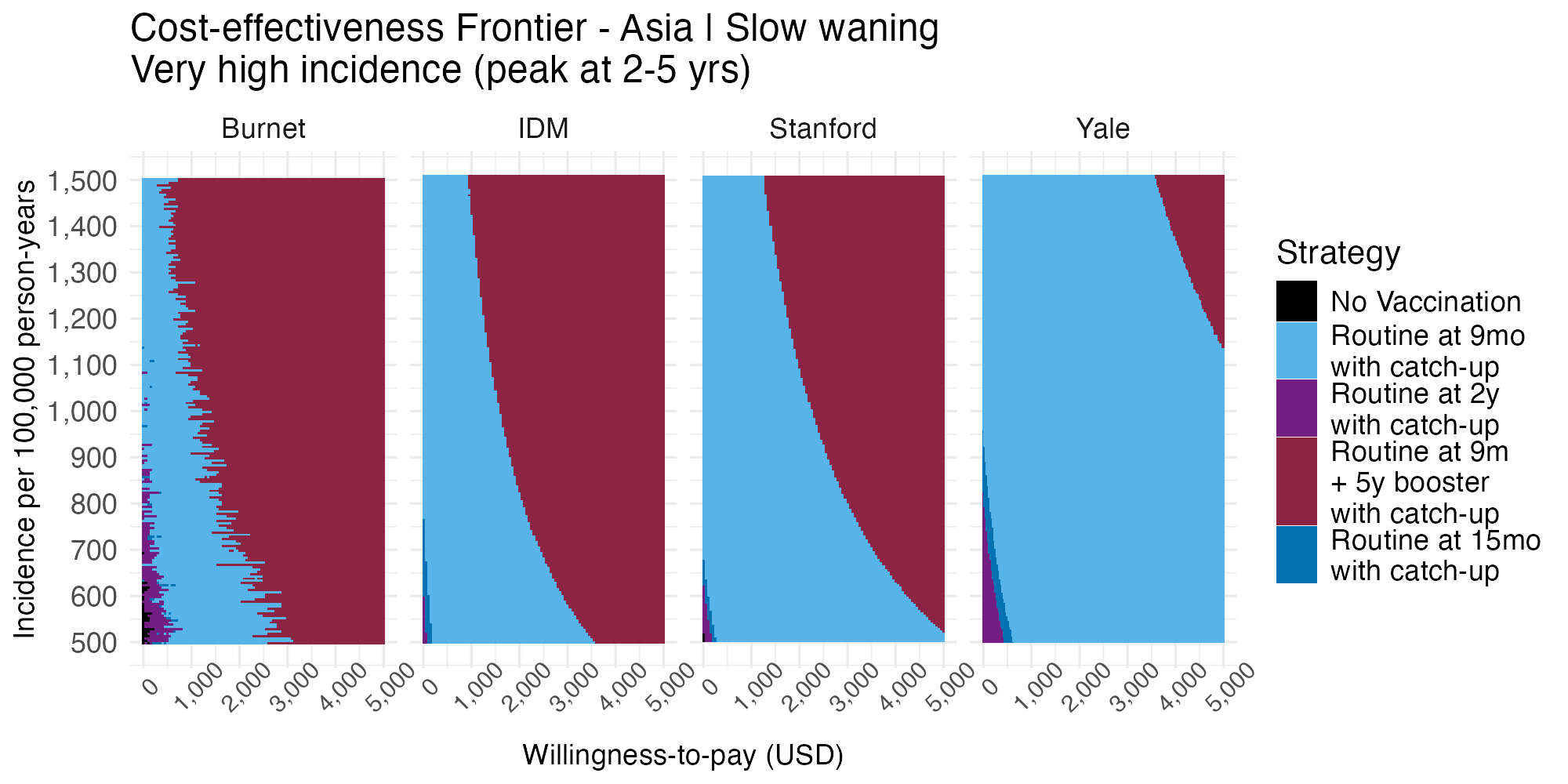

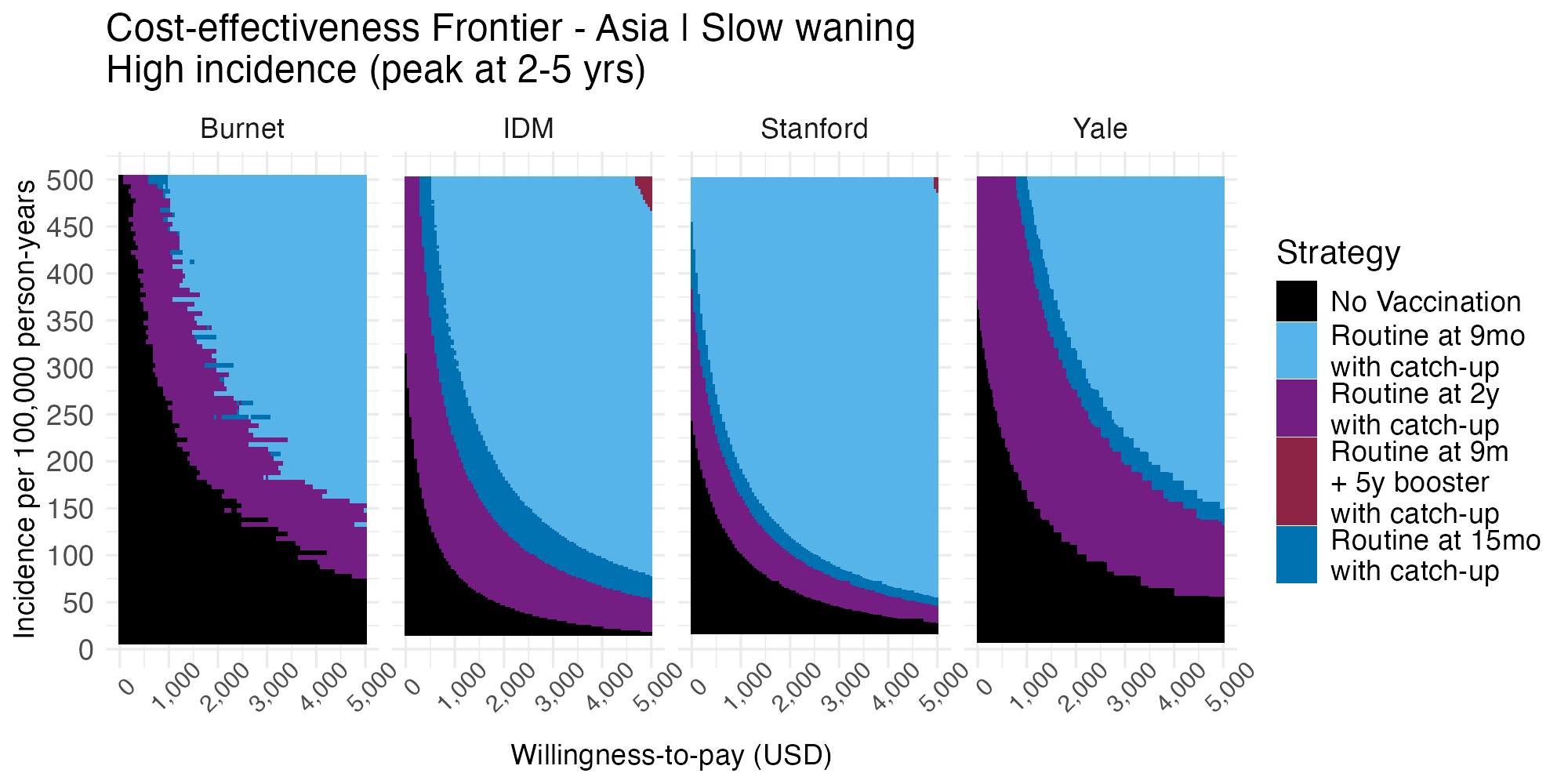

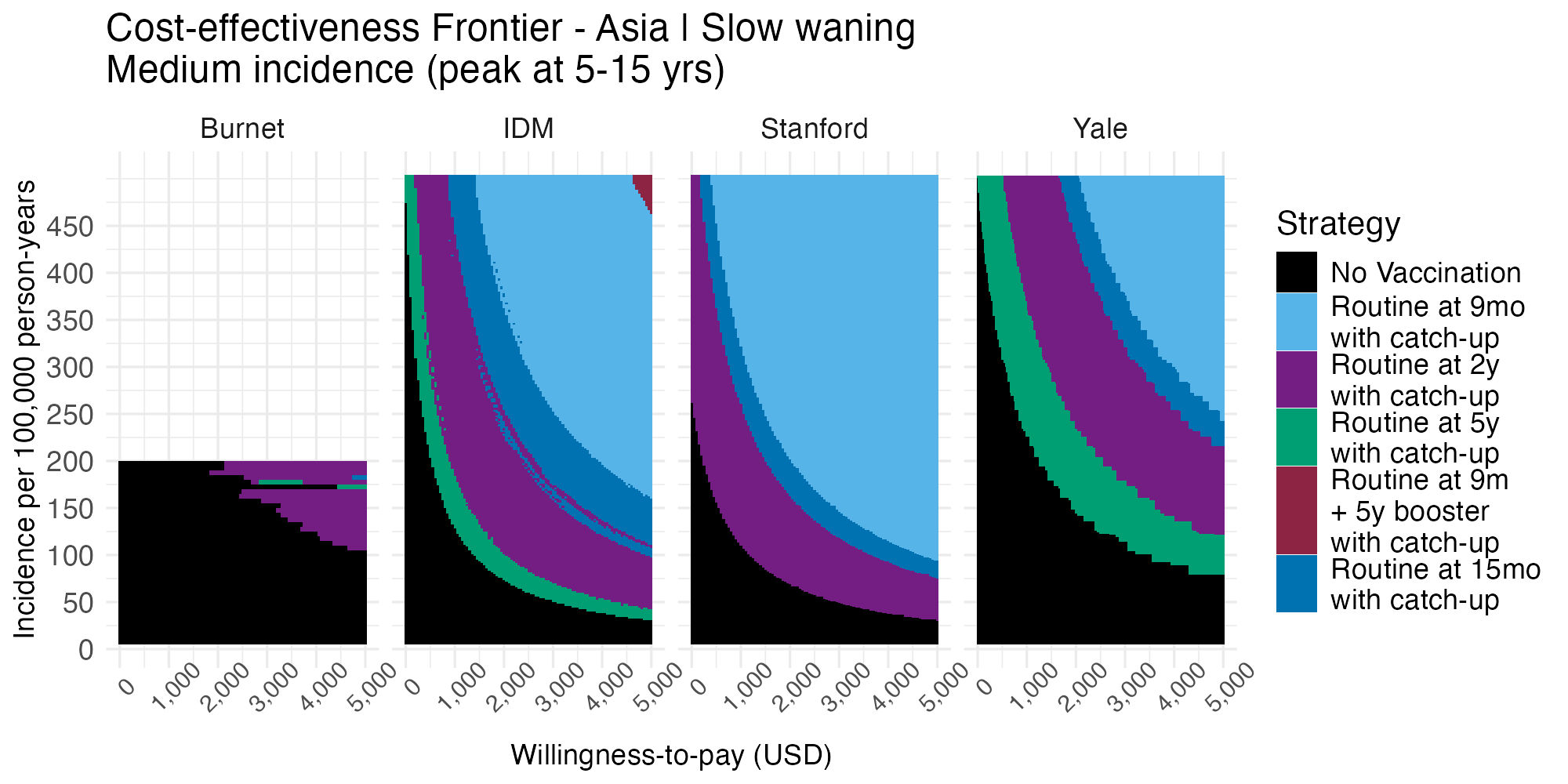

**Figure S7.7.5 Cost-effectiveness acceptability frontier across willingness-to-pay thresholds per disability-adjusted life years averted and incidence ranges for the fast-waning, lower-CFR/lower-cost Asia setting over a 10-year time horizon.** The optimal strategy predicted by each model is indicated by the color plotted at a given willingness-to-pay threshold per disability-adjusted life-year averted by the intervention.

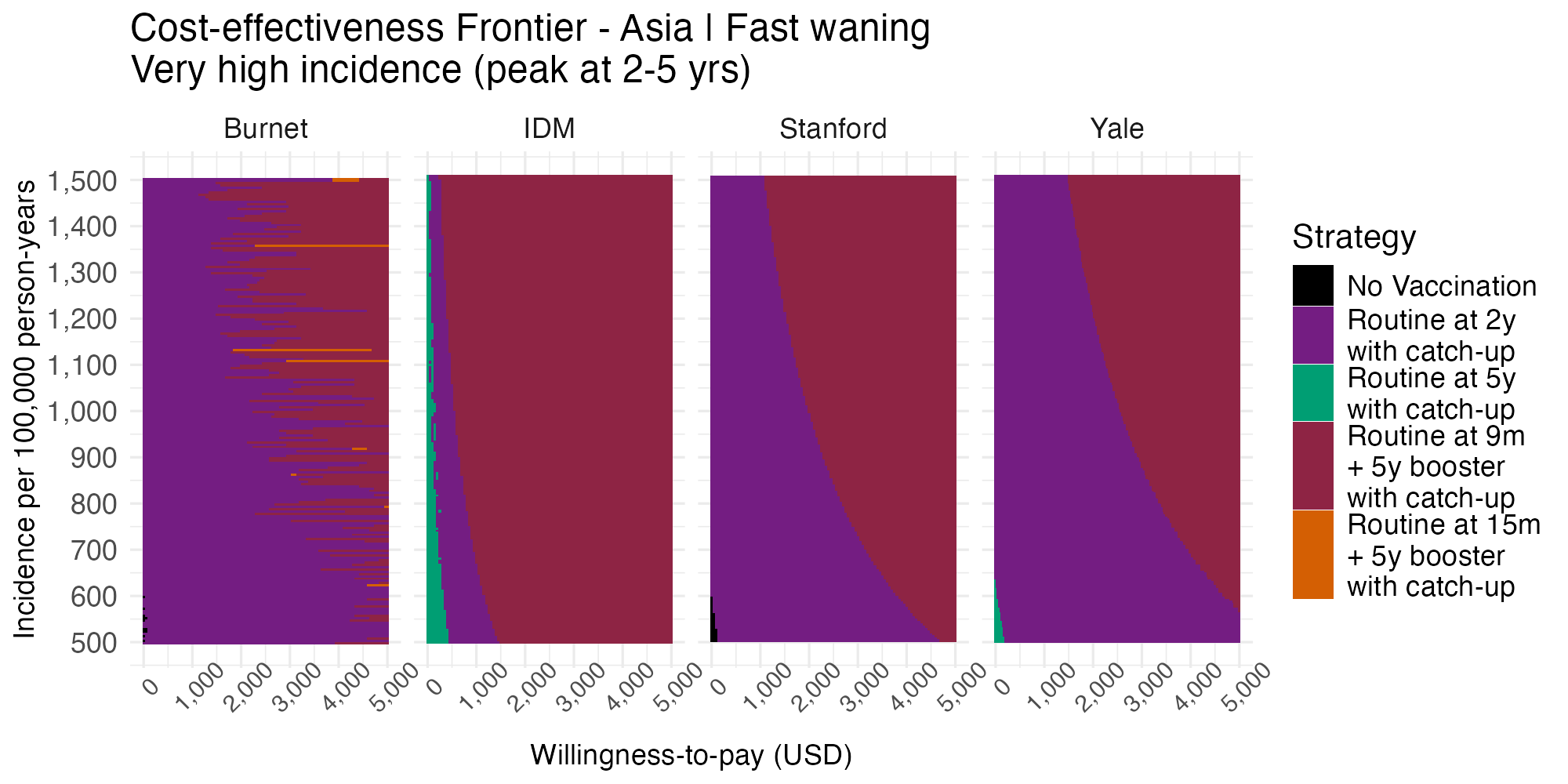

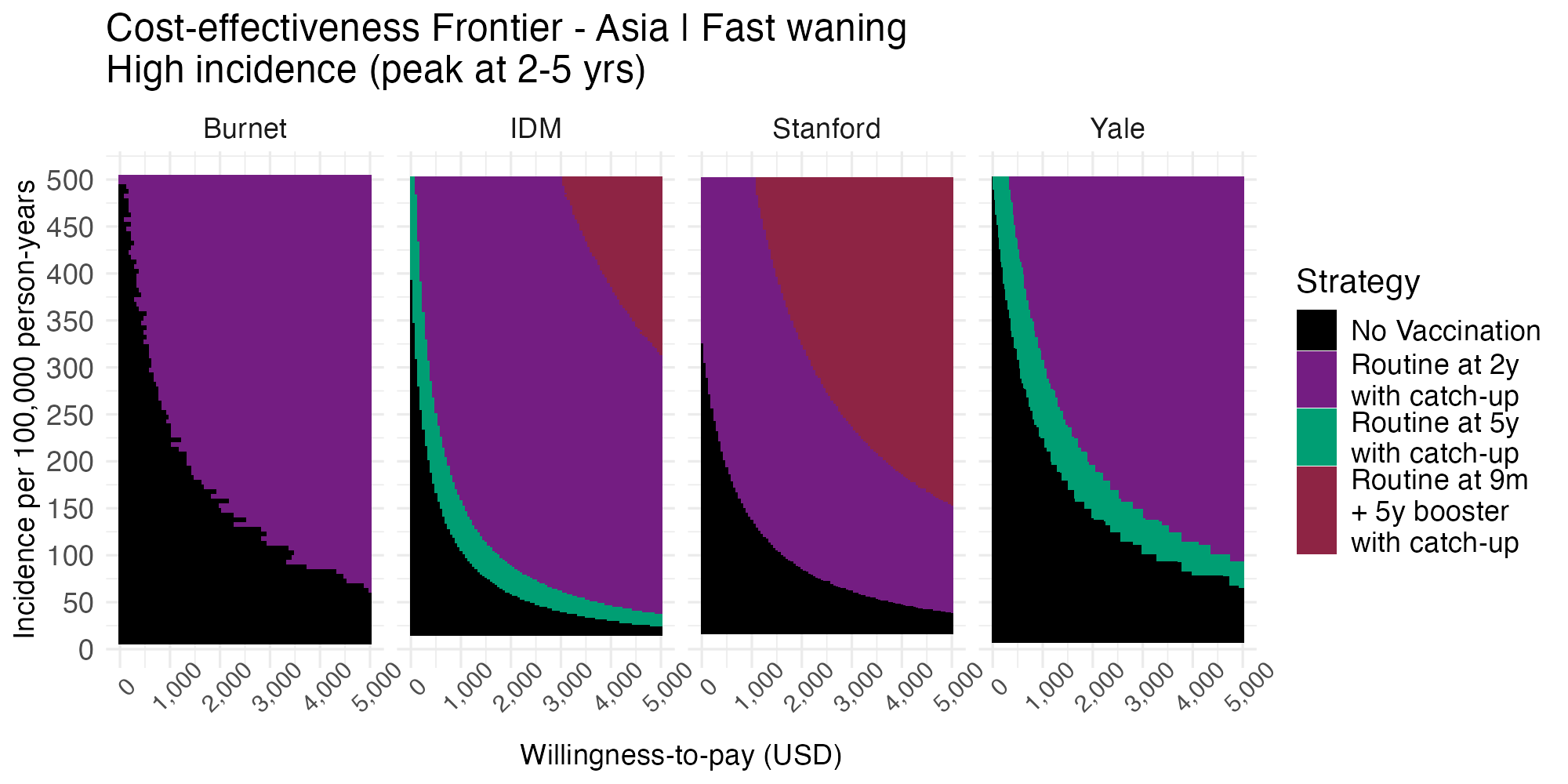

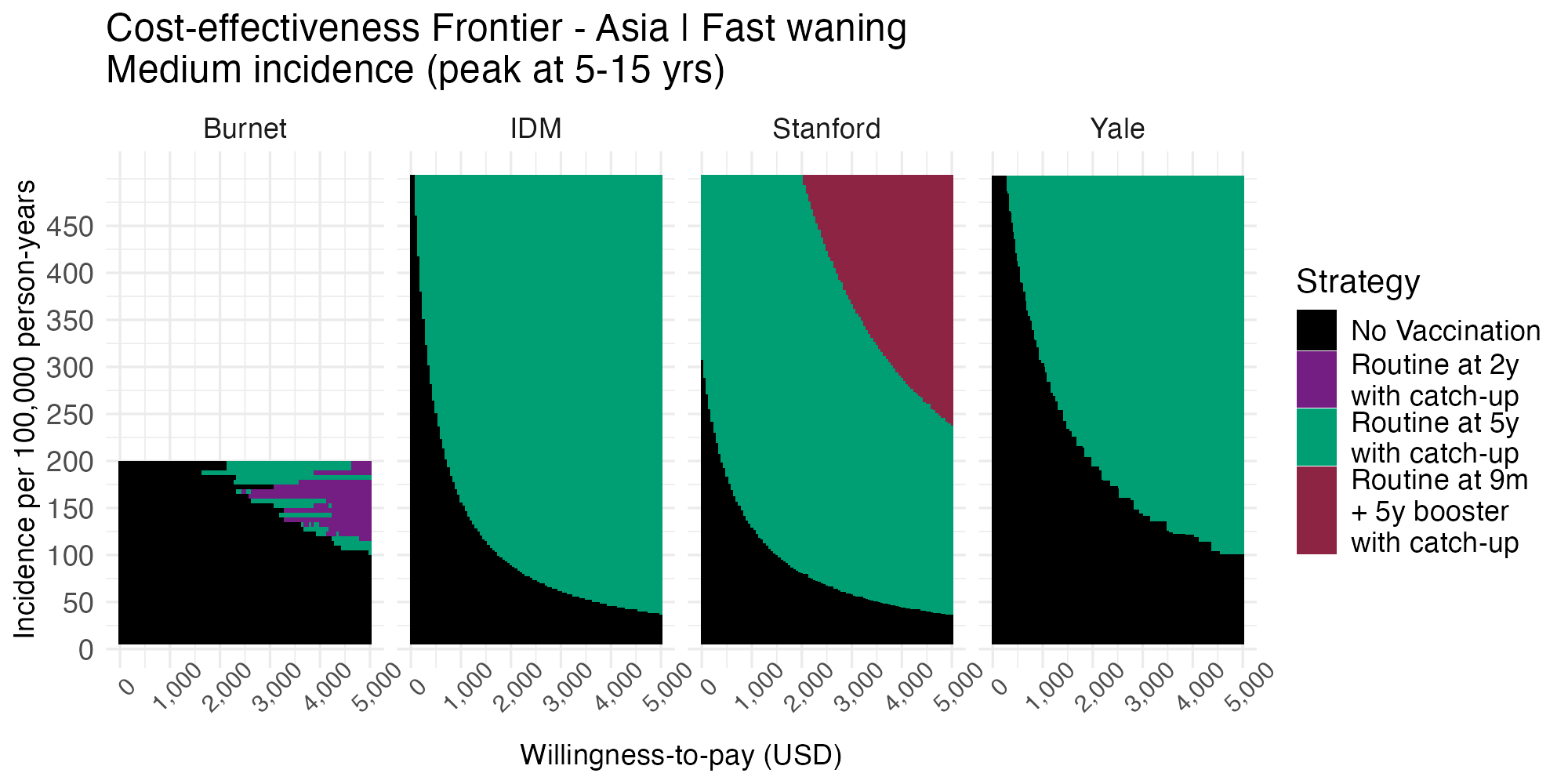

#### 7.8 Health savings compared to the cost of booster doses

**Figure S7.8.1 Cumulative 10-year costs of a 1-booster strategy compared to the cumulative health savings provided by the routine dose at 9 months in the slow-waning, higher-CFR/higher-cost Africa scenario across burden settings and models.**

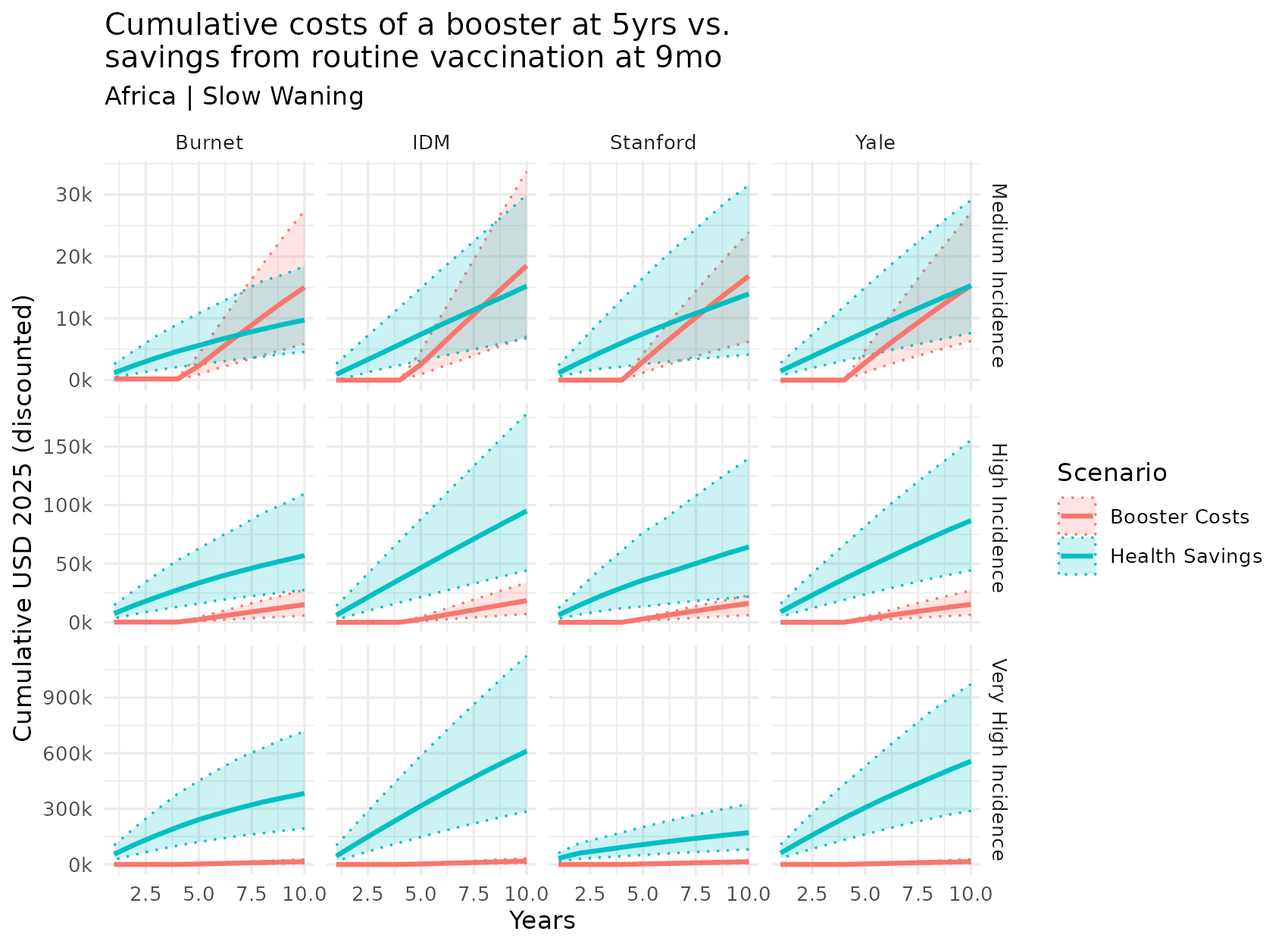

**Figure S7.8.2 Cumulative 10-year costs of a 1-booster strategy compared to the cumulative health savings provided by the routine dose at 9 months in the fast-waning, higher-CFR/higher-cost Africa scenario across burden settings and models.**

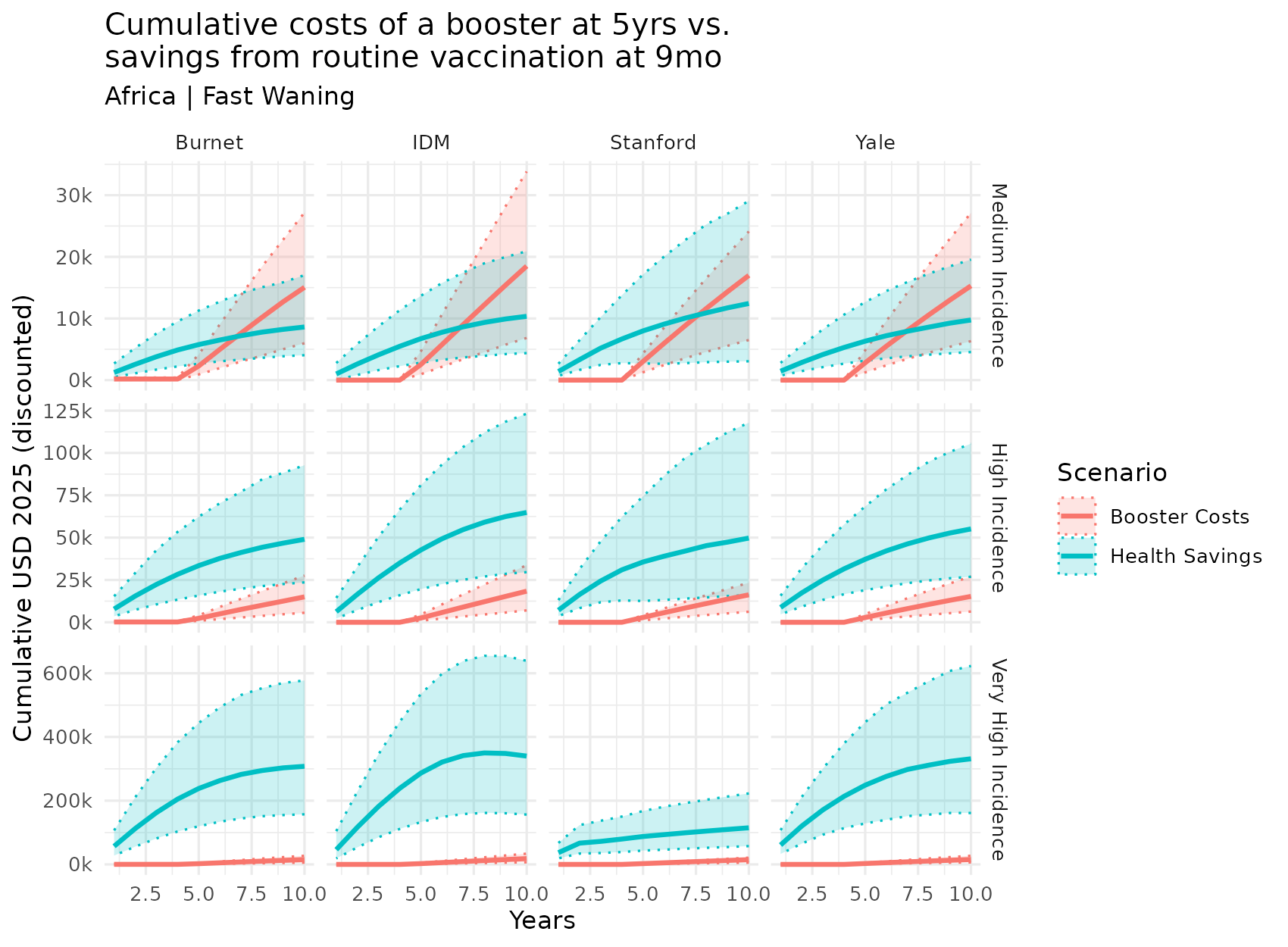

**Figure S7.8.3 Cumulative 10-year costs of a 1-booster strategy compared to the cumulative health savings provided by the routine dose at 9 months in the slow-waning, lower-CFR/lower-cost Asia scenario across burden settings and models.**

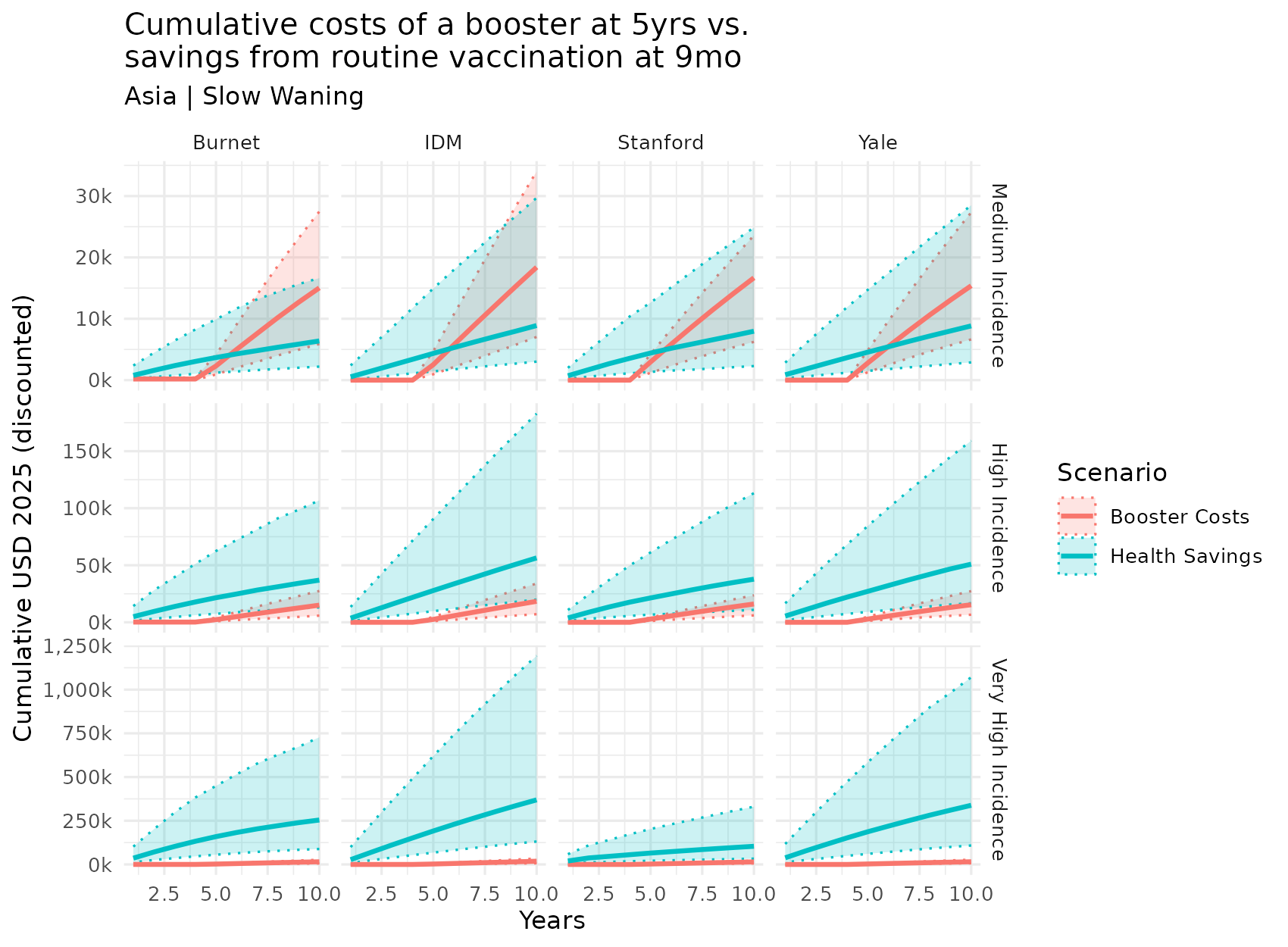

**Figure S7.8.4 Cumulative 10-year costs of a 1-booster strategy compared to the cumulative health savings provided by the routine dose at 9 months in the fast-waning, lower-CFR/lower-cost Asia scenario across burden settings and models.**

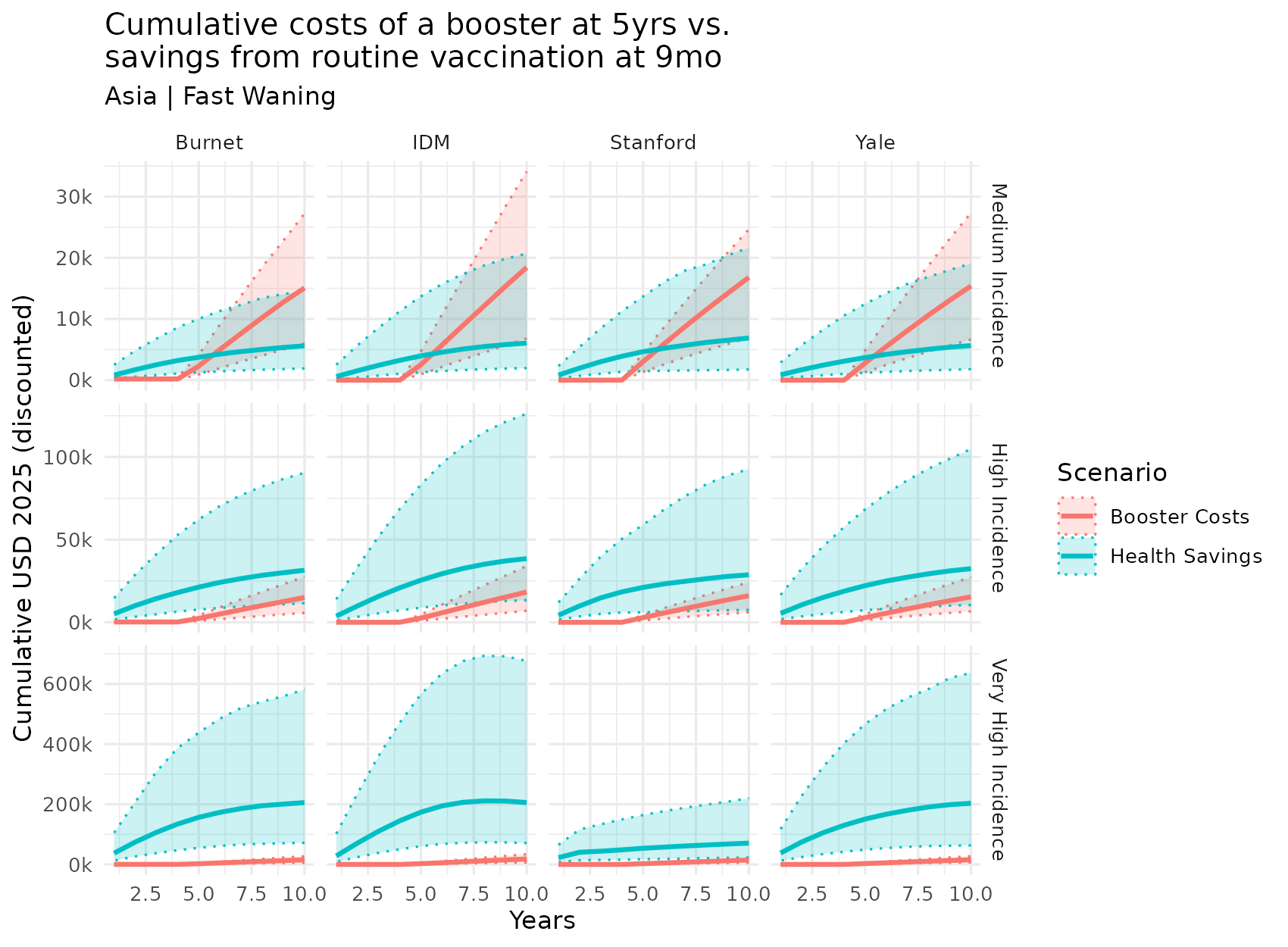

#### 7.9 One-way sensitivity analysis

**Figure S7.9.1 One-way sensitivity analysis tornado diagram for the high incidence, slow-waning, higher-CFR/higher-cost Africa scenario at a willingness-to-pay (WTP) threshold of $1,250.** This figure shows which parameters have the greatest influence on a strategy’s net monetary benefit (NMB) at the given WTP value. Parameters with a wider range of NMB have a greater influence on dictating the preferred strategy at that WTP. The colors indicate the positive or negative influence a parameter has on the NMB of a strategy.

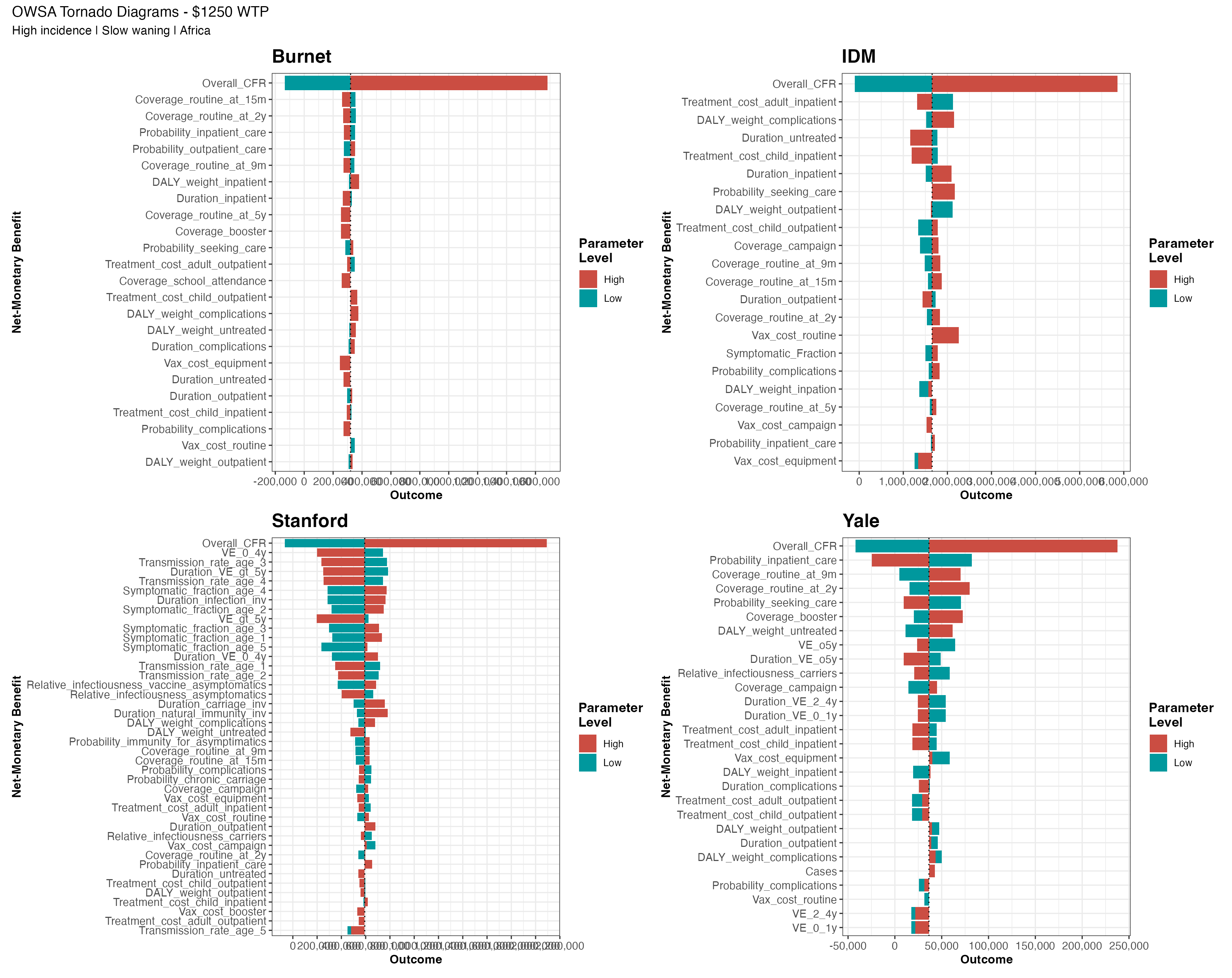

**Figure S7.9.2 One-way sensitivity analysis tornado diagram for the high incidence, fast-waning, higher-CFR/higher-cost Africa scenario at a willingness-to-pay (WTP) threshold of $1,250.** This figure shows which parameters have the greatest influence on a strategy’s net-monetary benefit (NMB) at the given WTP value. Parameters with a wider range of NMB have a greater influence on dictating the preferred strategy at that WTP. The colors indicate the positive or negative influence a parameter has on the NMB of a strategy.

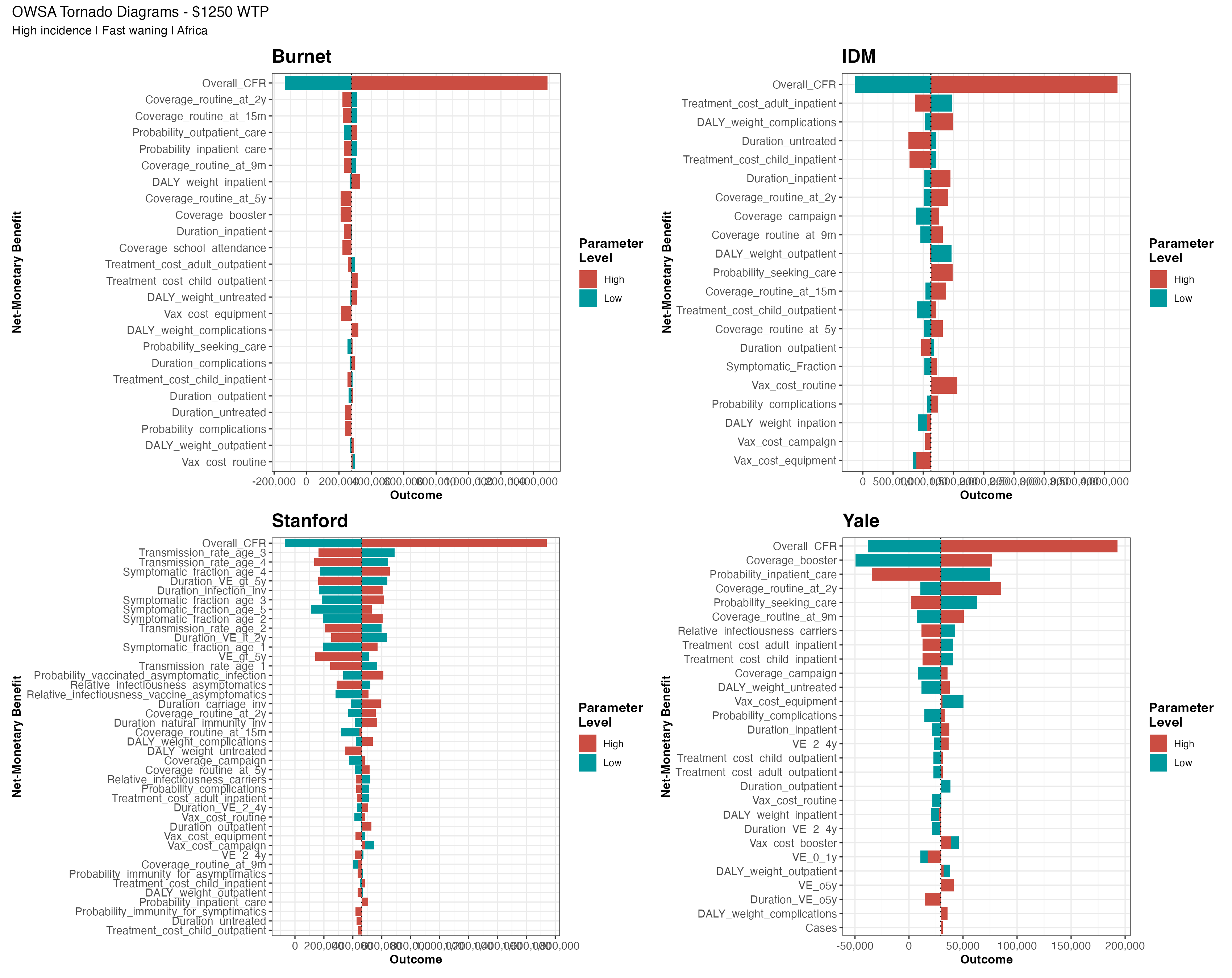

**Table S7.9.1 One-way sensitivity analysis parameter descriptions.**

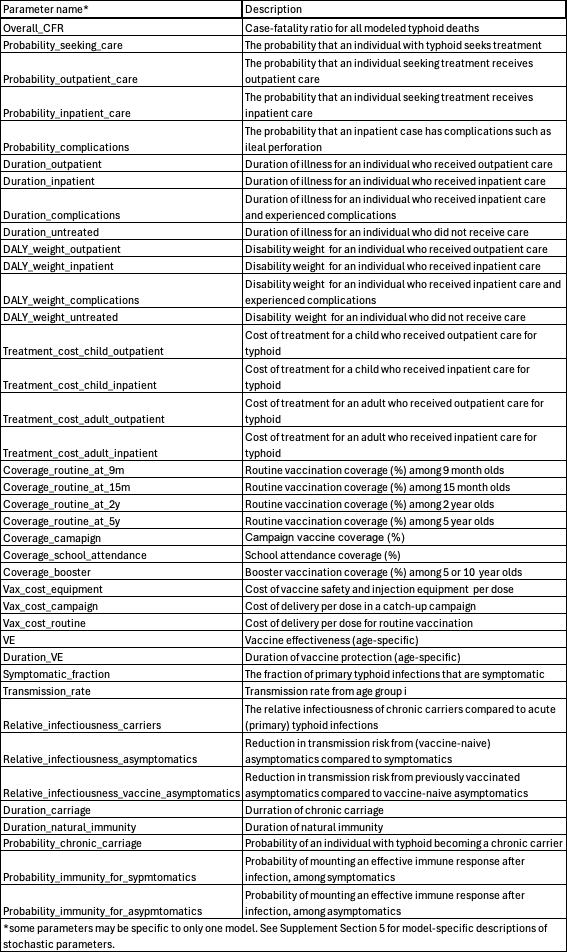
